# Understanding the Bidirectional Relationship Between Chronic Respiratory Disease and Cardiovascular Disease Using Genetic Evidence

**DOI:** 10.1101/2025.02.18.25322435

**Authors:** Naesilla, Jennifer Quint, Verena Zuber

**Author notes:** **Corresponding author:** Dr. Verena Zuber, School of Public Health, Imperial College London, White City Campus, 90 Wood Lane, London, W12 0BZ United Kingdom. Authors contributed equally (Jennifer Quint, Verena Zuber).

## Abstract

**Background:** Chronic respiratory diseases (CRD) and cardiovascular diseases (CVD) are leading global health burdens. Despite being common, CRD and CVD comorbidity is often underestimated due to overlapping symptoms and risk factors. Consequently, their relationship remains unclear.

**Aims and Objectives:** To determine the bidirectional genetic relationship between CRD and CVD and explore smoking and inflammation as potentially shared joint risk factors.

**Methods:** We conducted bidirectional Mendelian randomization (MR) to explore CRD-CVD relationships. Summary statistics from genome-wide association studies were retrieved for COPD, asthma, coronary artery disease (CAD), myocardial infarction (MI), heart failure (HF), atrial fibrillation (AF), and ischemic stroke (IS). We performed additional analysis including univariable MR for smoking, multivariable MR adjusting for smoking, and cis-MR to investigate the role of inflammatory markers.

**Results:** Our MR analysis found limited genetic evidence of relationships between CRD toward CVD, and vice versa. However, a nominally significant genetic association was observed between asthma and an increased risk of AF (OR_IVW_ 1.036, 95% CI: 1.003-1.070), remaining weakly significant after adjusting for smoking (OR_IVW_ 1.040, 95%CI 1.008 – 1.074). Genetically predicted lifetime smoking strongly increased all CRD and CVD risk. Additionally, genetically proxied IL6R concentration associated with increased asthma risk and decreased CAD, MI, AF, and IS risk, while IL1RN decreased COPD risk but increased CAD and MI risk.

**Conclusion:** While we found limited genetic evidence linking CRD and CVD, smoking and inflammatory markers commonly affect both. These findings highlight the complexity of CRD-CVD comorbidities, whose pathophysiology likely does not involve direct causation of each other.

## INTRODUCTION

Recognized as significant global burdens, cardiovascular diseases (CVD)—including coronary artery disease (CAD), myocardial infarction (MI), heart failure (HF), atrial fibrillation (AF), and stroke—and chronic respiratory diseases (CRD), such as asthma and chronic obstructive pulmonary disease (COPD), are currently key targets in the WHO 2023-2030 Global Action Plan for the Prevention and Control of Non-Communicable Diseases [1]. Despite the recognition of the significance of each disease, the coexistence between CVD and CRD has received relatively little attention. This is due to overlapping symptoms and risk factors, such as dyspnoea and smoking, which often results in underdiagnosis and undertreatment, eventually leading to increased hospitalisations, emergency room visits, mortality, and healthcare costs [2–5].

The uncertainty remains, whether one disease develops first and causes the other, or if they share a common pathway. Systemic inflammation and certain treatments, such as long-acting muscarinic antagonists (LAMA) or a long-acting beta-agonists (LABA), and inhaled corticosteroids, have been suggested as underlying factor contributing to the comorbidity between CRD and CVD, but studies report mixed effects on CVD risk, especially regarding the use of corticosteroids [6–9]. Observational studies have struggled to answer the question and demonstrate causality due to intrinsic biases, unmeasured confounding factors, and variability in reporting CRD cases [10,11]. Moreover, while observational studies are valuable, they often require long follow-up periods to detect chronic diseases and may still only reveal correlations, not true causal relationships. Interpreting associations as causal in these contexts is risky due to the potential for unmeasured confounding and reverse causation [12].

Here, Mendelian randomization (MR) offers a promising approach to triangulate evidence [12,13]. The MR study can provide stronger evidence for the potentially causal relationship between CRD and CVD by evaluating genetic liability data. Moreover, MR is more robust to address some limitations complicating observational studies with minimal ethical and practical constraints of clinical trials.

To date, MR studies have studied both asthma and COPD in relation to CVD comorbidities. However, the studies show inconsistent findings, with limited generalisability due to population-specific data and potential bias inherited from external GWAS source [14–18]. Numerous MR studies have also investigated the role of inflammatory markers in each CVD and CRD. Nevertheless, they have typically focused on each disease and marker separately. To our knowledge, no MR study has explored shared genetic risk factors related to inflammation in CRD and CVD comorbidity.

In this study, we aimed to address some gaps left by previous MR studies by examining CRD and CVD relationship using two-sample summary-level MR, focusing on high-burden CRD and CVD comorbidities. We want to determine whether a genetically predicted liability to certain CRD affects the risk of developing CVD and vice versa. Given the known role of smoking and inflammation in both CVD and CRD, and the variable responses to treatment, our study also aims to clarify the impact of smoking and inflammation markers. By employing updated GWAS sources with larger number of cases and using uniform analytical pipeline, we reduce researcher’s degree of freedom in individual studies and improve the finding’s reliability and provide a comprehensive analysis that agnostically explores the bi-directional relationship between CRD and CVD.

## METHODS

In brief, we retrieved publicly available summary statistics from larger, more recent and reliable GWAS if available from GWAS Catalog (https://www.ebi.ac.uk/gwas) [19], except where stated otherwise. There was no restriction on the ancestry in the GWAS to be included, but in the majority, we included European population, as it represents quite a large proportion in different available biobanks. All GWAS included are listed in Table 1.

**Table 1.**
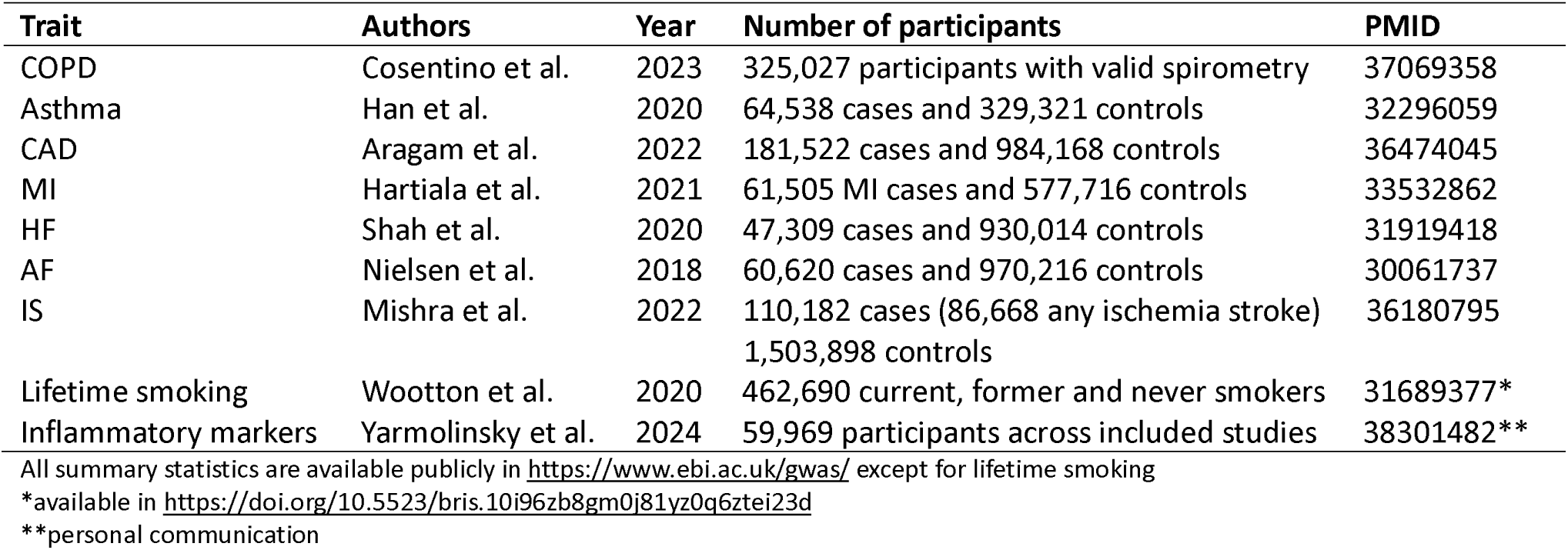
Data sources for current two-sample Mendelian Randomization analyses.

| Trait | Authors | Year | Number of participants | PMID |
| --- | --- | --- | --- | --- |
| COPD | Cosentino et al. | 2023 | 325,027 participants with valid spirometry | 37069358 |
| Asthma | Han et al. | 2020 | 64,538 cases and 329,321 controls | 32296059 |
| CAD | Aragam et al. | 2022 | 181,522 cases and 984,168 controls | 36474045 |
| MI | Hartiala et al. | 2021 | 61,505 MI cases and 577,716 controls | 33532862 |
| HF | Shah et al. | 2020 | 47,309 cases and 930,014 controls | 31919418 |
| AF | Nielsen et al. | 2018 | 60,620 cases and 970,216 controls | 30061737 |
| IS | Mishra et al. | 2022 | 110,182 cases (86,668 any ischemia stroke)<br>1,503,898 controls | 36180795 |
| Lifetime smoking | Wootton et al. | 2020 | 462,690 current, former and never smokers | 31689377* |
| Inflammatory markers | Yarmolinsky et al. | 2024 | 59,969 participants across included studies | 38301482** |

All summary statistics are available publicly in https://www.ebi.ac.uk/gwas/ except for lifetime smoking
\*available in https://doi.org/10.5523/bris.10i96zb8gm0j81yz0q6ztei23d
\*\*personal communication

### Instrument Selection

For the MR analysis, we selected genetic variants as instrumental variables that met three key assumptions for validity of relevance, independence assumption, and exclusion restriction [12,13,20]. Genetic variants or single nucleotide polymorphisms (SNP) with genome-wide significance threshold levels p-value < 5 × 10^-8^ were selected. To address the probability of non-Mendelian inheritance, we pruned for linkage disequilibrium (LD) between SNPs, with correlation coefficient threshold of r^2^ < 0.001 and a clumping window width of 10000 kb, based on European population reference. Finally, to avoid bias from weak instruments due to sample overlap, we calculated the F-statistic and excluded weak genetic variants with an F-statistic < 10 [21].

### Univariable Mendelian Randomization (UVMR)

After data pre-processing, we conducted two Univariable Mendelian Randomization (UVMR) as parts of the bidirectional univariate MR analysis for our primary objective. First, we examined the effect of the genetically predicted liability to each CRD trait on each CVD trait. Second, we conducted another UVMR where we flipped the direction of the analysis and re-selected the IV to represent the new exposure trait, in this case CVD. Our primary method for effect estimation was the inverse-variance weighted (IVW) approach. We included the sensitivity analyses to account for different assumptions of valid IVs and the potential of heterogeneity and pleiotropy, including MR-Egger, median-based methods, MR-PRESSO, and contamination mixture (CM) method [12,22,23].

As both CRD and CVD have smoking as a common risk factor, we conducted another MR analysis involving lifetime smoking as the exposure on each of CRD and CVD traits. The statistical methods employed here were the same as previously explained for bidirectional univariable MR analysis. For clarity, we referred to relationships as nominally significant if they met the p < 0.05 level without controlling for multiple testing. We did not adjust for multiple testing in UVMR, because the aim of our exploratory study was to detect any potential relationships between CRD and CVD traits keeping in mind the potential of false positive findings.

### Multivariable Mendelian Randomization (MVMR)

We conducted the MVMR analysis to assess the independent effect of each CRD and CVD (and vice versa) on each other, adjusting for smoking if the prior UVMR showed a nominally significant result. Similar to UVMR, our main method on MVMR was IVW (MVIVW) with MVMR-Egger to investigate the possibility of pleiotropy. The aim of MVMR analysis was to account smoking as common risk factor, rather than directed towards mediation analysis. To achieve this, genetic instruments were selected based solely on their association with the primary exposure of interest (either CRD or CVD). Variants associated with smoking were not specifically selected.

### Inflammation markers and CVD and CRD traits

We conducted cis-MR to investigate which inflammatory marker might be shared between CRD and CVD on cis genetic variants (variants in single gene region) as IVs for inflammation markers proxy [20]. These variants are located within or ±250 kb window from the gene encoding the relevant protein with weak LD (r^2^ < 0.10). The instrument selection process has been detailed in Yarmolinsky et al. [24].

The estimation of effect for cis-MR was as follows: if a single variant was available, we used the Wald ratio to estimate the causal effect of the inflammatory marker; otherwise, we used IVW techniques as the main effect estimation. We conducted sensitivity analyses accordingly, only when multiple independent IVs were available. Sensitivity analyses included MR-Egger, WM, MR-PRESSO, and CM.

We took into account multiple testing for the inflammation markers and corrected using false discovery rate (FDR) method, with q (adjusted p-value) < 0.05 indicating strong evidence and q-values between 0.05 and 0.20 considered suggestive [24].

### Software and Packages

All analyses are conducted in R software version 4.3.2. We used several packages in the analysis, including “ieugwasr” version 0.1.5 [25], “MendelianRandomization” package version 0.9.0 [26], and “MR-PRESSO” package version 1.0 [23].

## RESULTS

### Instrument Variables

For COPD, the number of IVs varied from 279 to 303, while for asthma, the number of IVs ranged from 140 to 146. All SNPs selected for COPD or asthma as exposure showed F-statistics greater than 10, indicating robust genetic instruments. Similarly, for CVD, each IV had an F-statistic higher than 10. The number of SNPs varied across traits, from 23 IVs for IS to 177 for CAD. The number of variables and respective F-statistics were presented in more detailed in the forest plot and Online Supplement Tables 1-6. For cis-MR, the number of IVs varied among 65 inflammation markers is presented in Online Supplement Table 7 - 8.

### How does the genetically predicted liability to COPD affect CVD?

Using the IVW method, we found no evidence that genetically predicted liability to COPD affects the risk of developing any CVD (CAD: OR (odds ratio)_IVW_ 0.984, 95% confidence interval (CI) 0.921 – 1.052, MI: OR_IVW_ 0.974, 95% CI 0.902 – 1.052, HF OR_IVW_ 1.037, 95% CI 0.974 – 1.103, AF: OR_IVW_ 1.039, 95% CI 0.968 – 1.114, IS OR_IVW_ 1.011, 95% CI 0.954 – 1.070) (Figure 1 and Online Supplement Figure 1). However, there was some inconsistency between genetically predicted liability to COPD and the risk of MI, CAD, and AF across sensitivity analyses. Specifically, while the IVW and MR-Egger found no significant association, the WM and CM methods suggested a potential decreased risk of MI associated with genetically liability to COPD. After removing 11 outliers identified by MR-PRESSO, the corrected IVW estimates supported a modest association between genetic liability to COPD and reduced risk of MI and CAD. All pairs were observed to have substantial heterogeneity, as shown by a high Cochrane’s Q test and p-het < 0.000 (Supplement Table 1).

**Figure 1.**
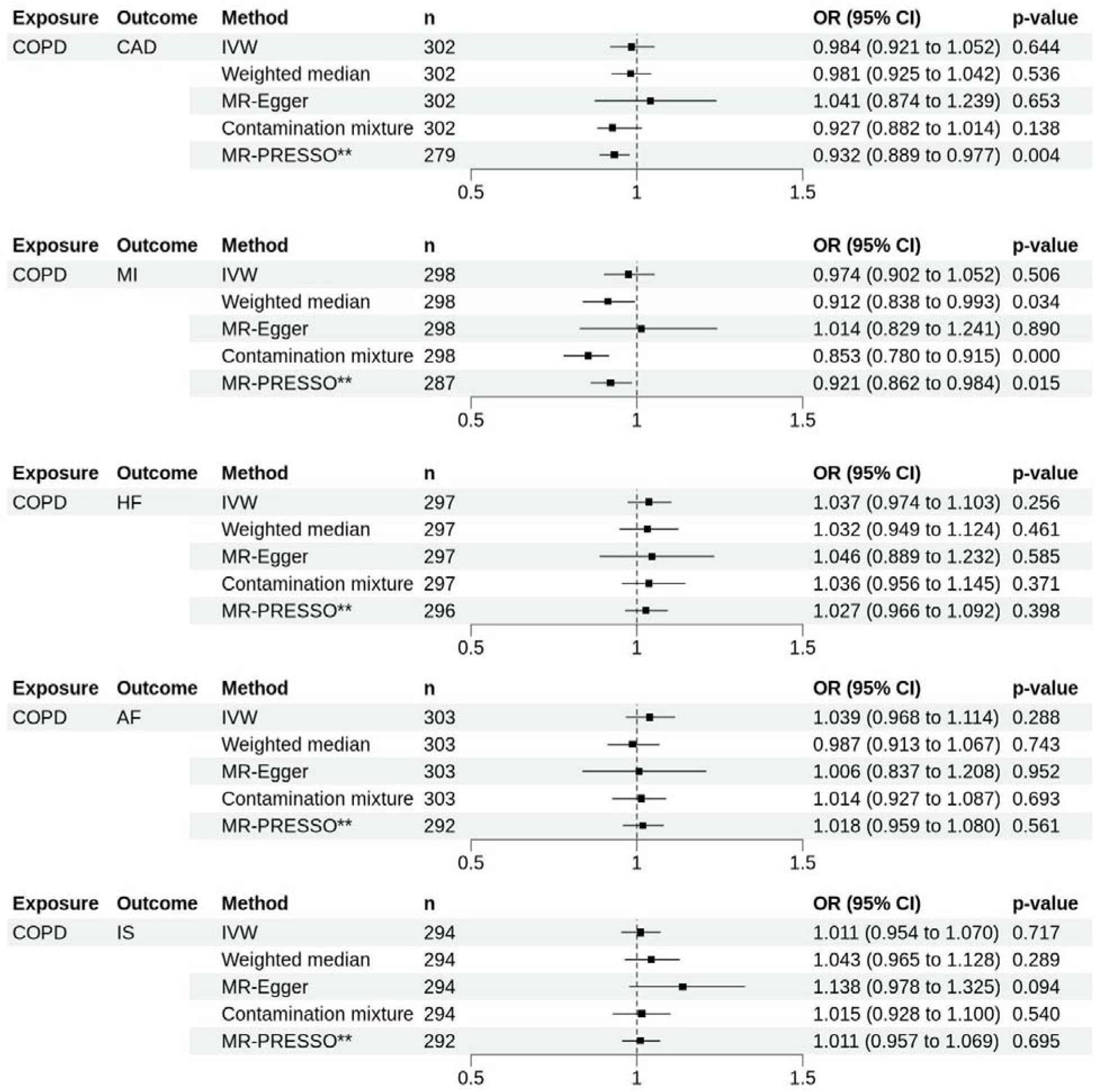
Forest plots showing the results of MR analyses on the effect of genetically predicted liability to COPD on the risk of cardiovascular diseases. Estimates are shown as OR. Horizontal lines represent the 95% CIs. AF: atrial fibrillation, CAD: coronary artery disease, COPD: chronic obstructive pulmonary disease, HF: heart failure, IS: ischemia stroke, MI: myocardial infarction, n: number of SNPs used as instrument variables in each method, SNPs single nucleotide polymorphism, CI confidence intervals, OR odds ratio. ** indicates corrected MR-PRESSO estimates

### How does the genetically predicted liability to asthma affect CVD?

We also examined how genetically predicted liability to asthma related to cardiovascular diseases (Figure 2 and Online Supplement Figure 2). The IVW analysis suggested a modest association between genetically predicted liability to asthma and increased odds of AF (OR_IVW_ 1.036, 95% CI 1.003-1.070). Nevertheless, while WM, MR-Egger, and CM did not show any significant association, MR-PRESSO supported the IVW finding after removing six potential outliers (OR_MR-PRESSO_ 1.036, 95% CI 1.008-1.064, P_CORRECTED_ = 0.012). Additionally, WM and CM methods showed suggestive evidence on the association of genetically predicted liability to asthma and increased risk of HF. Only one SNP was identified as an outlier by MR-PRESSO and removing it still did not indicate an association. Across analyses, consistent direction of estimates showed a lack of horizontal pleiotropy genetically predicted liability to asthma and the risk of HF and AF, supported by P_egger_intercept_. However, both noted the significantly high Cochrane’s Q test with p-het < 0.000 (Online Supplement Table 2).

**Figure 2.**
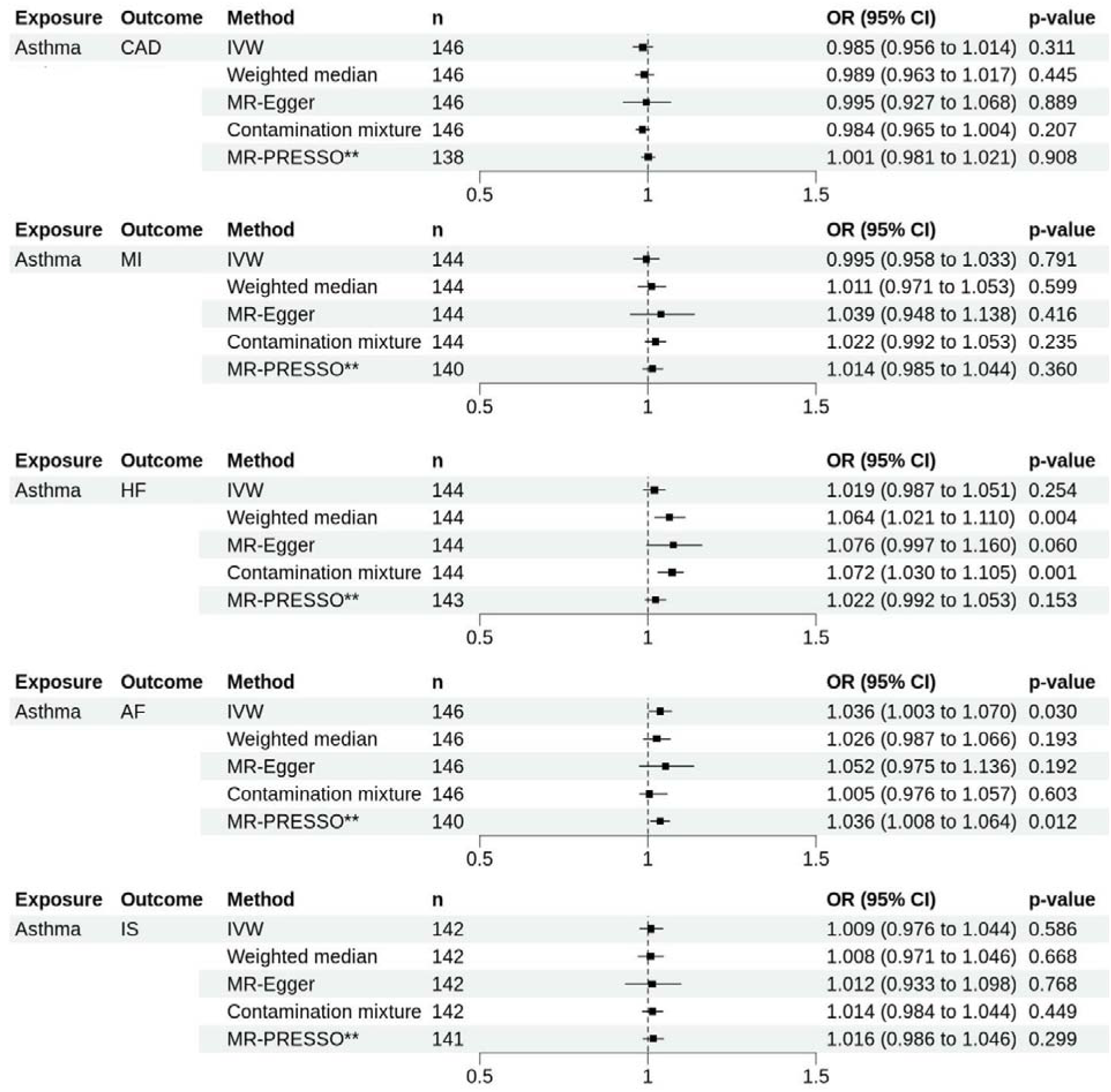
Forest plots showing the results of MR analyses on the effect of genetically predicted liability to asthma on the risk of cardiovascular diseases. Estimates are shown as OR. Horizontal lines represent the 95% CIs. AF: atrial fibrillation, CAD: coronary artery disease, HF: heart failure, IS: ischemia stroke, IVW: Inverse Variance Weighted. MI: myocardial infarction, n: number of SNPs used as instrument variables in each method, SNPs single nucleotide polymorphism, CI confidence intervals, OR odds ratio. ** indicates corrected MR-PRESSO estimates

### How does the genetically predicted liability to CVD affect COPD or asthma?

We did not identify any significant evidence supporting an effect of genetically predicted liability to any CVD on the risk of developing COPD or asthma using IVW analysis (Online Supplement Figure 3 – 6 and Online Supplement Table 3 - 4). Sensitivity analyses, including WM, MR-Egger, and CM methods, consistently reflected these findings.

### How does the genetically predicted lifetime smoking index affect each CRD and CVD traits?

The IVW analysis indicated strong evidence on the association between genetically predicted lifetime smoking index and increased COPD odds by 9.5% per 1 standard deviation (OR_IVW_ 1.095, 95% CI 1.032 to 1.162, P_IVW_ = 0.003) and moderate evidence for increased asthma odds by 21% (OR_IVW_ 1.207, 95% CI 1.019 to 1.428, P_IVW_ = 0.029). Our IVW analysis also revealed that genetically predicted lifetime smoking was associated with increased odds of all types of CVD included (Figure 3 and Online Supplement Figure 7). Sensitivity analyses showed consistent directions, though P_egger_intercept_ revealed the possibility of horizontal pleiotropy on the effect of lifetime smoking index on COPD, CAD and MI (Online Supplement Table 5). MR-PRESSO identified several potential outliers and upon those SNPs removal, the result still showed a strong genetic association between smoking and COPD, asthma, and all CVDs.

**Figure 3.**
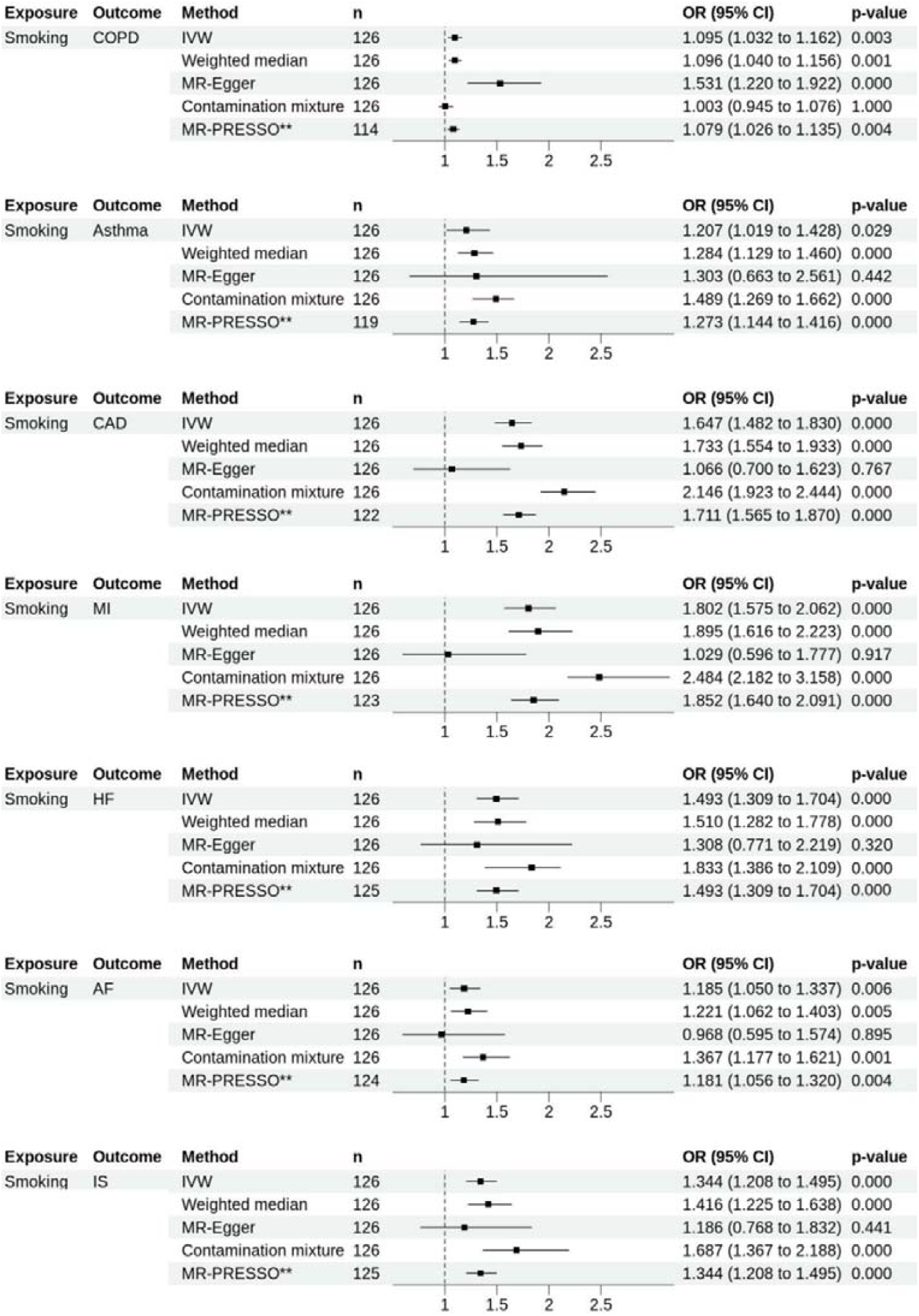
Forest plots showing the results of MR analyses on the effect of genetically predicted lifetime smoking on the risk of developing chronic respiratory diseases and cardiovascular diseases. Estimates are shown as OR. Horizontal lines represent the 95% CIs. AF: atrial fibrillation, CAD: coronary artery disease, HF: heart failure, IS: ischemia stroke, MI: myocardial infarction, n: number of SNPs used as instrument variables in each method, SNPs single nucleotide polymorphism, CI confidence intervals, OR odds ratio. ** indicates corrected MR-PRESSO estimates.

### Multivariable MR Analysis

Considering the close relationship between lifetime smoking toward COPD and asthma previously, we extended the analysis to account for lifetime smoking as a common risk factor in MVMR for COPD toward MI, and asthma towards AF and HF. From these MVMR analyses, we found that the effect of asthma on HF and COPD on MI were weakened when adjusted for lifetime smoking (Online Supplement Table 6). Meanwhile, the effect of asthma on AF was stronger when conditioning on smoking (Figure 4). We did not detect significant horizontal pleiotropy, as indicated by the P_egger_intercept_. In conclusion, our MVMR analysis provides evidence that when accounting for smoking, asthma or COPD may not affect the risk of developing CVDs, except AF.

**Figure 4.**
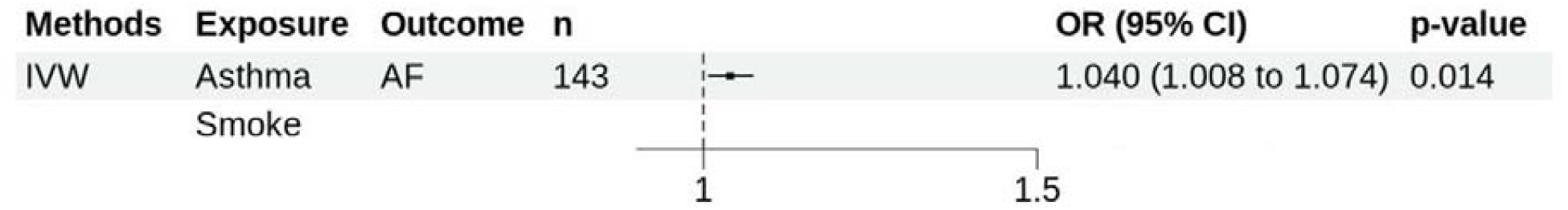
Multivariable Mendelian randomisation analysis results of the effect of genetically predicted liability to asthma towards AF adjusted for lifetime smoking. Estimates are shown as OR. Horizontal lines represent the 95% CI. AF: atrial fibrillation, MVIVW: multivariable inverse-variance weighted, n: number of SNPs used as instrument variables in each method, SNPs single nucleotide polymorphism, CI confidence intervals, OR odds ratio.

### How does the genetically proxied inflammatory markers affect CRD and CVD?

Based on IVW results, we found 12 genetically proxied cytokines and chronic respiratory diseases pairs and 24 genetically proxied cytokines and cardiovascular diseases pairs with strong evidence of an association after accounting for multiple testing adjustment. Among these inflammatory markers, we found several inflammatory markers association overlapping between CRD and CVD, presented in Figure 5 and Online Supplement Table 7.

**Figure 5.**
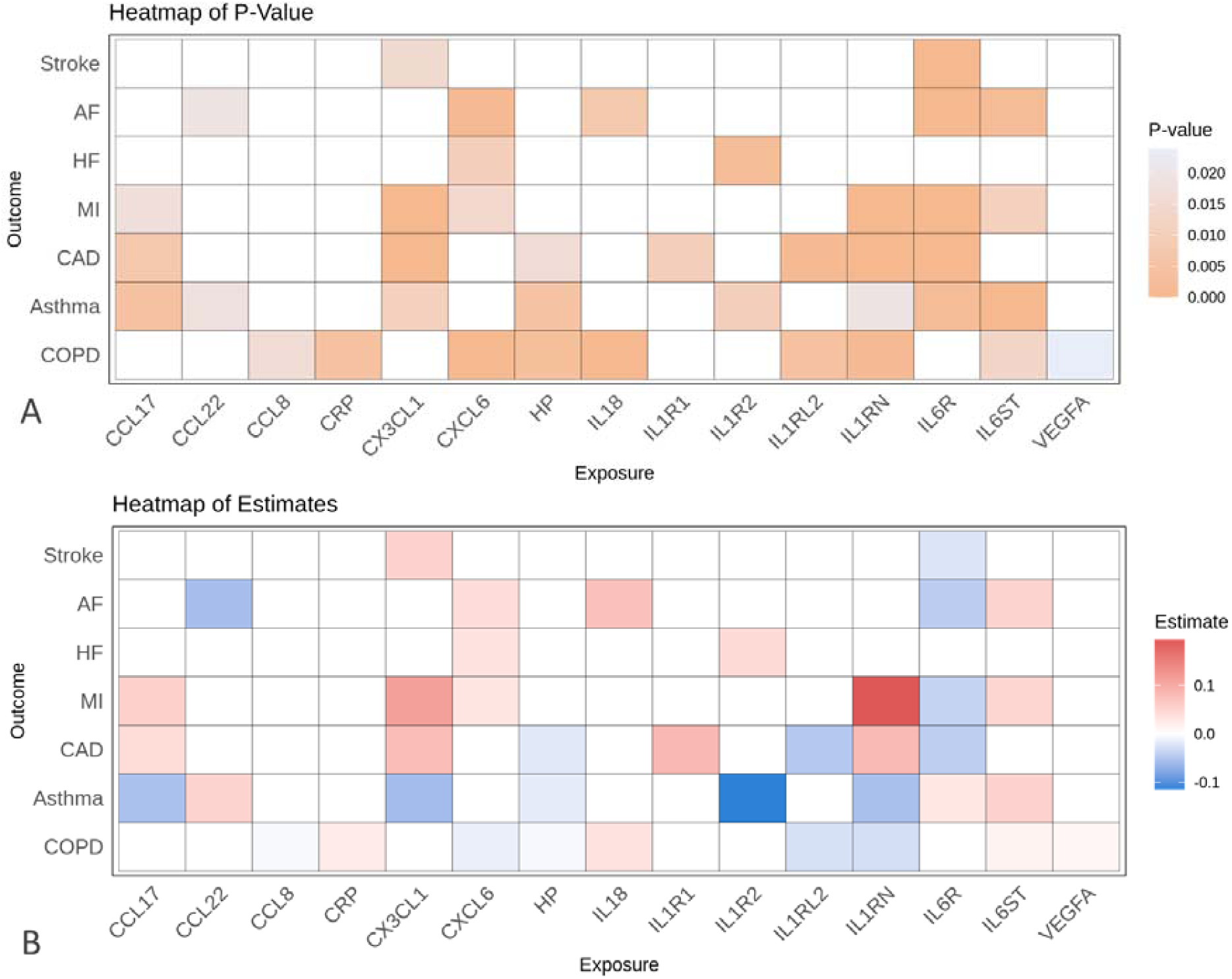
Heatmap of cis-MR analysis results of the effect of genetically predicted inflammatory markers on chronic respiratory disease and cardiovascular disease, based on (A) the effect estimate and (B) p-value from inverse weighted variance analysis. AF: atrial fibrillation, CAD: coronary artery disease, CCL: CC chemokine ligand, CXCL: CXC Chemokine Ligand, HF: heart failure, HP: haptoglobin, IL: interleukin, IL6R: IL6 Receptor, IL6ST: IL6 Cytokine Family Signal Transducer, IL1RN: IL1 receptor antagonist, IL1RL2: IL1 receptor-like 2, IL1R2: IL1 Receptor Type 2, IS: ischemia stroke, MI: myocardial infarction.

Two markers, IL6 Receptor subunit alpha (IL6R) and IL1 receptor antagonist (IL1RN), showed significant results. The results showed strong genetic evidence (q-value < 0.05) that IL6 Receptor subunit alpha (IL6R) concentration was associated with decreased risk of most of CVDs including CAD (OR_IVW_ 0.958, 95% CI 0.947 – 0.968), MI (OR_IVW_ 0.963, 95% CI 0.949 – 0.976), AF (OR_IVW_ 0.957, 95% CI 0.943 – 0.971) and IS (OR_IVW_ 0.977, 95% CI 0.965 – 0.989), but increased odds of asthma (OR_IVW_ 1.029, 95% CI 1.010 −1.049). Meanwhile, we found genetically predicted IL1 receptor antagonist (IL1RN) concentration to be strongly associated with increased odds of CAD (OR_IVW_ 1.087, 95% CI 1.046–1.129) and MI (OR_IVW_ 1.214, 95% CI 1.142–1.291) and decreased odds of COPD (OR_IVW_ 0.972, 95% CI 0.955–0.989) and weakly associated with decreased odds of asthma. Sensitivity methods for all pairs showed consistent direction of estimate, further supported by the MR-Egger p-intercept > 0.05, indicating no horizontal pleiotropy.

Other associations with strong evidence that overlapped between inflammatory markers and CVD and inflammatory markers and CRD in IVW analysis included CXCL6 and IL6ST. Additionally, there were mixed overlapping between strong and suggestive evidence between CRD and CVD including IL18, CX3CL1, IL1R2, HP, and IL18. Additional forest plots for these overlapping inflammatory markers are all shown in Online Supplement Figure 8 and Online Supplement Table 8.

## DISCUSSION

Few MR studies have utilised large GWAS to investigate the bidirectional effects and associations between CRD and CVD. We found no strong genetic evidence to support associations between genetically predicted liability to CRD and the risk of developing CVD. Similarly, we did not find significant genetic evidence indicating that CVD liability increases the risk of COPD or asthma. However, we confirmed that genetic liability to lifetime smoking strongly affects the risk of both CRD and CVD. Furthermore, out of 65 inflammatory markers analysed, we identified several with strong genetic associations related to both CRD and CVD.

We have explicitly not performed a multiple testing correction for the bidirectional analysis of CRD and CVD with the aim to detect as many putative associations as possible. The only nominally significant association detected was between genetically predicted liability to asthma and an increased risk of AF. Yet, this effect was weak and certainly would not withstand multiple testing corrections. This finding is consistent with a previous study by Chen et al. [18] who also reported diminished genetic associations between asthma and AF after correction. In contrast, another study found stronger evidence for a significant relationship between genetically predicted liability to asthma and AF, even after Bonferroni correction [17]. These conflicting results may potentially be due to differences in GWAS summary statistics they used and IV selection methods.

Current and previous studies on COPD presented conflicting findings depending on which CVD outcome. Contrary to our finding, two separate MR studies found that genetic liability to COPD increases the risk of developing HF [14,16]. However, only one of these studies found evidence of a reverse association, where genetically predicted liability to HF affects COPD [14]. Additionally, the latter study confirmed our findings of no genetic evidence of COPD affecting CAD, IHD, or IS. Once again, one reason that may explain the conflicting results is the use different summary statistics for the outcomes.

Two factors may explain the lack of strong genetic evidence association between CRD and CVD in our study. First, the wide range of phenotypes might obscure real associations, as different phenotypes might vary in heritability affecting disease risk and pathophysiology. We found significant heterogeneity and possibility of pleiotropy in our analyses. By including COPD and asthma as more general diagnoses, we may overlook multiple subtypes that may involve genetic heterogeneity. For example, in a study of former smokers, across four different COPD subtypes, there was some variety in symptoms despite lung function severity [27]. Another example, if onset is considered, a study observed only childhood-onset asthma had genetic evidence to increase CVD risk [28].

Second, there might be no direct pathway for each CRD and CVD pairs. The absence of a direct pathway can suggest that these conditions might instead share a common underlying mechanism, such as smoking, that may explain previous observational findings. Our findings support this hypothesis by showing that genetically predicted lifetime smoking increases the risk of various CVDs as well as COPD and asthma, as reported previously [29–32]. Here, we only included lifetime smoking index as it is itself a comprehensive measure incorporating various factors such as smoking status (current, former, or never), age at initiation and cessation, number of cigarettes smoked per day, smoking duration, and time since cessation [33]. Therefore, we believe it effectively reflects the impact of other smoking characteristics on disease outcomes.

We also provide new insights into the underlying mechanisms on CRD and CVD comorbidity, highlighting the role of several specific cytokine, particularly IL6R, IL6ST, and IL1RN.

We observed that genetically proxied IL6R concentrations were associated with a decreased risk of CVD but an increased risk of asthma. Our findings align with studies showing that higher genetically proxied sIL6R levels are linked to a modestly increased risk of asthma and related phenotypes [34]. Specifically, the IL6R Asp358Ala variant (rs2228145), which increases sIL6R levels, has been reported to be associated with a higher risk of asthma, but not COPD [35]. Conversely, a different SNP in the same Asp358Ala variant has been reported to reduce the risk of coronary heart disease [36]. To complicate our findings, our analysis showed that higher genetically predicted IL6ST concentrations were linked to an increased risk of both AF and asthma.

The complexity of IL6 signalling may explain these differing effects. In classic signalling, IL6 attaches to the membrane bound IL6R (mIL6R) and IL6ST on hepatocytes and immune cells, leading to anti-inflammatory and homeostatic effects. Conversely, in trans-signalling, IL-6 binds to the soluble form of IL6R (sIL6R) and attach to IL6ST on the surface of a broader range of cells that typically do not respond to IL-6, triggering pro-inflammatory responses [37,38]. This dual nature of IL6 signalling might explain the complex distinct direction of inflammatory markers association with different diseases.

Meanwhile, our analysis also associated genetically predicted IL1RN concentration with increased the risk of CAD and MI but also with reduced COPD and asthma risk. The result is consistent with one study linking an IL1RN variant to higher IL1Ra levels, low density lipoprotein and triglycerides, and eventually, increased CVD risk [39]. Only one meta-analysis of observational studies confirmed that polymorphism in IL1RN allele 2 may increase the risk of COPD [40]. The results highlight the importance of understanding inflammation profile, given the mixed reports on anti-inflammatory treatment efficacy in COPD and asthma patients.

Our study has also some strengths. First, MR design offers a unique perspective on the relationship between CRD – CVD comorbidity using genetic evidence minimizing unknown confounding factors. Second, our use of various sensitivity analyses, including the contamination mixture method, ensures robustness under different IV assumptions. By utilizing larger and more reliable GWAS, we gained more statistical power despite using more general terms for diseases. Furthermore, our use of many inflammatory markers from a metanalysis across six studies provided stronger instruments for our analysis.

One limitation of our study is the GWAS included in the MR use broader and more general terms for conditions with multiple and complex phenotypes. Additionally, while we focused only on lifetime smoking to represent smoking, more detailed smoking metrics could provide further insights but were beyond our research scope. Also, we acknowledge the potential for sample overlap due to the inclusion of summary statistics from different studies that included UK Biobank. Still, the risk has been minimized as we excluded weak instruments with F-statistic < 10. Finally, some may consider the misclassification bias risk inherited from the use of external GWAS source. To mitigate this, the present study used COPD summary statistic from a GWAS enhanced by machine learning on spirograms to identify COPD status. Employing machine learning on spirograms may provide a more objective diagnosis than relying solely on patient records.

In conclusion, we did not find genetic evidence to support an association directly between CRD and CVD in either directionality, except for a weakly and nominally significant genetic association between genetic liability to asthma and AF. Our findings confirmed the role of smoking as an established common risk factor contributing to both CRD and CVD. Additionally, we identified several overlapping inflammatory markers that associated with both CRD and CVD. This suggests that the associations we observed in observational and clinical studies may not be due to one disease causing the other. Further work is necessary to extend our findings, to understand better of the overlapping conditions. This includes other epidemiology study designs to study inheritable exposures, such as air pollution, lifestyle, or sociocultural background.

## ETHICS STATEMENT

This study uses summary-level statistics obtained from publicly available GWAS data, for which appropriate ethics approval and participant consent had been previously obtained.

## Data Availability

All data produced in the present study are available upon reasonable request to the authors.

