## Supplement for "Understanding the Bidirectional Relationship Between Chronic Respiratory Disease and Cardiovascular Disease Using Genetic Evidence"

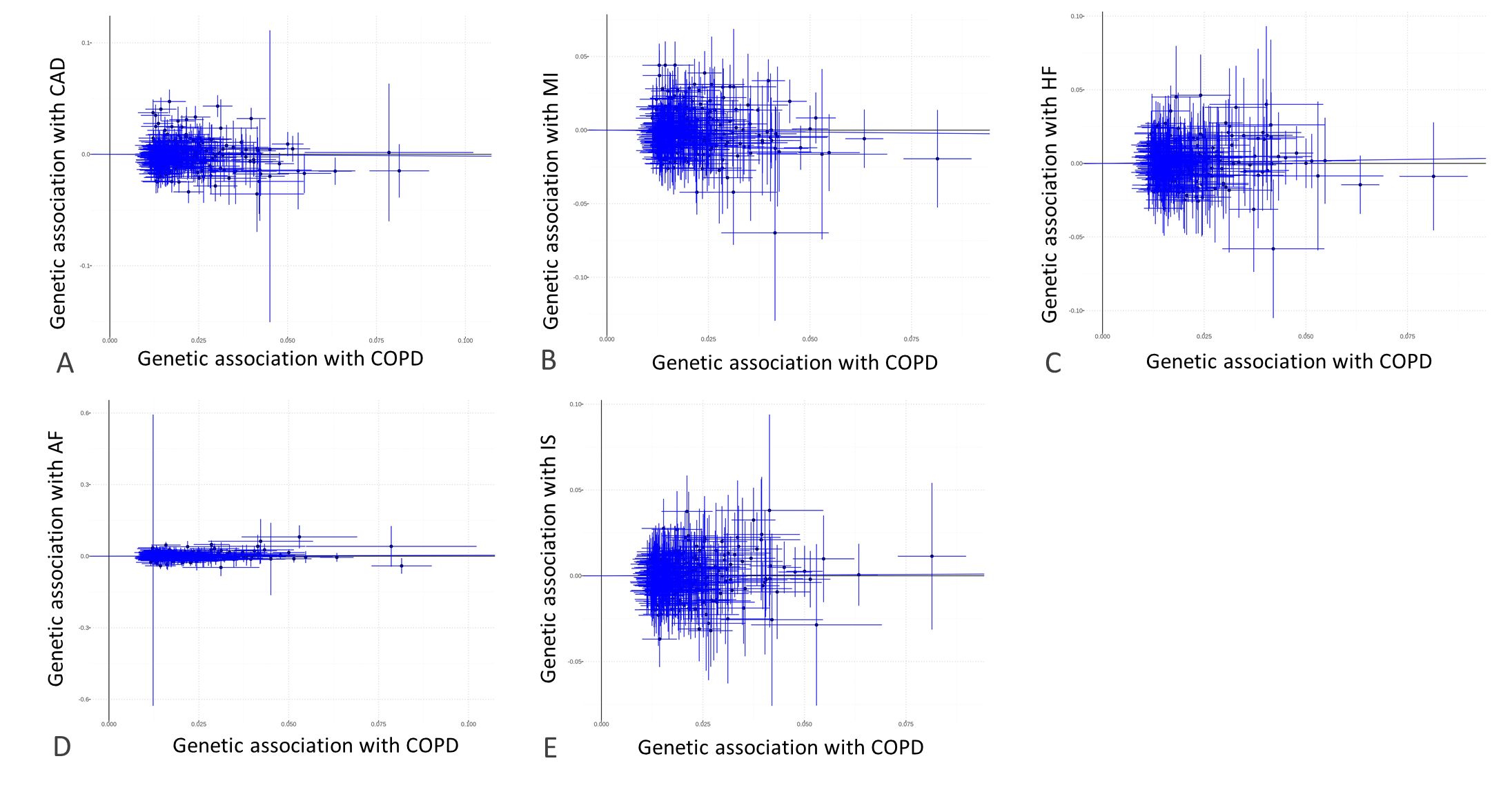

Supplement Figure 1. Scatter plots for MR analyses of the genetic associations between COPD as the exposure (horizontal axis) and (A) CAD, (B) MI (C) HF (D) AF (E) IS as the outcome (vertical axis). Each data point represents a genetic variant used as an instrument variable. Error bars are 95% confidence intervals. The estimate of MR effect from inverse variance weighted (IVW) method is shown as the slope of the regression fit through zero. AF: atrial fibrillation, CAD: coronary artery disease, COPD: chronic obstructive pulmonary diseases, HF: heart failure, IS: ischemia stroke, MI: myocardial infarction

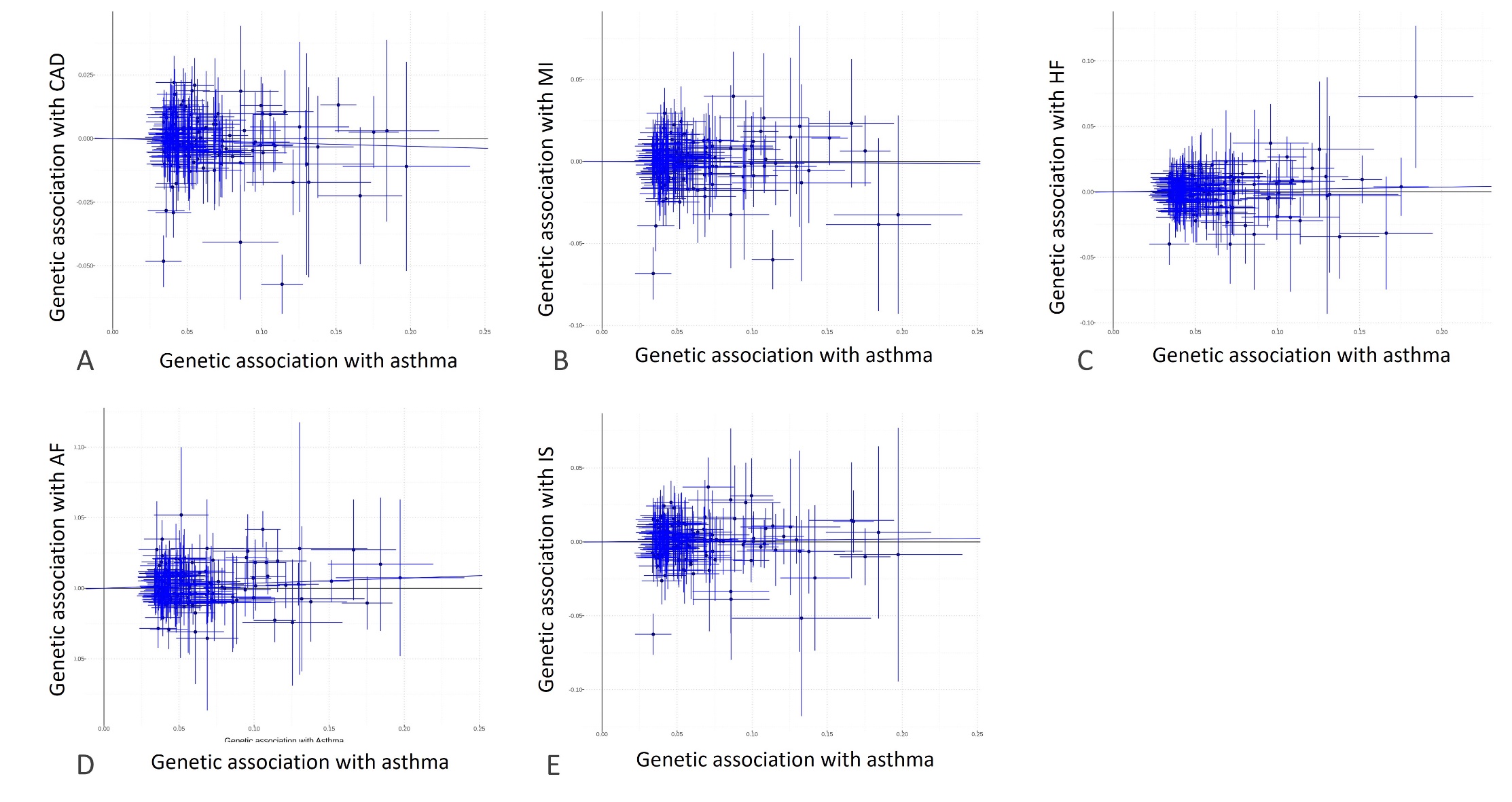

Supplement Figure 2. Scatter plots for MR analyses of the genetic associations between asthma as the exposure (horizontal axis) and (A) CAD (B) MI (C) HF (D) AF (E) IS as the outcome (vertical axis). Each data point represents a genetic variant used as an instrument variable. Error bars are 95% confidence intervals. The estimate of MR effect from inverse variance weighted (IVW) method is shown as the slope of the regression fit through zero. AF: atrial fibrillation CAD: coronary artery disease, HF: heart failure, IS: ischemia stroke, MI: myocardial infarction.

.

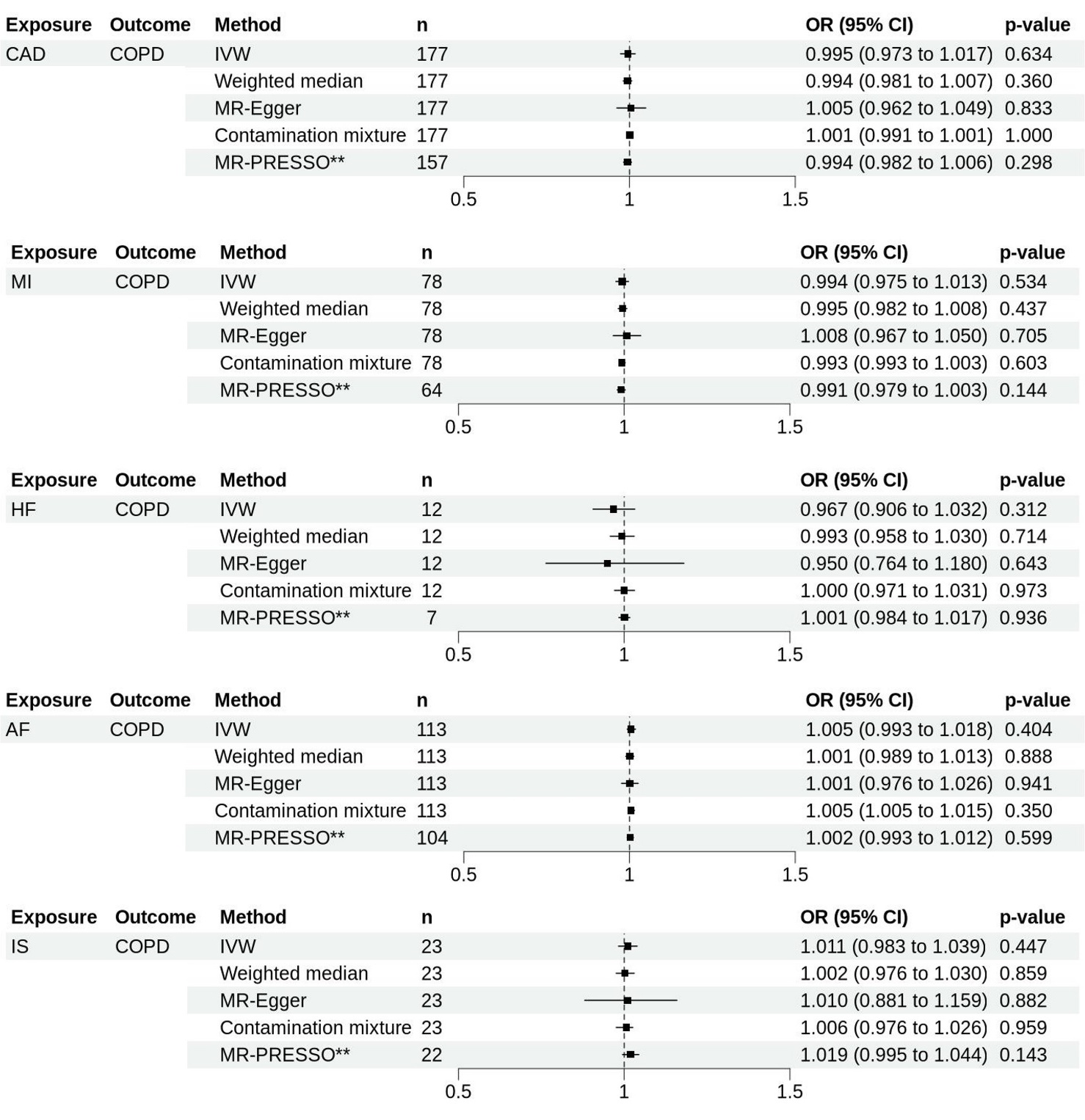

Supplement Figure 3. Forest plots showing the results of MR analyses on the effect of genetically predicted liability to cardiovascular diseases on the risk of developing COPD. Estimates are shown as OR. Horizontal lines represent the 95% CIs. AF: atrial fibrillation, CAD: coronary artery disease, COPD: chronic obstructive pulmonary disease, HF: heart failure, IS: ischemia stroke, MI: myocardial infarction, n: number of SNPs used as instrument variables in each method, SNPs single nucleotide polymorphism, CI confidence intervals, OR odds ratio. ** indicates corrected MR-PRESSO estimates

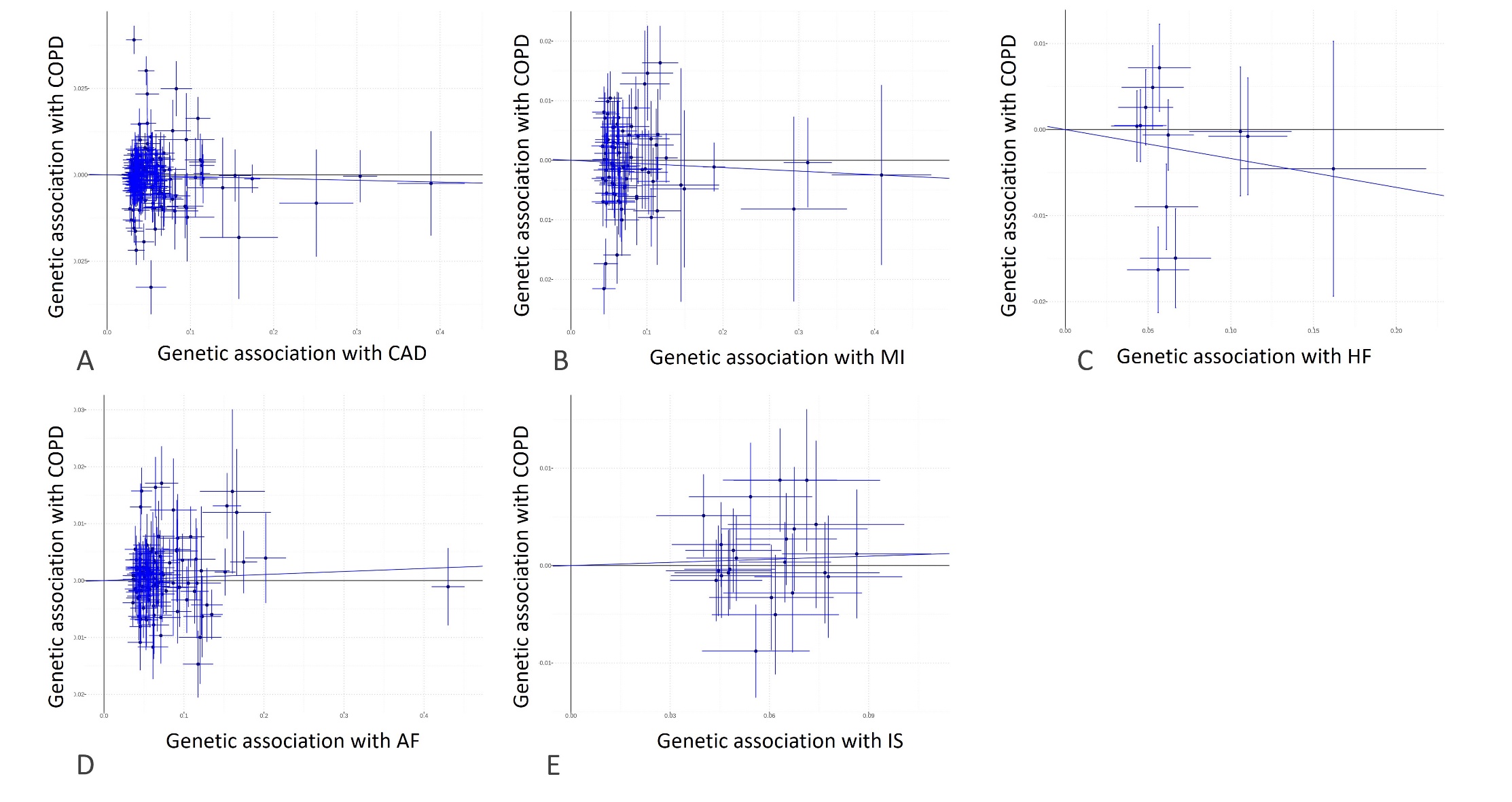

Supplement Figure 4. Scatter plots for MR analyses of the genetic associations between (A) CAD (B) MI (C) HF (D) AF (E) IS as the exposures (horizontal axis) towards COPD as the outcome (vertical axis). Each data point represents a genetic variant used as an instrument variable. Error bars are 95% confidence intervals. The estimate of MR effect from inverse variance weighted (IVW) method is shown as the slope of the regression fit through zero. AF: atrial fibrillation CAD: coronary artery disease, COPD: chronic obstructive pulmonary disease, HF: heart failure, IS: ischemia stroke, MI: myocardial infarction.

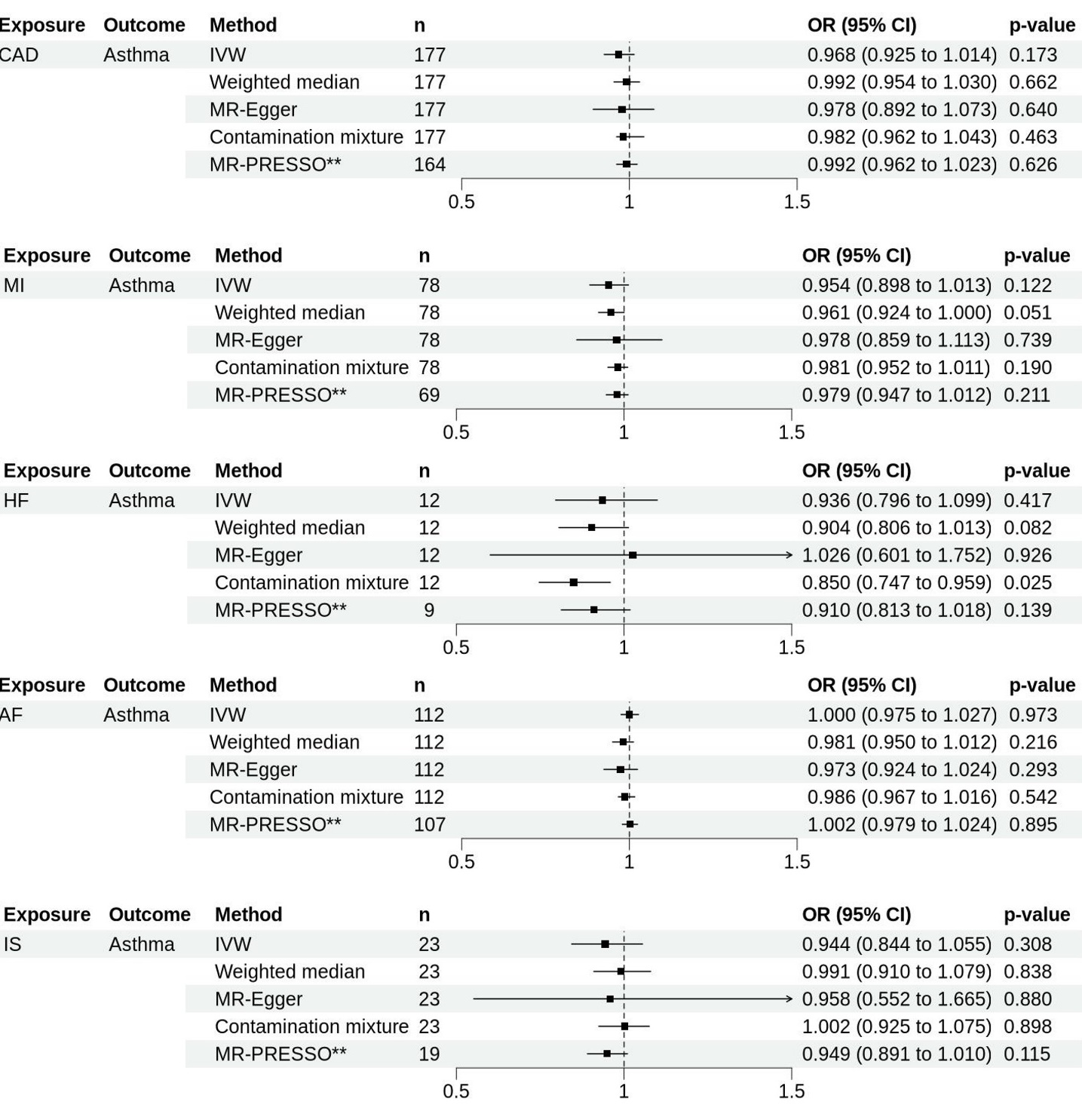

Supplement Figure 5. Forest plots showing the results of MR analyses on the effect of genetically predicted liability to cardiovascular diseases on the risk of asthma. Estimates are shown as OR. Horizontal lines represent the 95% CIs. AF: atrial fibrillation, CAD: coronary artery disease, HF: heart failure, IS: ischemia stroke, MI: myocardial infarction, n: number of SNPs used as instrument variables in each method, SNPs single nucleotide polymorphism, CI confidence intervals, OR odds ratio. ** indicates corrected MR-PRESSO estimates

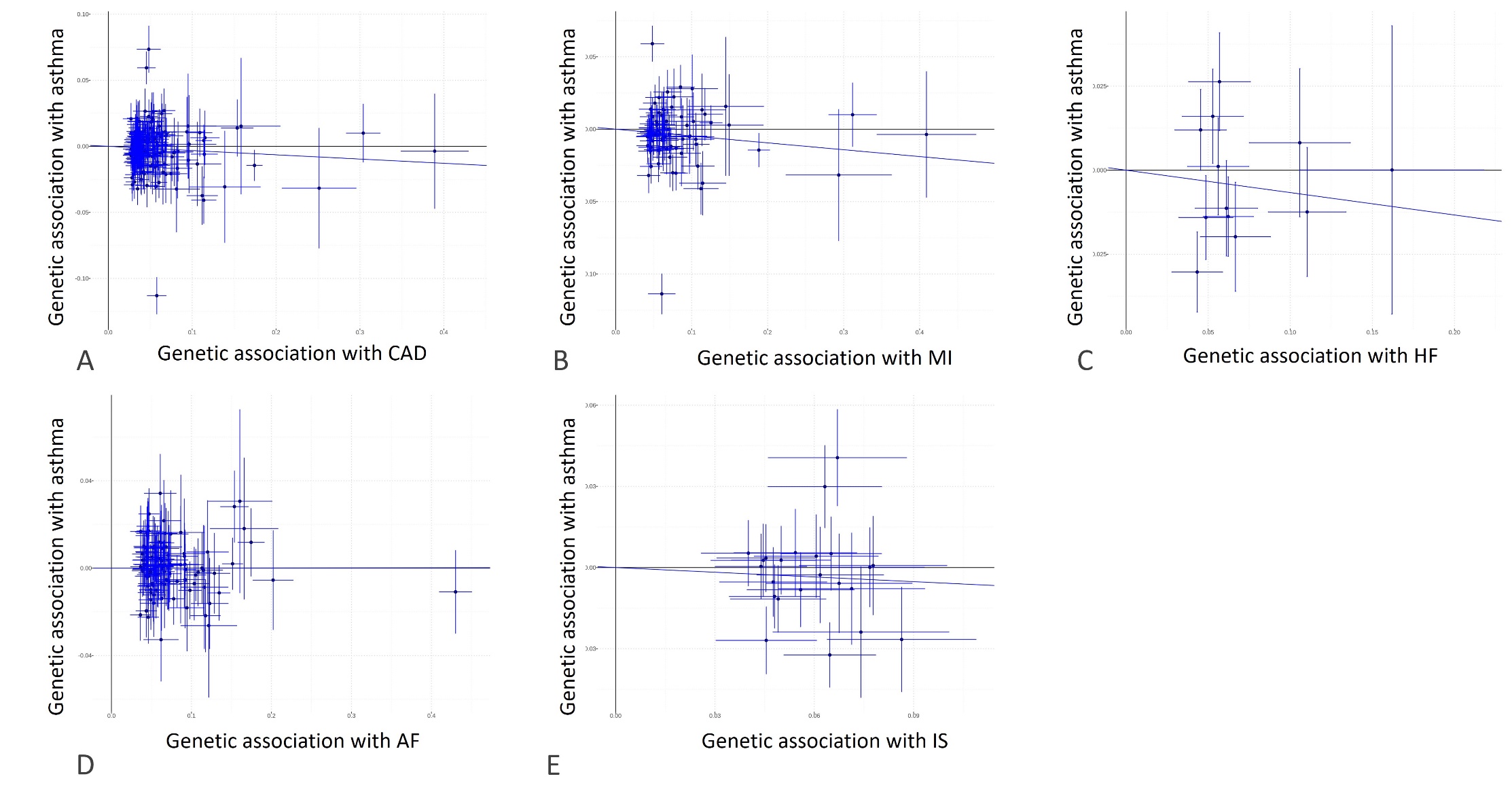

Supplement Figure 6. Scatter plots for MR analyses of the genetic associations between (A) CAD (B) MI (C) HF (D) AF (E) IS as the exposures towards asthma as the outcome (vertical axis). Each data point represents a genetic variant used as an instrument variable. Error bars are 95% confidence intervals. The estimate of MR effect from inverse variance weighted (IVW) method is shown as the slope of the regression fit through zero. AF: atrial fibrillation, CAD: coronary artery disease, HF: heart failure, IS: ischemia stroke, MI: myocardial infarction

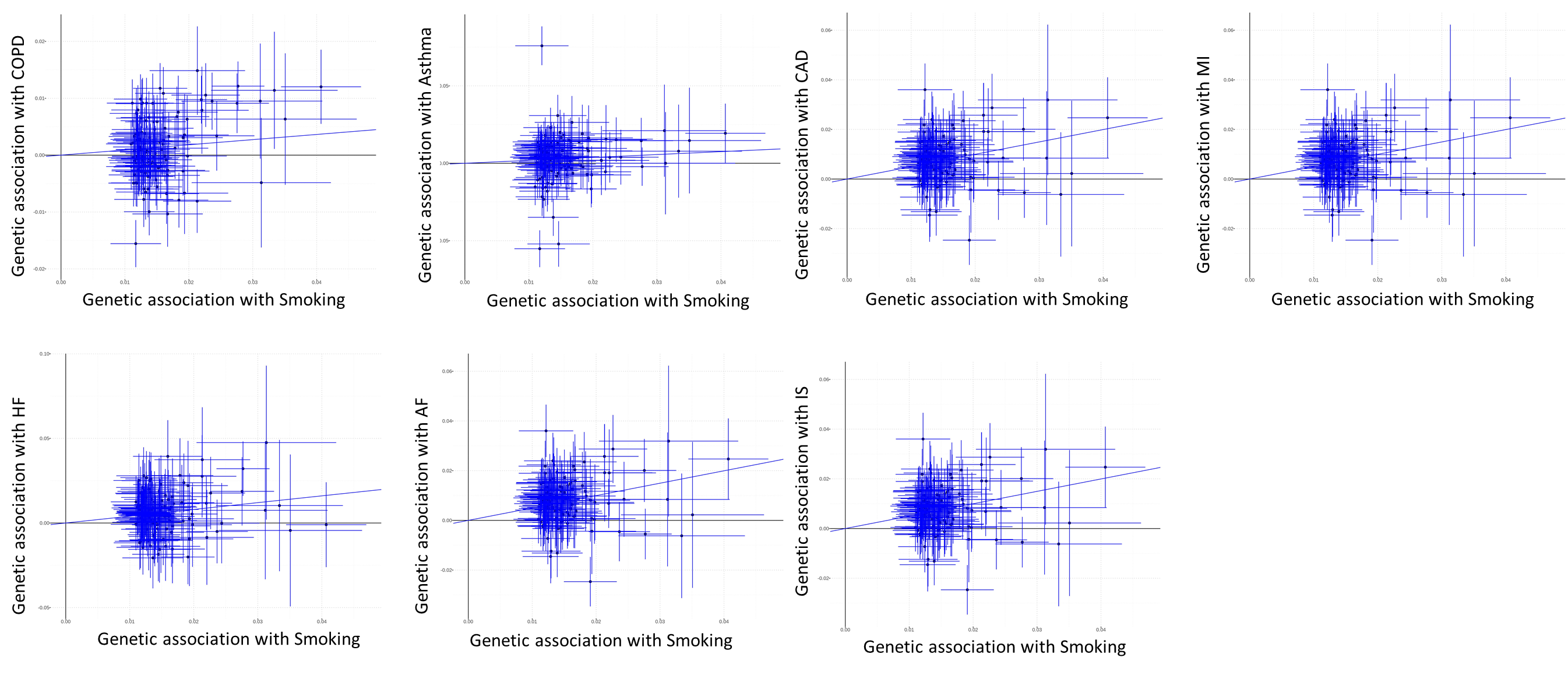

Supplement Figure 7. Scatter plots for MR analyses of the genetic associations between lifetime smoking as the exposure (horizontal axis) and CRD and CVD as the outcome (vertical axis). Each data point represents a genetic variant used as an instrument variable. Error bars are 95% confidence intervals. The estimate of MR effect from inverse variance weighted (IVW) method is shown as the slope of the regression fit through zero. AF: atrial fibrillation, CAD: coronary artery disease, COPD: chronic obstructive pulmonary disease, HF: heart failure, IS: ischemia stroke, MI: myocardial infarction

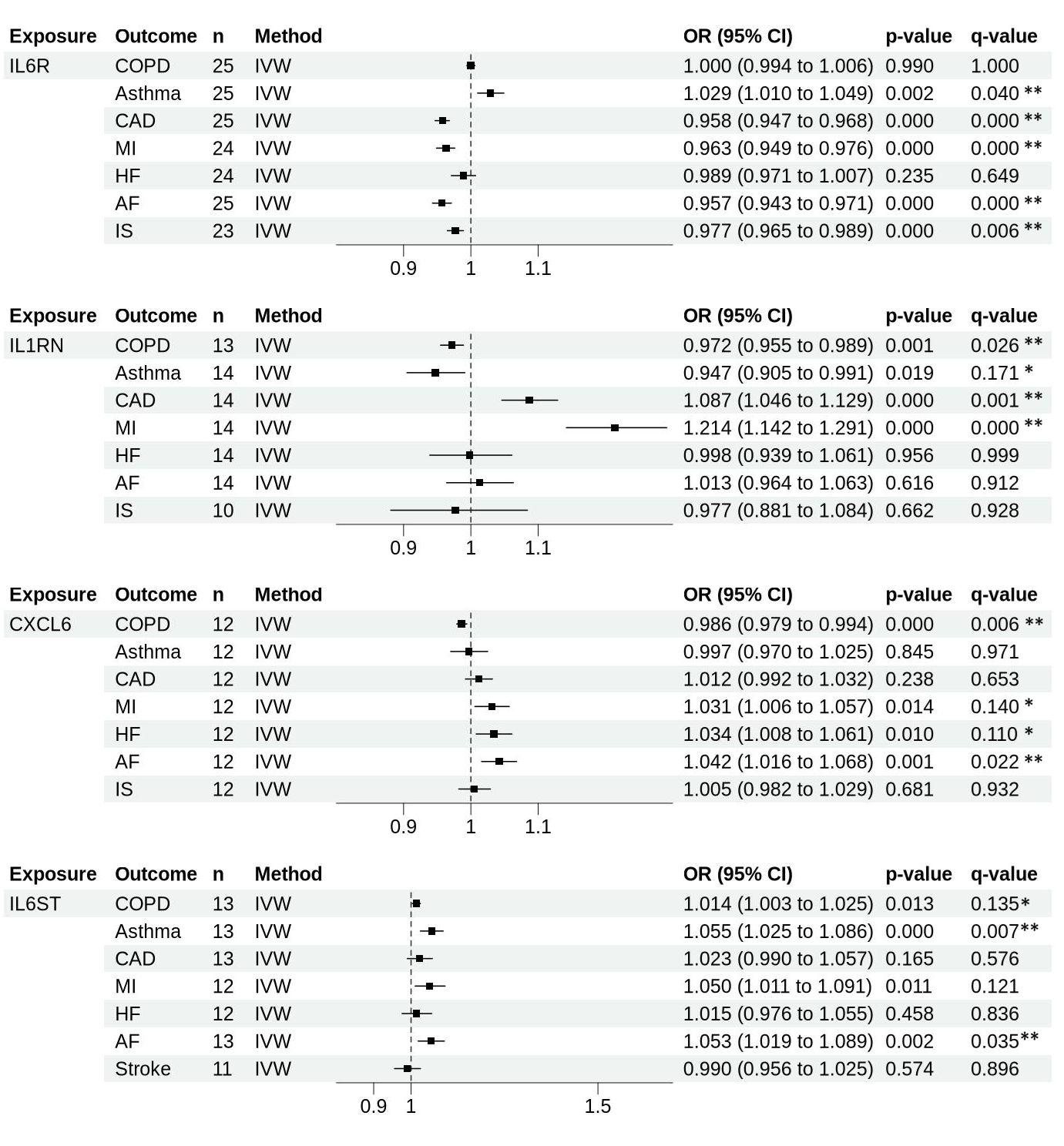

Supplement Figure 8. Forest plot showing the result of cis-MR analysis on the effect of genetically predicted inflammatory markers associations found to be overlapped in both CRD and CVD. * denotes suggestive evidence (0.05 < q-value ≤ 0.2) ** denotes strong evidence (q-value <0.05). Horizontal lines represent the 95% CIs. AF: atrial fibrillation, CAD: coronary artery disease, COPD: chronic obstructive pulmonary disease, HF: heart failure, IL: interleukin, IL6R: IL6 Receptor, IL6ST: IL6 Cytokine Family Signal Transducer, IL1RN: IL1 receptor antagonist, IL1RL2: IL1 receptor-like 2, IL1R2: IL1 Receptor Type 2, IS: ischemia stroke, MI: myocardial infarction, n: number of SNPs used as instrument variables in each method, SNPs single nucleotide polymorphism, CI confidence intervals, OR odds ratio

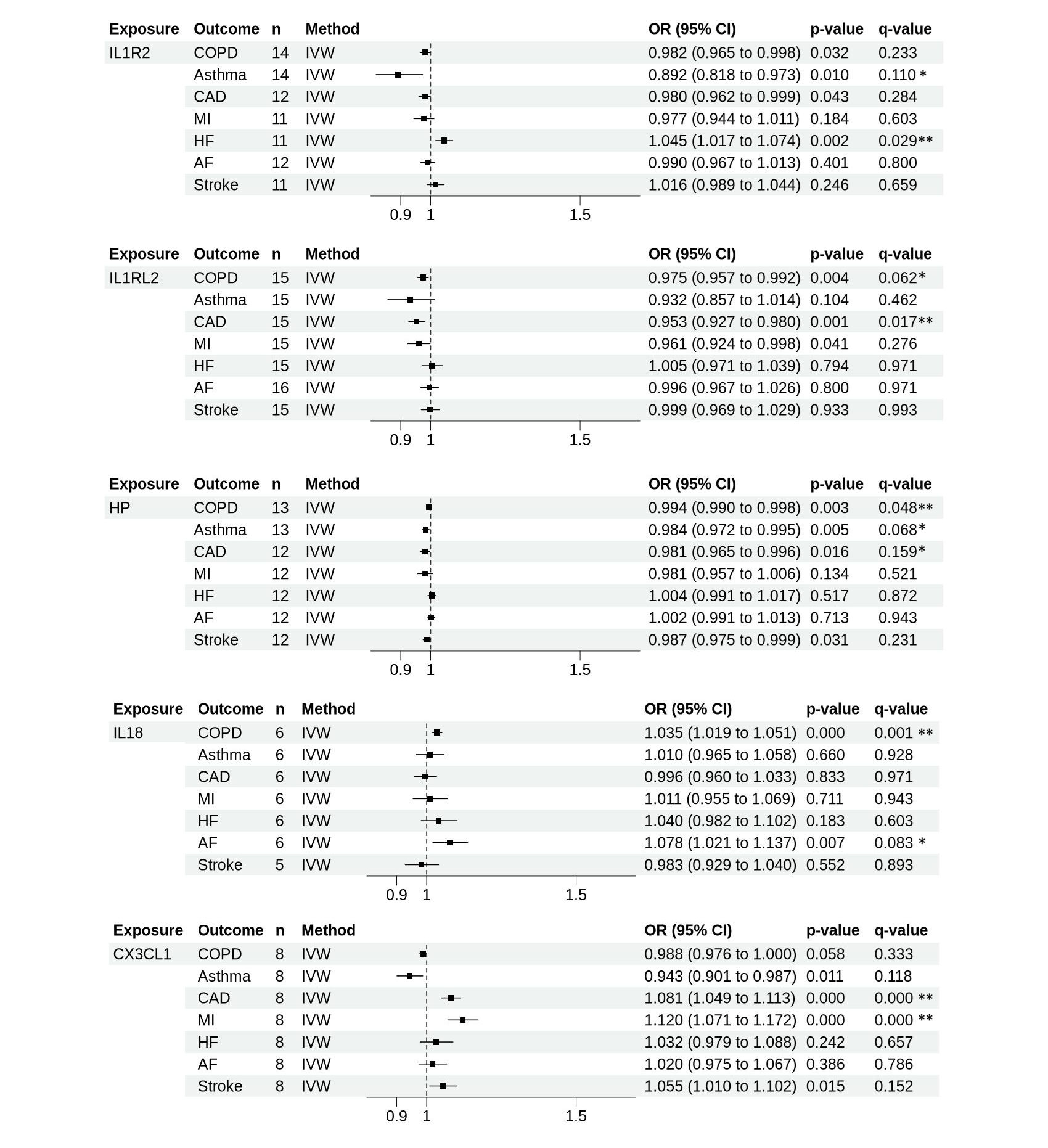
Supplement Figure 8. cont. Forest plot showing the result of cis-MR analysis on the effect of genetically predicted inflammatory markers associations found to be overlapped in both CRD and CVD. * denotes suggestive evidence (0.05 < q-value ≤ 0.2) ** denotes strong evidence (q-value <0.05). Horizontal lines represent the 95% CIs. AF: atrial fibrillation, CAD: coronary artery disease, COPD: chronic obstructive pulmonary disease, HF: heart failure, IL: interleukin, IL6R: IL6 Receptor, IL6ST: IL6 Cytokine Family Signal Transducer, IL1RN: IL1 receptor antagonist, IL1RL2: IL1 receptor-like 2, IL1R2: IL1 Receptor Type 2, IS: ischemia stroke, MI: myocardial infarction, n: number of SNPs used as instrument variables in each method, SNPs single nucleotide polymorphism, CI confidence intervals, OR odds ratio

**Supplement Table 1. Mendelian Randomization analysis result for the effect of chronic obstructive pulmonary disease on cardiovascular diseases**

| **Exposure** | **Outcome** | **Methods** | **n** | **Estimate** | **betalow** | **betahi** | **OR** | **ORlow** | **ORhi** | **p-value** | **Heterogeneity test statistic (Cochran's Q)** | **df** | **I**^2^ | **F-statistic** | **P-het value** | **P**_egger_intercept_ |
| --- | --- | --- | --- | --- | --- | --- | --- | --- | --- | --- | --- | --- | --- | --- | --- | --- |
| COPD | CAD | IVW | 302 | -0.016 | -0.082 | 0.051 | 0.984 | 0.921 | 1.052 | 0.644 | 1283.238 | 301 | 76.5% | 72.7 | 0.000 |  |
|  |  | Weighted median | 302 | -0.019 | -0.078 | 0.041 | 0.981 | 0.925 | 1.042 | 0.536 |  |  |  |  |  |  |
|  |  | MR-Egger | 302 | 0.040 | -0.134 | 0.214 | 1.041 | 0.874 | 1.239 | 0.653 |  |  |  |  |  | 0.498 |
|  |  | Contamination mixture | 302 | -0.076 | -0.126 | 0.014 | 0.927 | 0.882 | 1.014 | 0.138 |  |  |  |  |  |  |
|  |  | MR-PRESSO | 279 | -0.070 | -0.117 | -0.023 | 0.932 | 0.889 | 0.977 | 0.004 |  |  |  |  |  |  |
| COPD | MI | IVW | 298 | -0.026 | -0.103 | 0.051 | 0.974 | 0.902 | 1.052 | 0.506 | 744.5619 | 297 | 60.1% | 71.8 | 0.000 |  |
|  |  | Weighted median | 298 | -0.092 | -0.177 | -0.007 | 0.912 | 0.838 | 0.993 | 0.034 |  |  |  |  |  |  |
|  |  | MR-Egger | 298 | 0.014 | -0.187 | 0.216 | 1.014 | 0.829 | 1.241 | 0.890 |  |  |  |  |  | 0.672 |
|  |  | Contamination mixture | 298 | -0.159 | -0.249 | -0.089 | 0.853 | 0.780 | 0.915 | 0.000 |  |  |  |  |  |  |
|  |  | MR-PRESSO | 287 | -0.082 | -0.148 | -0.016 | 0.921 | 0.862 | 0.984 | 0.015 |  |  |  |  |  |  |
| COPD | HF | IVW | 297 | 0.036 | -0.026 | 0.098 | 1.037 | 0.974 | 1.103 | 0.256 | 413.055 | 296 | 28.3% | 71.5 | 0.000 |  |
|  |  | Weighted median | 297 | 0.032 | -0.053 | 0.117 | 1.032 | 0.949 | 1.124 | 0.461 |  |  |  |  |  |  |
|  |  | MR-Egger | 297 | 0.045 | -0.118 | 0.209 | 1.046 | 0.889 | 1.232 | 0.585 |  |  |  |  |  | 0.901 |
|  |  | Contamination mixture | 297 | 0.035 | -0.045 | 0.135 | 1.036 | 0.956 | 1.145 | 0.371 |  |  |  |  |  |  |
|  |  | MR-PRESSO | 296 | 0.027 | -0.035 | 0.088 | 1.027 | 0.966 | 1.092 | 0.398 |  |  |  |  |  |  |
| COPD | AF | IVW | 303 | 0.038 | -0.032 | 0.108 | 1.039 | 0.968 | 1.114 | 0.288 | 751.299 | 302 | 59.8% | 72.4 | 0.000 |  |
|  |  | Weighted median | 303 | -0.013 | -0.091 | 0.065 | 0.987 | 0.913 | 1.067 | 0.743 |  |  |  |  |  |  |
|  |  | MR-Egger | 303 | 0.006 | -0.178 | 0.189 | 1.006 | 0.837 | 1.208 | 0.952 |  |  |  |  |  | 0.711 |
|  |  | Contamination mixture | 303 | 0.014 | -0.076 | 0.084 | 1.014 | 0.927 | 1.087 | 0.693 |  |  |  |  |  |  |
|  |  | MR-PRESSO | 292 | 0.018 | -0.042 | 0.077 | 1.018 | 0.959 | 1.080 | 0.561 |  |  |  |  |  |  |
| COPD | IS | IVW | 294 | 0.011 | -0.047 | 0.068 | 1.011 | 0.954 | 1.070 | 0.717 | 439.141 | 293 | 33.3% | 71.8 | 0.000 |  |
|  |  | Weighted median | 294 | 0.042 | -0.036 | 0.120 | 1.043 | 0.965 | 1.128 | 0.289 |  |  |  |  |  |  |
|  |  | MR-Egger | 294 | 0.130 | -0.022 | 0.281 | 1.138 | 0.978 | 1.325 | 0.094 |  |  |  |  |  | 0.097 |
|  |  | Contamination mixture | 294 | 0.015 | -0.075 | 0.095 | 1.015 | 0.928 | 1.100 | 0.540 |  |  |  |  |  |  |
|  |  | MR-PRESSO | 292 | 0.011 | -0.044 | 0.067 | 1.011 | 0.957 | 1.069 | 0.695 |  |  |  |  |  |  |

betalow: 95% lower CI for the estimate, betahi: 95% upper CI for the estimate.

ORlow: 95% lower CI for the OR, ORhi: 95% upper CI for the OR.

AF: atrial fibrillation, CAD: coronary artery disease, COPD: chronic obstructive pulmonary diseases, HF: heart failure, IS: ischemia stroke, MI: myocardial infarction, n: number of SNPs used as instrument variables in each method, SNPs single nucleotide polymorphism, CI confidence intervals, OR odds ratio.

** indicates corrected MR-PRESSO estimates
**Supplement Table 2. Mendelian Randomization analysis result for the effect of asthma on cardiovascular disease**

| **Exposure** | **Outcome** | **Methods** | **n** | **Estimate** | **betalow** | **betahi** | **OR** | **ORlow** | **ORhi** | **p-value** | **Heterogeneity test statistic (Cochran's Q)** | **df** | **I**^2^ | **F-statistic** | **P-het value** | **P**_egger_intercept_ |
| --- | --- | --- | --- | --- | --- | --- | --- | --- | --- | --- | --- | --- | --- | --- | --- | --- |
| Asthma | CAD | IVW | 146 | -0.015 | -0.045 | 0.014 | 0.985 | 0.956 | 1.014 | 0.311 | 497.874 | 145 | 70.9% | 74.4 | 0.000 |  |
|  |  | Weighted median | 146 | -0.011 | -0.038 | 0.017 | 0.989 | 0.963 | 1.017 | 0.445 |  |  |  |  |  |  |
|  |  | MR-Egger | 146 | -0.005 | -0.076 | 0.066 | 0.995 | 0.927 | 1.068 | 0.889 |  |  |  |  |  | 0.756 |
|  |  | Contamination mixture | 146 | -0.016 | -0.036 | 0.004 | 0.984 | 0.965 | 1.004 | 0.207 |  |  |  |  |  |  |
|  |  | MR-PRESSO | 138 | 0.001 | -0.019 | 0.021 | 1.001 | 0.981 | 1.021 | 0.908 |  |  |  |  |  |  |
| Asthma | MI | IVW | 144 | -0.005 | -0.043 | 0.033 | 0.995 | 0.958 | 1.033 | 0.791 | 346.010 | 143 | 58.7% | 72.8 | 0.000 |  |
|  |  | Weighted median | 144 | 0.011 | -0.030 | 0.051 | 1.011 | 0.971 | 1.053 | 0.599 |  |  |  |  |  |  |
|  |  | MR-Egger | 144 | 0.038 | -0.053 | 0.129 | 1.039 | 0.948 | 1.138 | 0.416 |  |  |  |  |  | 0.310 |
|  |  | Contamination mixture | 144 | 0.022 | -0.008 | 0.052 | 1.022 | 0.992 | 1.053 | 0.235 |  |  |  |  |  |  |
|  |  | MR-PRESSO | 140 | 0.014 | -0.016 | 0.043 | 1.014 | 0.985 | 1.044 | 0.360 |  |  |  |  |  |  |
| Asthma | HF | IVW | 144 | 0.018 | -0.013 | 0.050 | 1.019 | 0.987 | 1.051 | 0.254 | 220.246 | 143 | 35.1% | 74.7 | 0.000 |  |
|  |  | Weighted median | 144 | 0.062 | 0.020 | 0.105 | 1.064 | 1.021 | 1.110 | 0.004 |  |  |  |  |  |  |
|  |  | MR-Egger | 144 | 0.073 | -0.003 | 0.149 | 1.076 | 0.997 | 1.160 | 0.060 |  |  |  |  |  | 0.122 |
|  |  | Contamination mixture | 144 | 0.070 | 0.030 | 0.100 | 1.072 | 1.030 | 1.105 | 0.001 |  |  |  |  |  |  |
|  |  | MR-PRESSO | 143 | 0.022 | -0.008 | 0.052 | 1.022 | 0.992 | 1.053 | 0.153 |  |  |  |  |  |  |
| Asthma | AF | IVW | 146 | 0.035 | 0.003 | 0.068 | 1.036 | 1.003 | 1.070 | 0.030 | 317.592 | 145 | 54.3% | 74.3 | 0.000 |  |
|  |  | Weighted median | 146 | 0.025 | -0.013 | 0.064 | 1.026 | 0.987 | 1.066 | 0.193 |  |  |  |  |  |  |
|  |  | MR-Egger | 146 | 0.051 | -0.026 | 0.128 | 1.052 | 0.975 | 1.136 | 0.192 |  |  |  |  |  | 0.661 |
|  |  | Contamination mixture | 146 | 0.005 | -0.025 | 0.055 | 1.005 | 0.976 | 1.057 | 0.603 |  |  |  |  |  |  |
|  |  | MR-PRESSO | 140 | 0.035 | 0.008 | 0.062 | 1.036 | 1.008 | 1.064 | 0.012 |  |  |  |  |  |  |
| Asthma | IS | IVW | 142 | 0.009 | -0.025 | 0.043 | 1.009 | 0.976 | 1.044 | 0.586 | 295.668 | 141 | 52.3% | 75.8 | 0.000 |  |
|  |  | Weighted median | 142 | 0.008 | -0.029 | 0.045 | 1.008 | 0.971 | 1.046 | 0.668 |  |  |  |  |  |  |
|  |  | MR-Egger | 142 | 0.012 | -0.069 | 0.094 | 1.012 | 0.933 | 1.098 | 0.768 |  |  |  |  |  | 0.940 |
|  |  | Contamination mixture | 142 | 0.013 | -0.017 | 0.043 | 1.014 | 0.984 | 1.044 | 0.449 |  |  |  |  |  |  |
|  |  | MR-PRESSO | 141 | 0.016 | -0.014 | 0.045 | 1.016 | 0.986 | 1.046 | 0.299 |  |  |  |  |  |  |

betalow: 95% lower CI for the estimate; betahi: 95% upper CI for the estimate.

ORlow: 95% lower CI for the OR; ORhi: 95% upper CI for the OR.

AF: atrial fibrillation, CAD: coronary artery disease, HF: heart failure, IS: ischemia stroke, MI: myocardial infarction, n: number of SNPs used as instrument variables in each method, SNPs single nucleotide polymorphism, CI confidence intervals, OR odds ratio.

** indicates corrected MR-PRESSO estimates

**Supplement Table 3. Mendelian Randomization analysis result for the effect of cardiovascular disease on chronic obstructive pulmonary disease**

| **Exposure** | **Outcome** | **Method** | **n** | **Estimate** | **betalow** | **betahi** | **OR** | **ORlow** | **ORhi** | **p-value** | **Heterogeneity test statistic (Cochran's Q)** | **df** | **I**^2^ | **F-statistic** | **P-het value** | | **P**_egger_intercept_ |
| --- | --- | --- | --- | --- | --- | --- | --- | --- | --- | --- | --- | --- | --- | --- | --- | --- | --- |
| CAD | COPD | IVW | 177 | -0.005 | -0.027 | 0.016 | 0.995 | 0.973 | 1.017 | 0.634 | 1704.448 | 176 | 89.70% | 76.9 | 0 | |  |
|  |  | Weighted median | 177 | -0.006 | -0.020 | 0.007 | 0.994 | 0.981 | 1.007 | 0.360 |  |  |  |  |  | |  |
|  |  | MR-Egger | 177 | 0.005 | -0.038 | 0.048 | 1.005 | 0.962 | 1.049 | 0.833 |  |  |  |  |  | | 0.602 |
|  |  | Contamination mixture | 177 | 0.001 | -0.009 | 0.001 | 1.001 | 0.991 | 1.001 | 1.000 |  |  |  |  |  | |  |
|  |  | MR-PRESSO** | 157 | -0.006 | -0.018 | 0.006 | 0.994 | 0.982 | 1.006 | 0.298 |  |  |  |  |  | |  |
| MI | COPD | IVW | 78 | -0.006 | -0.025 | 0.013 | 0.994 | 0.975 | 1.013 | 0.534 | 512.379 | 77 | 85.00% | 65.2 | 0 | |  |
|  |  | Weighted median | 78 | -0.005 | -0.018 | 0.008 | 0.995 | 0.982 | 1.008 | 0.437 |  |  |  |  |  | |  |
|  |  | MR-Egger | 78 | 0.008 | -0.033 | 0.049 | 1.008 | 0.967 | 1.050 | 0.705 |  |  |  |  |  | | 0.449 |
|  |  | Contamination mixture | 78 | -0.007 | -0.007 | 0.003 | 0.993 | 0.993 | 1.003 | 0.603 |  |  |  |  |  | |  |
|  |  | MR-PRESSO** | 64 | -0.009 | -0.021 | 0.003 | 0.991 | 0.979 | 1.003 | 0.144 |  |  |  |  |  | |  |
| HF | COPD | IVW | 12 | -0.034 | -0.099 | 0.032 | 0.967 | 0.906 | 1.032 | 0.312 | 85.09 | 11 | 87.10% | 41.5 | 0 | |  |
|  |  | Weighted median | 12 | -0.007 | -0.043 | 0.029 | 0.993 | 0.958 | 1.030 | 0.714 |  |  |  |  |  | |  |
|  |  | MR-Egger | 12 | -0.051 | -0.269 | 0.166 | 0.950 | 0.764 | 1.180 | 0.643 |  |  |  |  |  | | 0.866 |
|  |  | Contamination mixture | 12 | 0.000 | -0.030 | 0.030 | 1.000 | 0.971 | 1.031 | 0.973 |  |  |  |  |  | |  |
|  |  | MR-PRESSO** | 7 | 0.001 | -0.016 | 0.017 | 1.001 | 0.984 | 1.017 | 0.936 |  |  |  |  |  | |  |
| AF | COPD | IVW | 113 | 0.005 | -0.007 | 0.018 | 1.005 | 0.993 | 1.018 | 0.404 | 458.647 | 112 | 75.60% | 89.6 | 0 | |  |
|  |  | Weighted median | 113 | 0.001 | -0.011 | 0.012 | 1.001 | 0.989 | 1.013 | 0.888 |  |  |  |  |  | |  |
|  |  | MR-Egger | 113 | 0.001 | -0.024 | 0.026 | 1.001 | 0.976 | 1.026 | 0.941 |  |  |  |  |  | | 0.693 |
|  |  | Contamination mixture | 113 | 0.005 | 0.005 | 0.015 | 1.005 | 1.005 | 1.015 | 0.350 |  |  |  |  |  | |  |
|  |  | MR-PRESSO** | 104 | 0.002 | -0.007 | 0.012 | 1.002 | 0.993 | 1.012 | 0.599 |  |  |  |  |  | |  |
| IS | COPD | IVW | 23 | 0.011 | -0.017 | 0.038 | 1.011 | 0.983 | 1.039 | 0.447 | 51.077 | 22 | 56.90% | 44.7 | 0 | |  |
|  |  | Weighted median | 23 | 0.002 | -0.025 | 0.030 | 1.002 | 0.976 | 1.030 | 0.859 |  |  |  |  |  | |  |
|  |  | MR-Egger | 23 | 0.010 | -0.127 | 0.148 | 1.010 | 0.881 | 1.159 | 0.882 |  |  |  |  |  | | 0.996 |
|  |  | Contamination mixture | 23 | 0.006 | -0.024 | 0.026 | 1.006 | 0.976 | 1.026 | 0.959 |  |  |  |  |  | |  |
|  |  | MR-PRESSO** | 22 | 0.019 | -0.005 | 0.043 | 1.019 | 0.995 | 1.044 | 0.143 |  |  |  |  | |  | |

betalow: 95% lower CI for the estimate; betahi: 95% upper CI for the estimate.

ORlow: 95% lower CI for the OR; ORhi: 95% upper CI for the OR.

AF: atrial fibrillation, CAD: coronary artery disease, COPD: chronic obstructive pulmonary diseases, HF: heart failure, IS: ischemia stroke, MI: myocardial infarction, n: number of SNPs used as instrument variables in each method, SNPs single nucleotide polymorphism, CI confidence intervals, OR odds ratio.

** indicates corrected MR-PRESSO estimates

**Supplement Table 4. Mendelian Randomization analysis result for the effect of cardiovascular disease on asthma**

| **Exposure** | **Outcome** | **Methods** | **n** | **Estimate** | **betalow** | **betahi** | **OR** | **ORlow** | **ORhi** | **p-value** | **Heterogeneity test statistic (Cochran's Q)** | **df** | **I**^2^ | **F-statistic** | **P-het value** | **P**_egger_intercept_ |
| --- | --- | --- | --- | --- | --- | --- | --- | --- | --- | --- | --- | --- | --- | --- | --- | --- |
| CAD | Asthma | IVW | 177 | -0.032 | -0.078 | 0.014 | 0.968 | 0.925 | 1.014 | 0.173 | 934.867 | 176 | 81.2% | 76.9 | 0.000 |  |
|  |  | Weighted median | 177 | -0.008 | -0.047 | 0.030 | 0.992 | 0.954 | 1.030 | 0.662 |  |  |  |  |  |  |
|  |  | MR-Egger | 177 | -0.022 | -0.114 | 0.070 | 0.978 | 0.892 | 1.073 | 0.640 |  |  |  |  |  | 0.805 |
|  |  | Contamination mixture | 177 | -0.018 | -0.038 | 0.042 | 0.982 | 0.962 | 1.043 | 0.463 |  |  |  |  |  |  |
|  |  | MR-PRESSO** | 164 | -0.008 | -0.038 | 0.023 | 0.992 | 0.962 | 1.023 | 0.626 |  |  |  |  |  |  |
| MI | Asthma | IVW | 78 | -0.047 | -0.107 | 0.013 | 0.954 | 0.898 | 1.013 | 0.122 | 606.138 | 77 | 87.3% | 65.2 | 0.000 |  |
|  |  | Weighted median | 78 | -0.039 | -0.079 | 0.000 | 0.961 | 0.924 | 1.000 | 0.051 |  |  |  |  |  |  |
|  |  | MR-Egger | 78 | -0.022 | -0.151 | 0.107 | 0.978 | 0.859 | 1.113 | 0.739 |  |  |  |  |  | 0.665 |
|  |  | Contamination mixture | 78 | -0.019 | -0.049 | 0.011 | 0.981 | 0.952 | 1.011 | 0.190 |  |  |  |  |  |  |
|  |  | MR-PRESSO** | 69 | -0.021 | -0.054 | 0.012 | 0.979 | 0.947 | 1.012 | 0.211 |  |  |  |  |  |  |
| HF | Asthma | IVW | 12 | -0.067 | -0.228 | 0.094 | 0.936 | 0.796 | 1.099 | 0.417 | 62.767 | 11 | 82.5% | 41.5 | 0.000 |  |
|  |  | Weighted median | 12 | -0.101 | -0.216 | 0.013 | 0.904 | 0.806 | 1.013 | 0.082 |  |  |  |  |  |  |
|  |  | MR-Egger | 12 | 0.025 | -0.510 | 0.561 | 1.026 | 0.601 | 1.752 | 0.926 |  |  |  |  |  | 0.722 |
|  |  | Contamination mixture | 12 | -0.162 | -0.292 | -0.042 | 0.850 | 0.747 | 0.959 | 0.025 |  |  |  |  |  |  |
|  |  | MR-PRESSO** | 9 | -0.095 | -0.208 | 0.018 | 0.910 | 0.813 | 1.018 | 0.139 |  |  |  |  |  |  |
| AF | Asthma | IVW | 112 | 0.000 | -0.025 | 0.026 | 1.000 | 0.975 | 1.027 | 0.973 | 238.337 | 111 | 53.4% | 90.0 | 0.000 |  |
|  |  | Weighted median | 112 | -0.020 | -0.051 | 0.012 | 0.981 | 0.950 | 1.012 | 0.216 |  |  |  |  |  |  |
|  |  | MR-Egger | 112 | -0.028 | -0.079 | 0.024 | 0.973 | 0.924 | 1.024 | 0.293 |  |  |  |  |  | 0.216 |
|  |  | Contamination mixture | 112 | -0.014 | -0.034 | 0.016 | 0.986 | 0.967 | 1.016 | 0.542 |  |  |  |  |  |  |
|  |  | MR-PRESSO** | 107 | 0.002 | -0.021 | 0.024 | 1.002 | 0.979 | 1.024 | 0.895 |  |  |  |  |  |  |
| IS | Asthma | IVW | 23 | -0.058 | -0.169 | 0.053 | 0.944 | 0.844 | 1.055 | 0.308 | 99.239 | 22 | 77.8% | 44.7 | 0.000 |  |
|  |  | Weighted median | 23 | -0.009 | -0.094 | 0.076 | 0.991 | 0.910 | 1.079 | 0.838 |  |  |  |  |  |  |
|  |  | MR-Egger | 23 | -0.043 | -0.595 | 0.510 | 0.958 | 0.552 | 1.665 | 0.880 |  |  |  |  |  | 0.956 |
|  |  | Contamination mixture | 23 | 0.002 | -0.078 | 0.072 | 1.002 | 0.925 | 1.075 | 0.898 |  |  |  |  |  |  |
|  |  | MR-PRESSO** | 19 | -0.053 | -0.115 | 0.010 | 0.949 | 0.891 | 1.010 | 0.115 |  |  |  |  |  |  |

betalow: 95% lower CI for the estimate; betahi: 95% upper CI for the estimate.

ORlow: 95% lower CI for the OR; ORhi: 95% upper CI for the OR.

AF: atrial fibrillation, CAD: coronary artery disease, HF: heart failure, IS: ischemia stroke, MI: myocardial infarction, n: number of SNPs used as instrument variables in each method, SNPs single nucleotide polymorphism, CI confidence intervals, OR odds ratio.

** indicates corrected MR-PRESSO estimates

**Supplement Table 5. Mendelian Randomization for the effect of lifetime smoking on chronic respiratory diseases and cardiovascular diseases**

| **Exposure** | **Outcome** | **Methods** | **n** | **Estimate** | **betalow** | **betahi** | **OR** | **ORlow** | **ORhi** | **p-value** | **Heterogeneity test statistic (Cochran's Q)** | **df** | **I**^2^ | **F-statistic** | **P-het value** | **P**_egger_intercept_ |
| --- | --- | --- | --- | --- | --- | --- | --- | --- | --- | --- | --- | --- | --- | --- | --- | --- |
| Smoking | COPD | IVW | 126 | 0.091 | 0.032 | 0.150 | 1.095 | 1.032 | 1.162 | 0.003 | 575.8285 | 125 | 78.3% | 44.1 | 0.000 |  |
|  |  | Weighted median | 126 | 0.092 | 0.040 | 0.145 | 1.096 | 1.040 | 1.156 | 0.001 |  |  |  |  |  |  |
|  |  | MR-Egger | 126 | 0.426 | 0.199 | 0.653 | 1.531 | 1.220 | 1.922 | 0.000 |  |  |  |  |  | 0.003 |
|  |  | Contamination mixture | 126 | 0.003 | -0.057 | 0.073 | 1.003 | 0.945 | 1.076 | 1.000 |  |  |  |  |  |  |
|  |  | MR-PRESSO | 114 | 0.076 | 0.026 | 0.127 | 1.079 | 1.026 | 1.135 | 0.004 |  |  |  |  |  |  |
| Smoking | Asthma | IVW | 126 | 0.188 | 0.019 | 0.357 | 1.207 | 1.019 | 1.428 | 0.029 | 142.1496 | 125 | 12.1% | 44.1 | 0.140 |  |
|  |  | Weighted median | 126 | 0.250 | 0.121 | 0.378 | 1.284 | 1.129 | 1.460 | 0.000 |  |  |  |  |  |  |
|  |  | MR-Egger | 126 | 0.265 | -0.411 | 0.940 | 1.303 | 0.663 | 2.561 | 0.442 |  |  |  |  |  | 0.817 |
|  |  | Contamination mixture | 126 | 0.398 | 0.238 | 0.508 | 1.489 | 1.269 | 1.662 | 0.000 |  |  |  |  |  |  |
|  |  | MR-PRESSO | 119 | 0.241 | 0.134 | 0.348 | 1.273 | 1.144 | 1.416 | 0.000 |  |  |  |  |  |  |
| Smoking | CAD | IVW | 126 | 0.499 | 0.393 | 0.604 | 1.647 | 1.482 | 1.830 | 0.000 | 324.7559 | 125 | 61.5% | 44.1 | 0.000 |  |
|  |  | Weighted median | 126 | 0.550 | 0.441 | 0.659 | 1.733 | 1.554 | 1.933 | 0.000 |  |  |  |  |  |  |
|  |  | MR-Egger | 126 | 0.064 | -0.357 | 0.484 | 1.066 | 0.700 | 1.623 | 0.767 |  |  |  |  |  | 0.037 |
|  |  | Contamination mixture | 126 | 0.764 | 0.654 | 0.894 | 2.146 | 1.923 | 2.444 | 0.000 |  |  |  |  |  |  |
|  |  | MR-PRESSO | 122 | 0.537 | 0.448 | 0.626 | 1.711 | 1.565 | 1.870 | 0.000 |  |  |  |  |  |  |
| Smoking | MI | IVW | 126 | 0.589 | 0.562 | 0.724 | 1.802 | 1.575 | 2.062 | 0.000 | 230.2148 | 125 | 45.7% | 44.1 | 0.000 |  |
|  |  | Weighted Median | 126 | 0.639 | 0.562 | 0.799 | 1.895 | 1.616 | 2.223 | 0.000 |  |  |  |  |  |  |
|  |  | MR-Egger | 126 | 0.029 | -0.517 | 0.575 | 1.029 | 0.596 | 1.777 | 0.917 |  |  |  |  |  | 0.038 |
|  |  | Contamination mixture | 126 | 0.910 | 0.780 | 1.150 | 2.484 | 2.181 | 3.158 | 0.000 |  |  |  |  |  |  |
|  |  | MR-PRESSO** | 123 | 0.617 | 0.495 | 0.738 | 1.853 | 1.641 | 2.091 | 0.000 |  |  |  |  |  |  |
| Smoking | HF | IVW | 126 | 0.401 | 0.269 | 0.533 | 1.493 | 1.309 | 1.704 | 0.000 | 199.6419 | 125 | 37.4% | 44.1 | 0.000 |  |
|  |  | Weighted Median | 126 | 0.412 | 0.248 | 0.576 | 1.510 | 1.282 | 1.778 | 0.000 |  |  |  |  |  |  |
|  |  | MR-Egger | 126 | 0.268 | -0.260 | 0.797 | 1.308 | 0.771 | 2.219 | 0.320 |  |  |  |  |  | 0.611 |
|  |  | Contamination mixture | 126 | 0.610 | 0.330 | 0.750 | 1.840 | 1.391 | 2.117 | 0.000 |  |  |  |  |  |  |
|  |  | MR-PRESSO | 126 | 0.401 | 0.269 | 0.533 | 1.493 | 1.309 | 1.704 | 0.000 |  |  |  |  |  |  |
| Smoking | AF | IVW | 126 | 0.170 | 0.049 | 0.290 | 1.185 | 1.050 | 1.337 | 0.006 | 232.7816 | 125 | 46.3% | 44.1 | 0.000 |  |
|  |  | Weighted Median | 126 | 0.200 | 0.060 | 0.339 | 1.221 | 1.062 | 1.403 | 0.005 |  |  |  |  |  |  |
|  |  | MR-Egger | 126 | -0.033 | -0.519 | 0.454 | 0.968 | 0.595 | 1.574 | 0.895 |  |  |  |  |  | 0.400 |
|  |  | Contamination mixture | 126 | 0.310 | 0.160 | 0.480 | 1.363 | 1.174 | 1.616 | 0.001 |  |  |  |  |  |  |
|  |  | MR-PRESSO** | 125 | 0.153 | 0.038 | 0.268 | 1.165 | 1.039 | 1.307 | 0.010 |  |  |  |  |  |  |
| Smoking | IS | IVW | 126 | 0.296 | 0.189 | 0.402 | 1.344 | 1.208 | 1.495 | 0.000 | 157.4129 | 125 | 20.6% | 44.1 | 0.026 |  |
|  |  | Weighted Median | 126 | 0.348 | 0.203 | 0.493 | 1.416 | 1.225 | 1.638 | 0.000 |  |  |  |  |  |  |
|  |  | MR-Egger | 126 | 0.171 | -0.264 | 0.605 | 1.186 | 0.768 | 1.832 | 0.441 |  |  |  |  |  | 0.561 |
|  |  | Contamination mixture | 126 | 0.520 | 0.310 | 0.780 | 1.682 | 1.363 | 2.181 | 0.000 |  |  |  |  |  |  |
|  |  | MR-PRESSO | 126 | 0.296 | 0.189 | 0.402 | 1.344 | 1.208 | 1.495 | 0.000 |  |  |  |  |  |  |

betalow: 95% lower CI for the estimate; betahi: 95% upper CI for the estimate.

ORlow: 95% lower CI for the OR; ORhi: 95% upper CI for the OR.

AF: atrial fibrillation, CAD: coronary artery disease, HF: heart failure, IS: ischemia stroke, MI: myocardial infarction, n: number of SNPs used as instrument variables in each method, SNPs single nucleotide polymorphism, CI confidence intervals, OR odds ratio.

** indicates corrected MR-PRESSO estimates

**Supplement Table 6. Multivariable Mendelian Randomization adjusted for lifetime smoking.**

| **Exposure** | **Outcome** | **Methods** | **n** | **Estimate** | **Std Error** | **95%CI** | | **p-value** | **Cochran's Q** | **df** | **p-het** |
| --- | --- | --- | --- | --- | --- | --- | --- | --- | --- | --- | --- |
|  |  |  |  |  |  | **lower** | **upper** |  |  |  |  |
| COPD | MI | MVIVW | 293 | -0.014 | 0.040 | -0.092 | 0.064 | 0.725 | 718.287 | 291 | 0.000 |
| Smoking |  |  | 293 | 0.640 | 0.213 | 0.223 | 1.058 | 0.003 |  |  |  |
| COPD |  | MVEgger | 293 | 0.019 | 0.103 | -0.183 | 0.221 | 0.856 |  |  |  |
| Smoking |  |  | 293 | 0.643 | 0.213 | 0.225 | 1.062 | 0.003 |  |  |  |
| (intercept) |  |  | 293 | -0.001 | 0.002 | -0.005 | 0.003 | 0.730 |  |  |  |
| Asthma | AF | MVIVW | 143 | 0.039 | 0.016 | 0.008 | 0.071 | 0.014 | 298.118 | 141 | 0.000 |
| Smoking |  |  | 143 | 0.491 | 0.267 | -0.032 | 1.014 | 0.066 |  |  |  |
| Asthma |  | MVEgger | 143 | 0.054 | 0.039 | -0.022 | 0.129 | 0.163 |  |  |  |
| Smoking |  |  | 143 | 0.488 | 0.268 | -0.037 | 1.013 | 0.069 |  |  |  |
| (intercept) |  |  | 143 | -0.001 | 0.002 | -0.006 | 0.004 | 0.684 | 297.758 | 140 | 0.000 |
| Asthma | HF | MVIVW | 143 | 0.025 | 0.016 | -0.006 | 0.057 | 0.113 | 211.067 | 141 | 0.000 |
| Smoking |  |  | 143 | 0.279 | 0.263 | -0.236 | 0.794 | 0.288 |  |  |  |
| Asthma |  | MVEgger | 143 | 0.084 | 0.038 | 0.010 | 0.159 | 0.027 |  |  |  |
| Smoking |  |  | 143 | 0.266 | 0.261 | -0.245 | 0.778 | 0.308 |  |  |  |
| (intercept) |  | MVIVW | 143 | -0.004 | 0.002 | -0.009 | 0.001 | 0.088 | 297.758 | 140 | 0.000 |

AF: atrial fibrillation, COPD: chronic obstructive pulmonary diseases, HF: heart failure, IS: ischemia stroke, MI: myocardial infarction, MVIVW: multivariable inverse-variance weighted, MVEgger: multivariable Egger analysis, n: number of SNPs used as instrument variables in each method, SNPs single nucleotide polymorphism, CI confidence intervals, OR odds ratio.

**Supplement Table 7. Cis-Mendelian Randomization for the effect of inflammatory markers on chronic respiratory diseases and cardiovascular diseases based on Wald test/IVW method.**

| **Exposure** | **Outcome** | **n** | **Estimate** | **StdError** | **CI95Lower** | **CI95Upper** | **p-value** | **OR** | **ORlow** | **ORhi** | **q-value** |
| --- | --- | --- | --- | --- | --- | --- | --- | --- | --- | --- | --- |
| ADM | COPD | 3 | -0.019 | 0.015 | -0.049 | 0.011 | 0.213 | 0.981 | 0.952 | 1.011 | 0.623 |
| ADM | Asthma | 3 | 0.000 | 0.044 | -0.086 | 0.086 | 0.997 | 1.000 | 0.917 | 1.090 | 1.000 |
| ADM | CAD | 3 | -0.005 | 0.037 | -0.077 | 0.068 | 0.903 | 0.995 | 0.926 | 1.071 | 0.980 |
| ADM | MI | 3 | 0.004 | 0.058 | -0.109 | 0.117 | 0.949 | 1.004 | 0.896 | 1.124 | 0.997 |
| ADM | HF | 3 | -0.004 | 0.059 | -0.120 | 0.111 | 0.940 | 0.996 | 0.887 | 1.117 | 0.995 |
| ADM | AF | 3 | 0.044 | 0.051 | -0.055 | 0.143 | 0.384 | 1.045 | 0.946 | 1.154 | 0.786 |
| ADM | Stroke | 3 | -0.010 | 0.054 | -0.117 | 0.096 | 0.849 | 0.990 | 0.890 | 1.101 | 0.971 |
| APCS | COPD | 3 | -0.007 | 0.007 | -0.020 | 0.006 | 0.299 | 0.993 | 0.980 | 1.006 | 0.722 |
| APCS | Asthma | 3 | -0.038 | 0.020 | -0.077 | 0.000 | 0.051 | 0.962 | 0.926 | 1.000 | 0.309 |
| APCS | CAD | 3 | -0.009 | 0.016 | -0.040 | 0.022 | 0.561 | 0.991 | 0.961 | 1.022 | 0.896 |
| APCS | MI | 3 | 0.009 | 0.024 | -0.037 | 0.055 | 0.706 | 1.009 | 0.963 | 1.057 | 0.943 |
| APCS | HF | 3 | -0.037 | 0.024 | -0.085 | 0.011 | 0.127 | 0.964 | 0.919 | 1.011 | 0.516 |
| APCS | AF | 3 | -0.029 | 0.021 | -0.071 | 0.013 | 0.177 | 0.972 | 0.932 | 1.013 | 0.594 |
| APCS | Stroke | 3 | -0.006 | 0.022 | -0.050 | 0.038 | 0.794 | 0.994 | 0.952 | 1.039 | 0.971 |
| CCL11 | COPD | 1 | 0.004 | 0.028 | -0.051 | 0.058 | 0.890 | 1.004 | 0.951 | 1.060 | 0.976 |
| CCL11 | Asthma | 1 | 0.073 | 0.080 | -0.083 | 0.229 | 0.360 | 1.076 | 0.920 | 1.257 | 0.767 |
| CCL11 | CAD | 1 | -0.011 | 0.065 | -0.138 | 0.116 | 0.868 | 0.989 | 0.871 | 1.123 | 0.971 |
| CCL11 | MI | 1 | -0.009 | 0.099 | -0.203 | 0.186 | 0.932 | 0.992 | 0.816 | 1.204 | 0.993 |
| CCL11 | HF | 1 | -0.080 | 0.105 | -0.285 | 0.125 | 0.443 | 0.923 | 0.752 | 1.133 | 0.824 |
| CCL11 | AF | 1 | -0.045 | 0.087 | -0.216 | 0.127 | 0.609 | 0.956 | 0.806 | 1.135 | 0.908 |
| CCL11 | Stroke | 1 | -0.013 | 0.092 | -0.194 | 0.168 | 0.886 | 0.987 | 0.823 | 1.183 | 0.975 |
| CCL14 | COPD | 6 | 0.000 | 0.003 | -0.007 | 0.007 | 0.974 | 1.000 | 0.993 | 1.007 | 1.000 |
| CCL14 | Asthma | 6 | 0.024 | 0.011 | 0.001 | 0.046 | 0.038 | 1.024 | 1.001 | 1.047 | 0.264 |
| CCL14 | CAD | 6 | 0.000 | 0.008 | -0.016 | 0.015 | 0.958 | 1.000 | 0.984 | 1.015 | 0.999 |
| CCL14 | MI | 6 | -0.001 | 0.012 | -0.024 | 0.022 | 0.948 | 0.999 | 0.976 | 1.023 | 0.997 |
| CCL14 | HF | 6 | 0.000 | 0.014 | -0.026 | 0.027 | 0.977 | 1.000 | 0.974 | 1.027 | 1.000 |
| CCL14 | AF | 6 | 0.005 | 0.014 | -0.023 | 0.032 | 0.729 | 1.005 | 0.978 | 1.033 | 0.948 |
| CCL14 | Stroke | 6 | 0.008 | 0.014 | -0.021 | 0.036 | 0.595 | 1.008 | 0.980 | 1.036 | 0.900 |
| CCL15 | COPD | 2 | 0.003 | 0.003 | -0.003 | 0.008 | 0.365 | 1.003 | 0.997 | 1.008 | 0.767 |
| CCL15 | Asthma | 2 | -0.020 | 0.008 | -0.036 | -0.004 | 0.014 | 0.980 | 0.965 | 0.996 | 0.144 |
| CCL15 | CAD | 2 | -0.001 | 0.007 | -0.014 | 0.013 | 0.926 | 0.999 | 0.986 | 1.013 | 0.991 |
| CCL15 | MI | 2 | -0.007 | 0.009 | -0.026 | 0.011 | 0.430 | 0.993 | 0.974 | 1.011 | 0.818 |
| CCL15 | HF | 2 | -0.019 | 0.012 | -0.043 | 0.006 | 0.137 | 0.982 | 0.958 | 1.006 | 0.528 |
| CCL15 | AF | 2 | 0.015 | 0.010 | -0.004 | 0.034 | 0.126 | 1.015 | 0.996 | 1.035 | 0.512 |
| CCL15 | Stroke | 2 | 0.016 | 0.010 | -0.004 | 0.036 | 0.118 | 1.016 | 0.996 | 1.037 | 0.490 |
| CCL16 | COPD | 4 | 0.001 | 0.002 | -0.003 | 0.006 | 0.511 | 1.001 | 0.997 | 1.006 | 0.870 |
| CCL16 | Asthma | 4 | 0.005 | 0.014 | -0.022 | 0.033 | 0.703 | 1.005 | 0.978 | 1.033 | 0.942 |
| CCL16 | CAD | 4 | -0.002 | 0.005 | -0.012 | 0.008 | 0.712 | 0.998 | 0.988 | 1.008 | 0.943 |
| CCL16 | MI | 4 | -0.001 | 0.008 | -0.017 | 0.014 | 0.858 | 0.999 | 0.983 | 1.014 | 0.971 |
| CCL16 | HF | 4 | 0.006 | 0.009 | -0.012 | 0.024 | 0.522 | 1.006 | 0.988 | 1.025 | 0.877 |
| CCL16 | AF | 4 | -0.007 | 0.007 | -0.021 | 0.008 | 0.371 | 0.993 | 0.979 | 1.008 | 0.772 |
| CCL16 | Stroke | 4 | -0.002 | 0.008 | -0.017 | 0.013 | 0.770 | 0.998 | 0.983 | 1.013 | 0.965 |
| CCL17 | COPD | 3 | -0.008 | 0.007 | -0.021 | 0.005 | 0.217 | 0.992 | 0.979 | 1.005 | 0.627 |
| CCL17 | Asthma | 3 | -0.054 | 0.019 | -0.091 | -0.017 | 0.004 | 0.947 | 0.913 | 0.983 | 0.062 |
| CCL17 | CAD | 3 | 0.042 | 0.015 | 0.012 | 0.072 | 0.007 | 1.043 | 1.012 | 1.075 | 0.081 |
| CCL17 | MI | 3 | 0.056 | 0.023 | 0.010 | 0.102 | 0.017 | 1.057 | 1.010 | 1.107 | 0.163 |
| CCL17 | HF | 3 | 0.016 | 0.025 | -0.033 | 0.065 | 0.524 | 1.016 | 0.967 | 1.068 | 0.879 |
| CCL17 | AF | 3 | 0.023 | 0.021 | -0.018 | 0.064 | 0.281 | 1.023 | 0.982 | 1.066 | 0.704 |
| CCL17 | Stroke | 3 | 0.030 | 0.023 | -0.015 | 0.076 | 0.191 | 1.031 | 0.985 | 1.079 | 0.605 |
| CCL2 | COPD | 2 | -0.036 | 0.033 | -0.099 | 0.028 | 0.274 | 0.965 | 0.905 | 1.029 | 0.695 |
| CCL2 | Asthma | 2 | -0.022 | 0.093 | -0.204 | 0.160 | 0.813 | 0.978 | 0.815 | 1.174 | 0.971 |
| CCL2 | CAD | 2 | -0.034 | 0.074 | -0.179 | 0.112 | 0.650 | 0.967 | 0.836 | 1.119 | 0.925 |
| CCL2 | MI | 2 | -0.020 | 0.113 | -0.242 | 0.202 | 0.859 | 0.980 | 0.785 | 1.224 | 0.971 |
| CCL2 | HF | 2 | -0.152 | 0.120 | -0.388 | 0.084 | 0.206 | 0.859 | 0.678 | 1.087 | 0.615 |
| CCL2 | AF | 2 | -0.051 | 0.102 | -0.250 | 0.149 | 0.620 | 0.951 | 0.779 | 1.161 | 0.912 |
| CCL2 | Stroke | 2 | 0.219 | 0.109 | 0.006 | 0.433 | 0.044 | 1.245 | 1.006 | 1.543 | 0.287 |
| CCL22 | COPD | 3 | 0.017 | 0.008 | 0.001 | 0.032 | 0.032 | 1.017 | 1.001 | 1.033 | 0.233 |
| CCL22 | Asthma | 3 | 0.052 | 0.022 | 0.009 | 0.095 | 0.018 | 1.053 | 1.009 | 1.100 | 0.167 |
| CCL22 | CAD | 3 | -0.031 | 0.018 | -0.067 | 0.004 | 0.080 | 0.969 | 0.936 | 1.004 | 0.400 |
| CCL22 | MI | 3 | -0.040 | 0.026 | -0.092 | 0.012 | 0.128 | 0.961 | 0.912 | 1.012 | 0.516 |
| CCL22 | HF | 3 | -0.012 | 0.029 | -0.069 | 0.046 | 0.689 | 0.988 | 0.933 | 1.047 | 0.935 |
| CCL22 | AF | 3 | -0.057 | 0.025 | -0.106 | -0.009 | 0.019 | 0.944 | 0.900 | 0.991 | 0.171 |
| CCL22 | Stroke | 3 | -0.005 | 0.027 | -0.059 | 0.048 | 0.851 | 0.995 | 0.943 | 1.050 | 0.971 |
| CCL25 | COPD | 4 | -0.005 | 0.003 | -0.010 | 0.000 | 0.039 | 0.995 | 0.990 | 1.000 | 0.268 |
| CCL25 | Asthma | 4 | -0.004 | 0.007 | -0.018 | 0.010 | 0.576 | 0.996 | 0.982 | 1.010 | 0.896 |
| CCL25 | CAD | 4 | 0.022 | 0.009 | 0.004 | 0.040 | 0.019 | 1.022 | 1.004 | 1.041 | 0.171 |
| CCL25 | MI | 4 | 0.034 | 0.009 | 0.015 | 0.052 | 0.000 | 1.034 | 1.015 | 1.054 | 0.010 |
| CCL25 | HF | 4 | 0.003 | 0.012 | -0.020 | 0.025 | 0.828 | 1.003 | 0.980 | 1.025 | 0.971 |
| CCL25 | AF | 4 | -0.004 | 0.008 | -0.020 | 0.012 | 0.666 | 0.996 | 0.981 | 1.013 | 0.930 |
| CCL25 | Stroke | 4 | 0.007 | 0.009 | -0.010 | 0.024 | 0.430 | 1.007 | 0.990 | 1.024 | 0.818 |
| CCL27 | COPD | 2 | 0.002 | 0.009 | -0.016 | 0.020 | 0.799 | 1.002 | 0.985 | 1.020 | 0.971 |
| CCL27 | Asthma | 2 | 0.003 | 0.026 | -0.048 | 0.054 | 0.908 | 1.003 | 0.953 | 1.055 | 0.982 |
| CCL27 | CAD | 2 | -0.003 | 0.021 | -0.044 | 0.039 | 0.904 | 0.997 | 0.957 | 1.040 | 0.981 |
| CCL27 | MI | 2 | -0.017 | 0.032 | -0.079 | 0.044 | 0.581 | 0.983 | 0.924 | 1.045 | 0.896 |
| CCL27 | HF | 2 | 0.019 | 0.034 | -0.048 | 0.086 | 0.581 | 1.019 | 0.953 | 1.089 | 0.896 |
| CCL27 | AF | 2 | -0.033 | 0.029 | -0.091 | 0.024 | 0.251 | 0.967 | 0.913 | 1.024 | 0.668 |
| CCL27 | Stroke | 2 | -0.065 | 0.031 | -0.126 | -0.005 | 0.035 | 0.937 | 0.882 | 0.995 | 0.245 |
| CCL3 | COPD | 9 | 0.000 | 0.005 | -0.011 | 0.010 | 0.963 | 1.000 | 0.989 | 1.011 | 0.999 |
| CCL3 | Asthma | 10 | 0.019 | 0.026 | -0.032 | 0.070 | 0.461 | 1.019 | 0.969 | 1.073 | 0.838 |
| CCL3 | CAD | 10 | -0.002 | 0.012 | -0.026 | 0.022 | 0.889 | 0.998 | 0.975 | 1.023 | 0.976 |
| CCL3 | MI | 10 | 0.009 | 0.019 | -0.028 | 0.046 | 0.636 | 1.009 | 0.972 | 1.047 | 0.919 |
| CCL3 | HF | 12 | 0.029 | 0.021 | -0.013 | 0.071 | 0.179 | 1.029 | 0.987 | 1.073 | 0.597 |
| CCL3 | AF | 15 | 0.033 | 0.015 | 0.003 | 0.064 | 0.031 | 1.034 | 1.003 | 1.066 | 0.230 |
| CCL3 | Stroke | 10 | 0.023 | 0.021 | -0.018 | 0.064 | 0.264 | 1.024 | 0.983 | 1.066 | 0.684 |
| CCL4 | COPD | 7 | -0.007 | 0.010 | -0.026 | 0.012 | 0.457 | 0.993 | 0.975 | 1.012 | 0.836 |
| CCL4 | Asthma | 9 | -0.013 | 0.020 | -0.053 | 0.027 | 0.516 | 0.987 | 0.948 | 1.027 | 0.872 |
| CCL4 | CAD | 9 | 0.010 | 0.019 | -0.027 | 0.047 | 0.594 | 1.010 | 0.973 | 1.049 | 0.900 |
| CCL4 | MI | 7 | 0.038 | 0.042 | -0.045 | 0.122 | 0.366 | 1.039 | 0.956 | 1.129 | 0.769 |
| CCL4 | HF | 8 | -0.018 | 0.061 | -0.138 | 0.101 | 0.766 | 0.982 | 0.871 | 1.107 | 0.965 |
| CCL4 | AF | 12 | 0.034 | 0.029 | -0.023 | 0.092 | 0.245 | 1.035 | 0.977 | 1.096 | 0.658 |
| CCL4 | Stroke | 9 | -0.008 | 0.025 | -0.058 | 0.042 | 0.763 | 0.992 | 0.944 | 1.043 | 0.965 |
| CCL7 | COPD | 3 | -0.004 | 0.004 | -0.013 | 0.004 | 0.320 | 0.996 | 0.987 | 1.004 | 0.731 |
| CCL7 | Asthma | 3 | 0.011 | 0.013 | -0.014 | 0.036 | 0.395 | 1.011 | 0.986 | 1.037 | 0.796 |
| CCL7 | CAD | 3 | -0.002 | 0.011 | -0.024 | 0.020 | 0.848 | 0.998 | 0.976 | 1.020 | 0.971 |
| CCL7 | MI | 1 | -0.007 | 0.019 | -0.044 | 0.029 | 0.700 | 0.993 | 0.957 | 1.030 | 0.942 |
| CCL7 | HF | 1 | 0.010 | 0.020 | -0.030 | 0.050 | 0.631 | 1.010 | 0.970 | 1.051 | 0.916 |
| CCL7 | AF | 4 | 0.031 | 0.016 | -0.001 | 0.062 | 0.057 | 1.031 | 0.999 | 1.064 | 0.326 |
| CCL7 | Stroke | 2 | 0.002 | 0.022 | -0.041 | 0.046 | 0.927 | 1.002 | 0.959 | 1.047 | 0.992 |
| CCL8 | COPD | 5 | -0.005 | 0.002 | -0.009 | -0.001 | 0.016 | 0.995 | 0.991 | 0.999 | 0.157 |
| CCL8 | Asthma | 5 | 0.001 | 0.005 | -0.009 | 0.012 | 0.826 | 1.001 | 0.991 | 1.012 | 0.971 |
| CCL8 | CAD | 5 | 0.002 | 0.008 | -0.014 | 0.019 | 0.782 | 1.002 | 0.986 | 1.019 | 0.971 |
| CCL8 | MI | 5 | -0.002 | 0.006 | -0.014 | 0.011 | 0.802 | 0.998 | 0.986 | 1.011 | 0.971 |
| CCL8 | HF | 5 | 0.011 | 0.007 | -0.003 | 0.025 | 0.122 | 1.011 | 0.997 | 1.025 | 0.504 |
| CCL8 | AF | 5 | 0.009 | 0.006 | -0.003 | 0.021 | 0.139 | 1.009 | 0.997 | 1.021 | 0.529 |
| CCL8 | Stroke | 5 | 0.009 | 0.007 | -0.004 | 0.022 | 0.190 | 1.009 | 0.996 | 1.022 | 0.605 |
| CRP | COPD | 5 | 0.024 | 0.008 | 0.008 | 0.041 | 0.004 | 1.025 | 1.008 | 1.042 | 0.055 |
| CRP | Asthma | 5 | 0.001 | 0.024 | -0.046 | 0.048 | 0.974 | 1.001 | 0.955 | 1.049 | 1.000 |
| CRP | CAD | 5 | -0.012 | 0.023 | -0.058 | 0.033 | 0.597 | 0.988 | 0.944 | 1.034 | 0.901 |
| CRP | MI | 5 | 0.009 | 0.031 | -0.051 | 0.069 | 0.769 | 1.009 | 0.950 | 1.071 | 0.965 |
| CRP | HF | 5 | 0.005 | 0.032 | -0.057 | 0.068 | 0.868 | 1.005 | 0.944 | 1.070 | 0.971 |
| CRP | AF | 5 | 0.027 | 0.035 | -0.042 | 0.095 | 0.445 | 1.027 | 0.959 | 1.100 | 0.824 |
| CRP | Stroke | 4 | -0.013 | 0.037 | -0.086 | 0.060 | 0.727 | 0.987 | 0.918 | 1.062 | 0.948 |
| CSF1 | COPD | 3 | 0.009 | 0.013 | -0.016 | 0.034 | 0.480 | 1.009 | 0.984 | 1.034 | 0.848 |
| CSF1 | Asthma | 3 | -0.016 | 0.036 | -0.087 | 0.055 | 0.664 | 0.984 | 0.917 | 1.057 | 0.929 |
| CSF1 | CAD | 3 | 0.176 | 0.030 | 0.117 | 0.234 | 0.000 | 1.192 | 1.125 | 1.263 | 0.000 |
| CSF1 | MI | 3 | 0.207 | 0.045 | 0.119 | 0.295 | 0.000 | 1.230 | 1.126 | 1.343 | 0.000 |
| CSF1 | HF | 3 | 0.129 | 0.047 | 0.036 | 0.222 | 0.007 | 1.137 | 1.036 | 1.248 | 0.083 |
| CSF1 | AF | 3 | 0.042 | 0.040 | -0.037 | 0.120 | 0.297 | 1.043 | 0.964 | 1.128 | 0.719 |
| CSF1 | Stroke | 3 | 0.068 | 0.043 | -0.017 | 0.152 | 0.115 | 1.070 | 0.984 | 1.165 | 0.487 |
| CX3CL1 | COPD | 8 | -0.012 | 0.006 | -0.025 | 0.000 | 0.058 | 0.988 | 0.976 | 1.000 | 0.333 |
| CX3CL1 | Asthma | 8 | -0.059 | 0.023 | -0.104 | -0.014 | 0.011 | 0.943 | 0.901 | 0.987 | 0.118 |
| CX3CL1 | CAD | 8 | 0.078 | 0.015 | 0.048 | 0.107 | 0.000 | 1.081 | 1.049 | 1.113 | 0.000 |
| CX3CL1 | MI | 8 | 0.113 | 0.023 | 0.068 | 0.158 | 0.000 | 1.120 | 1.071 | 1.172 | 0.000 |
| CX3CL1 | HF | 8 | 0.032 | 0.027 | -0.021 | 0.085 | 0.242 | 1.032 | 0.979 | 1.088 | 0.657 |
| CX3CL1 | AF | 8 | 0.020 | 0.023 | -0.025 | 0.065 | 0.386 | 1.020 | 0.975 | 1.067 | 0.786 |
| CX3CL1 | Stroke | 8 | 0.054 | 0.022 | 0.010 | 0.097 | 0.015 | 1.055 | 1.010 | 1.102 | 0.152 |
| CXCL1 | COPD | 6 | -0.017 | 0.009 | -0.034 | 0.000 | 0.052 | 0.983 | 0.967 | 1.000 | 0.314 |
| CXCL1 | Asthma | 6 | -0.008 | 0.020 | -0.048 | 0.031 | 0.677 | 0.992 | 0.954 | 1.031 | 0.932 |
| CXCL1 | CAD | 6 | -0.025 | 0.014 | -0.053 | 0.003 | 0.080 | 0.975 | 0.948 | 1.003 | 0.399 |
| CXCL1 | MI | 6 | -0.002 | 0.022 | -0.046 | 0.041 | 0.911 | 0.998 | 0.955 | 1.042 | 0.984 |
| CXCL1 | HF | 6 | 0.032 | 0.024 | -0.014 | 0.079 | 0.172 | 1.033 | 0.986 | 1.082 | 0.587 |
| CXCL1 | AF | 6 | -0.026 | 0.032 | -0.089 | 0.038 | 0.429 | 0.975 | 0.915 | 1.039 | 0.818 |
| CXCL1 | Stroke | 6 | -0.012 | 0.021 | -0.053 | 0.030 | 0.591 | 0.989 | 0.948 | 1.031 | 0.899 |
| CXCL10 | COPD | 1 | -0.009 | 0.028 | -0.063 | 0.045 | 0.744 | 0.991 | 0.939 | 1.046 | 0.955 |
| CXCL10 | Asthma | 1 | 0.041 | 0.080 | -0.115 | 0.198 | 0.604 | 1.042 | 0.891 | 1.219 | 0.906 |
| CXCL10 | CAD | 1 | 0.020 | 0.069 | -0.115 | 0.154 | 0.775 | 1.020 | 0.891 | 1.167 | 0.965 |
| CXCL10 | MI | 1 | 0.125 | 0.102 | -0.074 | 0.325 | 0.218 | 1.134 | 0.929 | 1.384 | 0.627 |
| CXCL10 | HF | 1 | 0.105 | 0.106 | -0.103 | 0.313 | 0.323 | 1.110 | 0.902 | 1.367 | 0.733 |
| CXCL10 | AF | 1 | -0.200 | 0.089 | -0.374 | -0.026 | 0.024 | 0.819 | 0.688 | 0.974 | 0.198 |
| CXCL10 | Stroke | 1 | -0.099 | 0.093 | -0.281 | 0.082 | 0.284 | 0.905 | 0.755 | 1.086 | 0.708 |
| CXCL11 | COPD | 1 | 0.003 | 0.009 | -0.015 | 0.021 | 0.739 | 1.003 | 0.985 | 1.022 | 0.953 |
| CXCL11 | Asthma | 1 | -0.019 | 0.027 | -0.072 | 0.033 | 0.477 | 0.981 | 0.931 | 1.034 | 0.847 |
| CXCL11 | CAD | 1 | 0.001 | 0.029 | -0.055 | 0.057 | 0.967 | 1.001 | 0.946 | 1.059 | 1.000 |
| CXCL11 | AF | 1 | 0.021 | 0.058 | -0.093 | 0.136 | 0.714 | 1.022 | 0.911 | 1.145 | 0.944 |
| CXCL16 | COPD | 9 | 0.006 | 0.008 | -0.008 | 0.021 | 0.397 | 1.006 | 0.992 | 1.021 | 0.798 |
| CXCL16 | Asthma | 9 | -0.042 | 0.022 | -0.084 | 0.001 | 0.055 | 0.959 | 0.919 | 1.001 | 0.321 |
| CXCL16 | CAD | 9 | 0.108 | 0.021 | 0.067 | 0.150 | 0.000 | 1.114 | 1.069 | 1.161 | 0.000 |
| CXCL16 | MI | 9 | 0.102 | 0.029 | 0.046 | 0.158 | 0.000 | 1.107 | 1.047 | 1.171 | 0.010 |
| CXCL16 | HF | 9 | 0.054 | 0.042 | -0.027 | 0.136 | 0.192 | 1.056 | 0.973 | 1.145 | 0.605 |
| CXCL16 | AF | 9 | 0.096 | 0.035 | 0.027 | 0.166 | 0.007 | 1.101 | 1.027 | 1.180 | 0.081 |
| CXCL16 | Stroke | 9 | 0.029 | 0.035 | -0.041 | 0.098 | 0.419 | 1.029 | 0.960 | 1.103 | 0.812 |
| CXCL6 | COPD | 12 | -0.014 | 0.004 | -0.021 | -0.007 | 0.000 | 0.986 | 0.979 | 0.994 | 0.006 |
| CXCL6 | Asthma | 12 | -0.003 | 0.014 | -0.030 | 0.025 | 0.845 | 0.997 | 0.970 | 1.025 | 0.971 |
| CXCL6 | CAD | 12 | 0.012 | 0.010 | -0.008 | 0.032 | 0.238 | 1.012 | 0.992 | 1.032 | 0.653 |
| CXCL6 | MI | 12 | 0.031 | 0.013 | 0.006 | 0.055 | 0.014 | 1.031 | 1.006 | 1.057 | 0.140 |
| CXCL6 | HF | 12 | 0.034 | 0.013 | 0.008 | 0.059 | 0.010 | 1.034 | 1.008 | 1.061 | 0.110 |
| CXCL6 | AF | 12 | 0.041 | 0.013 | 0.016 | 0.065 | 0.001 | 1.042 | 1.016 | 1.068 | 0.022 |
| CXCL6 | Stroke | 12 | 0.005 | 0.012 | -0.018 | 0.028 | 0.681 | 1.005 | 0.982 | 1.029 | 0.932 |
| CXCL8 | COPD | 1 | 0.018 | 0.028 | -0.036 | 0.072 | 0.508 | 1.018 | 0.965 | 1.075 | 0.870 |
| CXCL8 | Asthma | 1 | 0.141 | 0.077 | -0.010 | 0.291 | 0.067 | 1.151 | 0.990 | 1.337 | 0.359 |
| CXCL8 | CAD | 1 | 0.045 | 0.061 | -0.074 | 0.164 | 0.456 | 1.046 | 0.929 | 1.178 | 0.835 |
| CXCL8 | MI | 1 | 0.180 | 0.087 | 0.009 | 0.350 | 0.039 | 1.197 | 1.009 | 1.419 | 0.265 |
| CXCL8 | HF | 1 | -0.213 | 0.112 | -0.431 | 0.006 | 0.056 | 0.808 | 0.650 | 1.006 | 0.324 |
| CXCL8 | AF | 1 | -0.168 | 0.092 | -0.348 | 0.012 | 0.067 | 0.845 | 0.706 | 1.012 | 0.359 |
| CXCL8 | Stroke | 1 | 0.013 | 0.100 | -0.182 | 0.208 | 0.899 | 1.013 | 0.833 | 1.231 | 0.980 |
| CXCL9 | COPD | 1 | 0.005 | 0.016 | -0.026 | 0.035 | 0.759 | 1.005 | 0.975 | 1.036 | 0.964 |
| CXCL9 | Asthma | 1 | 0.006 | 0.045 | -0.082 | 0.094 | 0.898 | 1.006 | 0.921 | 1.099 | 0.980 |
| CXCL9 | CAD | 1 | 0.071 | 0.038 | -0.003 | 0.145 | 0.060 | 1.074 | 0.997 | 1.156 | 0.337 |
| CXCL9 | MI | 1 | 0.007 | 0.055 | -0.102 | 0.115 | 0.900 | 1.007 | 0.903 | 1.122 | 0.980 |
| CXCL9 | HF | 1 | -0.049 | 0.058 | -0.164 | 0.065 | 0.397 | 0.952 | 0.849 | 1.067 | 0.798 |
| CXCL9 | AF | 1 | 0.059 | 0.049 | -0.038 | 0.156 | 0.232 | 1.061 | 0.963 | 1.169 | 0.645 |
| CXCL9 | Stroke | 1 | 0.080 | 0.051 | -0.020 | 0.181 | 0.118 | 1.084 | 0.980 | 1.198 | 0.491 |
| F2 | COPD | 1 | 0.025 | 0.021 | -0.016 | 0.066 | 0.236 | 1.025 | 0.984 | 1.069 | 0.650 |
| F2 | Asthma | 1 | 0.071 | 0.061 | -0.048 | 0.191 | 0.240 | 1.074 | 0.953 | 1.210 | 0.655 |
| F2 | CAD | 1 | 0.152 | 0.050 | 0.054 | 0.251 | 0.002 | 1.165 | 1.056 | 1.285 | 0.040 |
| F2 | MI | 1 | 0.293 | 0.077 | 0.142 | 0.443 | 0.000 | 1.340 | 1.153 | 1.558 | 0.005 |
| F2 | HF | 1 | 0.069 | 0.079 | -0.086 | 0.225 | 0.382 | 1.072 | 0.917 | 1.252 | 0.783 |
| F2 | AF | 1 | -0.002 | 0.067 | -0.134 | 0.130 | 0.976 | 0.998 | 0.875 | 1.139 | 1.000 |
| F2 | Stroke | 1 | 0.297 | 0.073 | 0.154 | 0.441 | 0.000 | 1.346 | 1.166 | 1.555 | 0.002 |
| FGF23 | COPD | 2 | 0.033 | 0.022 | -0.010 | 0.077 | 0.134 | 1.034 | 0.990 | 1.080 | 0.521 |
| FGF23 | Asthma | 2 | -0.015 | 0.064 | -0.141 | 0.111 | 0.820 | 0.985 | 0.869 | 1.118 | 0.971 |
| FGF23 | CAD | 2 | 0.071 | 0.055 | -0.036 | 0.179 | 0.191 | 1.074 | 0.965 | 1.196 | 0.605 |
| FGF23 | MI | 2 | 0.082 | 0.080 | -0.076 | 0.240 | 0.309 | 1.085 | 0.927 | 1.271 | 0.727 |
| FGF23 | HF | 2 | -0.146 | 0.085 | -0.312 | 0.020 | 0.084 | 0.864 | 0.732 | 1.020 | 0.409 |
| FGF23 | AF | 2 | -0.121 | 0.072 | -0.263 | 0.020 | 0.093 | 0.886 | 0.769 | 1.020 | 0.439 |
| FGF23 | Stroke | 2 | -0.075 | 0.086 | -0.243 | 0.093 | 0.381 | 0.928 | 0.785 | 1.097 | 0.782 |
| FGF7 | Asthma | 1 | -0.049 | 0.034 | -0.117 | 0.018 | 0.151 | 0.952 | 0.890 | 1.018 | 0.550 |
| FGF7 | CAD | 1 | 0.032 | 0.028 | -0.023 | 0.087 | 0.252 | 1.033 | 0.977 | 1.091 | 0.669 |
| FGF7 | MI | 1 | 0.064 | 0.043 | -0.019 | 0.148 | 0.132 | 1.066 | 0.981 | 1.159 | 0.521 |
| FGF7 | HF | 1 | 0.079 | 0.045 | -0.008 | 0.167 | 0.075 | 1.083 | 0.992 | 1.181 | 0.382 |
| FGF7 | AF | 1 | 0.126 | 0.038 | 0.051 | 0.201 | 0.001 | 1.135 | 1.053 | 1.223 | 0.020 |
| FGF7 | Stroke | 1 | 0.036 | 0.040 | -0.042 | 0.114 | 0.368 | 1.037 | 0.959 | 1.121 | 0.769 |
| FGG | COPD | 2 | -0.013 | 0.013 | -0.038 | 0.013 | 0.344 | 0.988 | 0.962 | 1.014 | 0.752 |
| FGG | Asthma | 2 | 0.020 | 0.038 | -0.055 | 0.095 | 0.603 | 1.020 | 0.947 | 1.099 | 0.906 |
| FGG | CAD | 2 | -0.061 | 0.031 | -0.121 | 0.000 | 0.049 | 0.941 | 0.886 | 1.000 | 0.302 |
| FGG | MI | 2 | -0.020 | 0.046 | -0.111 | 0.071 | 0.665 | 0.980 | 0.895 | 1.073 | 0.930 |
| FGG | HF | 2 | 0.009 | 0.049 | -0.086 | 0.105 | 0.851 | 1.009 | 0.917 | 1.110 | 0.971 |
| FGG | AF | 2 | -0.023 | 0.042 | -0.106 | 0.059 | 0.576 | 0.977 | 0.900 | 1.060 | 0.896 |
| FGG | Stroke | 2 | 0.014 | 0.043 | -0.071 | 0.099 | 0.740 | 1.014 | 0.932 | 1.104 | 0.953 |
| HGF | COPD | 3 | 0.004 | 0.016 | -0.027 | 0.034 | 0.812 | 1.004 | 0.974 | 1.035 | 0.971 |
| HGF | Asthma | 3 | -0.050 | 0.045 | -0.139 | 0.038 | 0.264 | 0.951 | 0.870 | 1.039 | 0.684 |
| HGF | CAD | 3 | -0.011 | 0.038 | -0.087 | 0.064 | 0.771 | 0.989 | 0.917 | 1.066 | 0.965 |
| HGF | MI | 3 | 0.012 | 0.058 | -0.102 | 0.127 | 0.830 | 1.013 | 0.903 | 1.135 | 0.971 |
| HGF | HF | 3 | 0.089 | 0.059 | -0.026 | 0.205 | 0.130 | 1.093 | 0.974 | 1.227 | 0.518 |
| HGF | AF | 3 | -0.061 | 0.050 | -0.160 | 0.038 | 0.225 | 0.941 | 0.852 | 1.038 | 0.637 |
| HGF | Stroke | 3 | 0.057 | 0.054 | -0.049 | 0.163 | 0.291 | 1.059 | 0.952 | 1.177 | 0.718 |
| HP | COPD | 13 | -0.006 | 0.002 | -0.010 | -0.002 | 0.003 | 0.994 | 0.990 | 0.998 | 0.048 |
| HP | Asthma | 13 | -0.017 | 0.006 | -0.028 | -0.005 | 0.005 | 0.984 | 0.972 | 0.995 | 0.068 |
| HP | CAD | 12 | -0.020 | 0.008 | -0.035 | -0.004 | 0.016 | 0.981 | 0.965 | 0.996 | 0.159 |
| HP | MI | 12 | -0.019 | 0.013 | -0.043 | 0.006 | 0.134 | 0.981 | 0.957 | 1.006 | 0.521 |
| HP | HF | 12 | 0.004 | 0.007 | -0.009 | 0.017 | 0.517 | 1.004 | 0.991 | 1.017 | 0.872 |
| HP | AF | 12 | 0.002 | 0.006 | -0.009 | 0.013 | 0.713 | 1.002 | 0.991 | 1.013 | 0.943 |
| HP | Stroke | 12 | -0.013 | 0.006 | -0.025 | -0.001 | 0.031 | 0.987 | 0.975 | 0.999 | 0.231 |
| ICAM1 | COPD | 6 | -0.014 | 0.010 | -0.033 | 0.006 | 0.165 | 0.987 | 0.968 | 1.006 | 0.576 |
| ICAM1 | Asthma | 6 | 0.020 | 0.029 | -0.036 | 0.076 | 0.480 | 1.020 | 0.965 | 1.079 | 0.848 |
| ICAM1 | CAD | 6 | 0.009 | 0.015 | -0.021 | 0.039 | 0.546 | 1.009 | 0.979 | 1.040 | 0.891 |
| ICAM1 | MI | 6 | -0.018 | 0.024 | -0.065 | 0.028 | 0.433 | 0.982 | 0.937 | 1.028 | 0.819 |
| ICAM1 | HF | 7 | -0.030 | 0.023 | -0.076 | 0.015 | 0.188 | 0.970 | 0.927 | 1.015 | 0.605 |
| ICAM1 | AF | 7 | -0.030 | 0.020 | -0.069 | 0.009 | 0.136 | 0.971 | 0.934 | 1.009 | 0.527 |
| ICAM1 | Stroke | 6 | 0.010 | 0.023 | -0.034 | 0.055 | 0.647 | 1.010 | 0.966 | 1.057 | 0.924 |
| IL12RB2 | COPD | 1 | 0.017 | 0.009 | 0.000 | 0.035 | 0.047 | 1.018 | 1.000 | 1.035 | 0.298 |
| IL12RB2 | Asthma | 1 | -0.031 | 0.025 | -0.080 | 0.018 | 0.213 | 0.969 | 0.923 | 1.018 | 0.623 |
| IL12RB2 | CAD | 1 | -0.012 | 0.021 | -0.052 | 0.029 | 0.571 | 0.988 | 0.949 | 1.029 | 0.896 |
| IL12RB2 | MI | 1 | -0.021 | 0.031 | -0.081 | 0.040 | 0.507 | 0.980 | 0.922 | 1.041 | 0.870 |
| IL12RB2 | HF | 1 | 0.007 | 0.032 | -0.056 | 0.071 | 0.819 | 1.007 | 0.946 | 1.073 | 0.971 |
| IL12RB2 | AF | 1 | 0.027 | 0.028 | -0.027 | 0.081 | 0.324 | 1.028 | 0.973 | 1.085 | 0.733 |
| IL12RB2 | Stroke | 1 | -0.058 | 0.030 | -0.116 | 0.001 | 0.055 | 0.944 | 0.890 | 1.001 | 0.321 |
| IL16 | COPD | 9 | -0.006 | 0.005 | -0.015 | 0.003 | 0.168 | 0.994 | 0.985 | 1.003 | 0.576 |
| IL16 | Asthma | 9 | -0.004 | 0.015 | -0.033 | 0.026 | 0.811 | 0.996 | 0.967 | 1.026 | 0.971 |
| IL16 | CAD | 9 | -0.008 | 0.013 | -0.033 | 0.018 | 0.546 | 0.992 | 0.967 | 1.018 | 0.891 |
| IL16 | MI | 9 | 0.003 | 0.022 | -0.040 | 0.047 | 0.878 | 1.003 | 0.961 | 1.048 | 0.971 |
| IL16 | HF | 9 | -0.001 | 0.015 | -0.030 | 0.028 | 0.960 | 0.999 | 0.971 | 1.028 | 0.999 |
| IL16 | AF | 9 | 0.005 | 0.014 | -0.022 | 0.032 | 0.693 | 1.005 | 0.979 | 1.033 | 0.937 |
| IL16 | Stroke | 9 | -0.010 | 0.014 | -0.038 | 0.017 | 0.468 | 0.990 | 0.963 | 1.017 | 0.838 |
| IL17D | COPD | 1 | -0.002 | 0.009 | -0.020 | 0.016 | 0.795 | 0.998 | 0.980 | 1.016 | 0.971 |
| IL17D | Asthma | 1 | -0.011 | 0.027 | -0.063 | 0.041 | 0.674 | 0.989 | 0.939 | 1.042 | 0.932 |
| IL17D | CAD | 1 | -0.006 | 0.022 | -0.049 | 0.036 | 0.766 | 0.994 | 0.952 | 1.037 | 0.965 |
| IL17D | MI | 1 | -0.030 | 0.033 | -0.095 | 0.035 | 0.369 | 0.971 | 0.910 | 1.036 | 0.770 |
| IL17D | HF | 1 | -0.023 | 0.041 | -0.103 | 0.057 | 0.569 | 0.977 | 0.902 | 1.058 | 0.896 |
| IL17D | AF | 1 | 0.007 | 0.032 | -0.055 | 0.069 | 0.835 | 1.007 | 0.946 | 1.071 | 0.971 |
| IL17D | Stroke | 1 | -0.051 | 0.032 | -0.115 | 0.013 | 0.117 | 0.950 | 0.892 | 1.013 | 0.490 |
| IL17RD | COPD | 5 | -0.034 | 0.010 | -0.053 | -0.014 | 0.001 | 0.967 | 0.948 | 0.986 | 0.020 |
| IL17RD | Asthma | 5 | -0.010 | 0.017 | -0.044 | 0.023 | 0.545 | 0.990 | 0.957 | 1.023 | 0.891 |
| IL17RD | CAD | 5 | -0.006 | 0.014 | -0.033 | 0.021 | 0.668 | 0.994 | 0.968 | 1.021 | 0.932 |
| IL17RD | MI | 5 | 0.002 | 0.013 | -0.024 | 0.028 | 0.876 | 1.002 | 0.976 | 1.028 | 0.971 |
| IL17RD | HF | 5 | 0.007 | 0.014 | -0.021 | 0.035 | 0.626 | 1.007 | 0.979 | 1.035 | 0.912 |
| IL17RD | AF | 5 | -0.035 | 0.019 | -0.072 | 0.002 | 0.063 | 0.966 | 0.931 | 1.002 | 0.349 |
| IL17RD | Stroke | 5 | 0.020 | 0.015 | -0.010 | 0.050 | 0.184 | 1.021 | 0.990 | 1.052 | 0.604 |
| IL18 | COPD | 6 | 0.034 | 0.008 | 0.019 | 0.049 | 0.000 | 1.035 | 1.019 | 1.051 | 0.001 |
| IL18 | Asthma | 6 | 0.010 | 0.024 | -0.036 | 0.057 | 0.660 | 1.010 | 0.965 | 1.058 | 0.928 |
| IL18 | CAD | 6 | -0.004 | 0.019 | -0.041 | 0.033 | 0.833 | 0.996 | 0.960 | 1.033 | 0.971 |
| IL18 | MI | 6 | 0.011 | 0.029 | -0.046 | 0.067 | 0.711 | 1.011 | 0.955 | 1.069 | 0.943 |
| IL18 | HF | 6 | 0.039 | 0.029 | -0.019 | 0.097 | 0.183 | 1.040 | 0.982 | 1.102 | 0.603 |
| IL18 | AF | 6 | 0.075 | 0.028 | 0.021 | 0.129 | 0.007 | 1.078 | 1.021 | 1.137 | 0.083 |
| IL18 | Stroke | 5 | -0.017 | 0.029 | -0.074 | 0.039 | 0.552 | 0.983 | 0.929 | 1.040 | 0.893 |
| IL1R1 | COPD | 5 | 0.006 | 0.024 | -0.040 | 0.053 | 0.786 | 1.006 | 0.961 | 1.054 | 0.971 |
| IL1R1 | Asthma | 5 | 0.186 | 0.158 | -0.123 | 0.496 | 0.238 | 1.205 | 0.884 | 1.642 | 0.653 |
| IL1R1 | CAD | 5 | 0.084 | 0.033 | 0.020 | 0.148 | 0.010 | 1.088 | 1.020 | 1.160 | 0.110 |
| IL1R1 | MI | 5 | 0.123 | 0.057 | 0.012 | 0.234 | 0.029 | 1.131 | 1.012 | 1.264 | 0.224 |
| IL1R1 | HF | 5 | -0.058 | 0.041 | -0.137 | 0.022 | 0.155 | 0.944 | 0.872 | 1.022 | 0.557 |
| IL1R1 | AF | 5 | -0.073 | 0.041 | -0.153 | 0.007 | 0.074 | 0.930 | 0.858 | 1.007 | 0.382 |
| IL1R1 | Stroke | 5 | 0.072 | 0.036 | 0.001 | 0.142 | 0.048 | 1.074 | 1.001 | 1.153 | 0.300 |
| IL1R2 | COPD | 14 | -0.018 | 0.009 | -0.035 | -0.002 | 0.032 | 0.982 | 0.965 | 0.998 | 0.233 |
| IL1R2 | Asthma | 14 | -0.114 | 0.044 | -0.201 | -0.027 | 0.010 | 0.892 | 0.818 | 0.973 | 0.110 |
| IL1R2 | CAD | 12 | -0.020 | 0.010 | -0.039 | -0.001 | 0.043 | 0.980 | 0.962 | 0.999 | 0.284 |
| IL1R2 | MI | 11 | -0.023 | 0.018 | -0.058 | 0.011 | 0.184 | 0.977 | 0.944 | 1.011 | 0.603 |
| IL1R2 | HF | 11 | 0.044 | 0.014 | 0.017 | 0.071 | 0.002 | 1.045 | 1.017 | 1.074 | 0.029 |
| IL1R2 | AF | 12 | -0.010 | 0.012 | -0.033 | 0.013 | 0.401 | 0.990 | 0.967 | 1.013 | 0.800 |
| IL1R2 | Stroke | 11 | 0.016 | 0.014 | -0.011 | 0.043 | 0.246 | 1.016 | 0.989 | 1.044 | 0.659 |
| IL1RAP | COPD | 32 | -0.002 | 0.002 | -0.005 | 0.001 | 0.134 | 0.998 | 0.995 | 1.001 | 0.521 |
| IL1RAP | Asthma | 31 | -0.010 | 0.005 | -0.021 | 0.000 | 0.050 | 0.990 | 0.980 | 1.000 | 0.307 |
| IL1RAP | CAD | 32 | 0.006 | 0.004 | -0.001 | 0.014 | 0.108 | 1.006 | 0.999 | 1.014 | 0.475 |
| IL1RAP | MI | 30 | 0.005 | 0.006 | -0.007 | 0.016 | 0.417 | 1.005 | 0.993 | 1.017 | 0.810 |
| IL1RAP | HF | 30 | -0.013 | 0.006 | -0.025 | 0.000 | 0.043 | 0.987 | 0.975 | 1.000 | 0.285 |
| IL1RAP | AF | 32 | 0.003 | 0.006 | -0.009 | 0.014 | 0.641 | 1.003 | 0.991 | 1.014 | 0.921 |
| IL1RAP | Stroke | 29 | -0.001 | 0.007 | -0.015 | 0.013 | 0.938 | 0.999 | 0.986 | 1.014 | 0.995 |
| IL1RL1 | COPD | 21 | -0.023 | 0.005 | -0.032 | -0.013 | 0.000 | 0.978 | 0.968 | 0.987 | 0.000 |
| IL1RL1 | Asthma | 21 | -0.123 | 0.036 | -0.194 | -0.052 | 0.001 | 0.884 | 0.824 | 0.949 | 0.016 |
| IL1RL1 | CAD | 21 | 0.013 | 0.007 | -0.001 | 0.028 | 0.079 | 1.013 | 0.999 | 1.028 | 0.395 |
| IL1RL1 | MI | 21 | 0.014 | 0.009 | -0.003 | 0.031 | 0.110 | 1.014 | 0.997 | 1.032 | 0.475 |
| IL1RL1 | HF | 21 | 0.001 | 0.009 | -0.016 | 0.018 | 0.910 | 1.001 | 0.984 | 1.018 | 0.984 |
| IL1RL1 | AF | 21 | -0.008 | 0.009 | -0.024 | 0.009 | 0.367 | 0.992 | 0.976 | 1.009 | 0.769 |
| IL1RL1 | Stroke | 20 | 0.005 | 0.008 | -0.010 | 0.021 | 0.507 | 1.005 | 0.990 | 1.021 | 0.870 |
| IL1RL2 | COPD | 15 | -0.026 | 0.009 | -0.043 | -0.008 | 0.004 | 0.975 | 0.957 | 0.992 | 0.062 |
| IL1RL2 | Asthma | 15 | -0.070 | 0.043 | -0.154 | 0.014 | 0.104 | 0.932 | 0.857 | 1.014 | 0.462 |
| IL1RL2 | CAD | 15 | -0.048 | 0.014 | -0.076 | -0.020 | 0.001 | 0.953 | 0.927 | 0.980 | 0.017 |
| IL1RL2 | MI | 15 | -0.040 | 0.020 | -0.079 | -0.002 | 0.041 | 0.961 | 0.924 | 0.998 | 0.276 |
| IL1RL2 | HF | 15 | 0.005 | 0.017 | -0.030 | 0.039 | 0.794 | 1.005 | 0.971 | 1.039 | 0.971 |
| IL1RL2 | AF | 16 | -0.004 | 0.015 | -0.033 | 0.026 | 0.800 | 0.996 | 0.967 | 1.026 | 0.971 |
| IL1RL2 | Stroke | 15 | -0.001 | 0.015 | -0.032 | 0.029 | 0.933 | 0.999 | 0.969 | 1.029 | 0.993 |
| IL1RN | COPD | 13 | -0.028 | 0.009 | -0.046 | -0.011 | 0.001 | 0.972 | 0.955 | 0.989 | 0.026 |
| IL1RN | Asthma | 14 | -0.055 | 0.023 | -0.100 | -0.009 | 0.019 | 0.947 | 0.905 | 0.991 | 0.171 |
| IL1RN | CAD | 14 | 0.083 | 0.019 | 0.045 | 0.121 | 0.000 | 1.087 | 1.046 | 1.129 | 0.001 |
| IL1RN | MI | 14 | 0.194 | 0.031 | 0.133 | 0.255 | 0.000 | 1.214 | 1.142 | 1.291 | 0.000 |
| IL1RN | HF | 14 | -0.002 | 0.031 | -0.063 | 0.059 | 0.956 | 0.998 | 0.939 | 1.061 | 0.999 |
| IL1RN | AF | 14 | 0.013 | 0.025 | -0.036 | 0.061 | 0.616 | 1.013 | 0.964 | 1.063 | 0.912 |
| IL1RN | Stroke | 10 | -0.023 | 0.053 | -0.126 | 0.080 | 0.662 | 0.977 | 0.881 | 1.084 | 0.928 |
| IL23R | COPD | 2 | -0.005 | 0.007 | -0.020 | 0.010 | 0.501 | 0.995 | 0.981 | 1.010 | 0.867 |
| IL23R | Asthma | 2 | -0.004 | 0.022 | -0.046 | 0.038 | 0.840 | 0.996 | 0.955 | 1.039 | 0.971 |
| IL23R | CAD | 2 | -0.006 | 0.018 | -0.042 | 0.029 | 0.725 | 0.994 | 0.959 | 1.030 | 0.947 |
| IL23R | MI | 2 | -0.031 | 0.028 | -0.085 | 0.023 | 0.260 | 0.969 | 0.918 | 1.023 | 0.677 |
| IL23R | HF | 2 | 0.020 | 0.028 | -0.036 | 0.075 | 0.489 | 1.020 | 0.965 | 1.078 | 0.855 |
| IL23R | AF | 2 | 0.014 | 0.024 | -0.033 | 0.061 | 0.559 | 1.014 | 0.967 | 1.063 | 0.896 |
| IL23R | Stroke | 2 | -0.010 | 0.025 | -0.060 | 0.039 | 0.681 | 0.990 | 0.942 | 1.040 | 0.932 |
| IL27RA | COPD | 7 | 0.002 | 0.003 | -0.003 | 0.007 | 0.465 | 1.002 | 0.997 | 1.007 | 0.838 |
| IL27RA | Asthma | 7 | 0.002 | 0.008 | -0.013 | 0.017 | 0.793 | 1.002 | 0.987 | 1.018 | 0.971 |
| IL27RA | CAD | 7 | -0.004 | 0.007 | -0.018 | 0.010 | 0.568 | 0.996 | 0.982 | 1.010 | 0.896 |
| IL27RA | MI | 7 | -0.015 | 0.010 | -0.035 | 0.005 | 0.133 | 0.985 | 0.965 | 1.005 | 0.521 |
| IL27RA | HF | 7 | -0.020 | 0.013 | -0.045 | 0.006 | 0.129 | 0.981 | 0.956 | 1.006 | 0.518 |
| IL27RA | AF | 7 | -0.013 | 0.011 | -0.034 | 0.007 | 0.207 | 0.987 | 0.967 | 1.007 | 0.615 |
| IL27RA | Stroke | 6 | -0.014 | 0.010 | -0.033 | 0.005 | 0.154 | 0.986 | 0.968 | 1.005 | 0.554 |
| IL2RA | COPD | 3 | -0.015 | 0.011 | -0.037 | 0.007 | 0.189 | 0.985 | 0.964 | 1.007 | 0.605 |
| IL2RA | Asthma | 3 | -0.031 | 0.033 | -0.095 | 0.033 | 0.343 | 0.970 | 0.910 | 1.033 | 0.752 |
| IL2RA | CAD | 3 | -0.004 | 0.027 | -0.057 | 0.049 | 0.891 | 0.996 | 0.945 | 1.051 | 0.976 |
| IL2RA | MI | 3 | -0.018 | 0.041 | -0.099 | 0.063 | 0.668 | 0.982 | 0.906 | 1.065 | 0.932 |
| IL2RA | HF | 3 | -0.083 | 0.043 | -0.167 | 0.002 | 0.054 | 0.921 | 0.846 | 1.002 | 0.321 |
| IL2RA | AF | 3 | 0.023 | 0.036 | -0.047 | 0.094 | 0.517 | 1.024 | 0.954 | 1.099 | 0.872 |
| IL2RA | Stroke | 3 | -0.017 | 0.039 | -0.093 | 0.060 | 0.666 | 0.983 | 0.911 | 1.061 | 0.930 |
| IL2RB | COPD | 1 | -0.032 | 0.021 | -0.074 | 0.010 | 0.133 | 0.968 | 0.928 | 1.010 | 0.521 |
| IL2RB | Asthma | 1 | -0.285 | 0.061 | -0.405 | -0.165 | 0.000 | 0.752 | 0.667 | 0.848 | 0.000 |
| IL2RB | CAD | 1 | 0.069 | 0.050 | -0.029 | 0.167 | 0.169 | 1.071 | 0.971 | 1.182 | 0.580 |
| IL2RB | MI | 1 | 0.016 | 0.077 | -0.134 | 0.166 | 0.833 | 1.016 | 0.874 | 1.181 | 0.971 |
| IL2RB | HF | 1 | -0.080 | 0.080 | -0.237 | 0.077 | 0.317 | 0.923 | 0.789 | 1.080 | 0.731 |
| IL2RB | AF | 1 | 0.100 | 0.068 | -0.033 | 0.233 | 0.140 | 1.106 | 0.968 | 1.263 | 0.531 |
| IL2RB | Stroke | 1 | -0.059 | 0.073 | -0.202 | 0.084 | 0.420 | 0.943 | 0.817 | 1.088 | 0.812 |
| IL6R | COPD | 25 | 0.000 | 0.003 | -0.006 | 0.006 | 0.990 | 1.000 | 0.994 | 1.006 | 1.000 |
| IL6R | Asthma | 25 | 0.029 | 0.010 | 0.010 | 0.048 | 0.002 | 1.029 | 1.010 | 1.049 | 0.040 |
| IL6R | CAD | 25 | -0.043 | 0.006 | -0.054 | -0.032 | 0.000 | 0.958 | 0.947 | 0.968 | 0.000 |
| IL6R | MI | 24 | -0.038 | 0.007 | -0.052 | -0.024 | 0.000 | 0.963 | 0.949 | 0.976 | 0.000 |
| IL6R | HF | 24 | -0.011 | 0.010 | -0.030 | 0.007 | 0.235 | 0.989 | 0.971 | 1.007 | 0.649 |
| IL6R | AF | 25 | -0.044 | 0.008 | -0.059 | -0.030 | 0.000 | 0.957 | 0.943 | 0.971 | 0.000 |
| IL6R | Stroke | 23 | -0.023 | 0.006 | -0.035 | -0.011 | 0.000 | 0.977 | 0.965 | 0.989 | 0.006 |
| IL6ST | COPD | 13 | 0.014 | 0.006 | 0.003 | 0.025 | 0.013 | 1.014 | 1.003 | 1.025 | 0.135 |
| IL6ST | Asthma | 13 | 0.054 | 0.015 | 0.025 | 0.082 | 0.000 | 1.055 | 1.025 | 1.086 | 0.007 |
| IL6ST | CAD | 13 | 0.023 | 0.017 | -0.010 | 0.056 | 0.165 | 1.023 | 0.990 | 1.057 | 0.576 |
| IL6ST | MI | 12 | 0.049 | 0.019 | 0.011 | 0.087 | 0.011 | 1.050 | 1.011 | 1.091 | 0.121 |
| IL6ST | HF | 12 | 0.015 | 0.020 | -0.024 | 0.054 | 0.458 | 1.015 | 0.976 | 1.055 | 0.836 |
| IL6ST | AF | 13 | 0.052 | 0.017 | 0.019 | 0.085 | 0.002 | 1.053 | 1.019 | 1.089 | 0.035 |
| IL6ST | Stroke | 11 | -0.010 | 0.018 | -0.045 | 0.025 | 0.574 | 0.990 | 0.956 | 1.025 | 0.896 |
| IL7R | COPD | 7 | 0.025 | 0.008 | 0.010 | 0.041 | 0.001 | 1.026 | 1.010 | 1.042 | 0.023 |
| IL7R | Asthma | 7 | 0.191 | 0.054 | 0.086 | 0.297 | 0.000 | 1.211 | 1.090 | 1.346 | 0.010 |
| IL7R | CAD | 7 | 0.006 | 0.019 | -0.030 | 0.043 | 0.738 | 1.006 | 0.970 | 1.044 | 0.953 |
| IL7R | MI | 7 | 0.024 | 0.028 | -0.032 | 0.079 | 0.403 | 1.024 | 0.969 | 1.082 | 0.800 |
| IL7R | HF | 7 | -0.030 | 0.030 | -0.088 | 0.028 | 0.315 | 0.971 | 0.916 | 1.029 | 0.731 |
| IL7R | AF | 7 | -0.057 | 0.031 | -0.117 | 0.003 | 0.064 | 0.945 | 0.890 | 1.003 | 0.352 |
| IL7R | Stroke | 7 | -0.001 | 0.029 | -0.058 | 0.057 | 0.979 | 0.999 | 0.943 | 1.059 | 1.000 |
| MBL2 | COPD | 18 | 0.000 | 0.002 | -0.003 | 0.004 | 0.762 | 1.000 | 0.997 | 1.004 | 0.965 |
| MBL2 | Asthma | 18 | -0.003 | 0.004 | -0.012 | 0.005 | 0.426 | 0.997 | 0.988 | 1.005 | 0.815 |
| MBL2 | CAD | 18 | -0.004 | 0.004 | -0.011 | 0.003 | 0.300 | 0.996 | 0.989 | 1.003 | 0.723 |
| MBL2 | MI | 17 | 0.000 | 0.006 | -0.011 | 0.011 | 0.991 | 1.000 | 0.989 | 1.011 | 1.000 |
| MBL2 | HF | 18 | -0.009 | 0.009 | -0.026 | 0.008 | 0.296 | 0.991 | 0.974 | 1.008 | 0.718 |
| MBL2 | AF | 19 | -0.011 | 0.005 | -0.021 | -0.002 | 0.023 | 0.989 | 0.979 | 0.998 | 0.195 |
| MBL2 | Stroke | 16 | -0.004 | 0.007 | -0.019 | 0.010 | 0.556 | 0.996 | 0.982 | 1.010 | 0.896 |
| MIF | COPD | 1 | -0.030 | 0.014 | -0.057 | -0.003 | 0.029 | 0.970 | 0.944 | 0.997 | 0.224 |
| MIF | Asthma | 1 | -0.050 | 0.040 | -0.127 | 0.028 | 0.206 | 0.951 | 0.880 | 1.028 | 0.615 |
| MIF | CAD | 1 | 0.140 | 0.032 | 0.077 | 0.204 | 0.000 | 1.151 | 1.080 | 1.226 | 0.001 |
| MIF | MI | 1 | 0.090 | 0.049 | -0.006 | 0.186 | 0.067 | 1.094 | 0.994 | 1.204 | 0.359 |
| MIF | HF | 1 | 0.124 | 0.052 | 0.023 | 0.225 | 0.016 | 1.132 | 1.023 | 1.253 | 0.159 |
| MIF | AF | 1 | -0.020 | 0.044 | -0.106 | 0.066 | 0.648 | 0.980 | 0.899 | 1.068 | 0.924 |
| MIF | Stroke | 1 | -0.003 | 0.047 | -0.096 | 0.089 | 0.945 | 0.997 | 0.909 | 1.093 | 0.996 |
| PGF | COPD | 2 | -0.016 | 0.016 | -0.046 | 0.015 | 0.323 | 0.985 | 0.955 | 1.015 | 0.733 |
| PGF | Asthma | 2 | 0.043 | 0.045 | -0.046 | 0.132 | 0.346 | 1.044 | 0.955 | 1.141 | 0.755 |
| PGF | CAD | 2 | -0.284 | 0.038 | -0.357 | -0.210 | 0.000 | 0.753 | 0.699 | 0.810 | 0.000 |
| PGF | MI | 2 | -0.272 | 0.057 | -0.385 | -0.159 | 0.000 | 0.762 | 0.681 | 0.853 | 0.000 |
| PGF | HF | 2 | 0.019 | 0.059 | -0.097 | 0.135 | 0.746 | 1.019 | 0.908 | 1.145 | 0.956 |
| PGF | AF | 2 | 0.028 | 0.050 | -0.071 | 0.127 | 0.580 | 1.028 | 0.932 | 1.135 | 0.896 |
| PGF | Stroke | 2 | -0.055 | 0.054 | -0.162 | 0.052 | 0.312 | 0.946 | 0.851 | 1.053 | 0.729 |
| SAA1 | COPD | 12 | 0.008 | 0.004 | 0.000 | 0.015 | 0.051 | 1.008 | 1.000 | 1.015 | 0.310 |
| SAA1 | Asthma | 12 | 0.009 | 0.015 | -0.020 | 0.037 | 0.555 | 1.009 | 0.980 | 1.038 | 0.896 |
| SAA1 | CAD | 12 | 0.020 | 0.009 | 0.002 | 0.037 | 0.030 | 1.020 | 1.002 | 1.038 | 0.225 |
| SAA1 | MI | 12 | 0.032 | 0.019 | -0.006 | 0.070 | 0.097 | 1.032 | 0.994 | 1.072 | 0.448 |
| SAA1 | HF | 12 | 0.018 | 0.014 | -0.010 | 0.046 | 0.205 | 1.018 | 0.990 | 1.047 | 0.615 |
| SAA1 | AF | 12 | 0.004 | 0.012 | -0.020 | 0.027 | 0.752 | 1.004 | 0.980 | 1.028 | 0.958 |
| SAA1 | Stroke | 11 | 0.014 | 0.013 | -0.011 | 0.039 | 0.257 | 1.015 | 0.990 | 1.040 | 0.675 |
| SAA2 | COPD | 12 | 0.007 | 0.004 | 0.000 | 0.015 | 0.059 | 1.007 | 1.000 | 1.015 | 0.333 |
| SAA2 | Asthma | 12 | 0.010 | 0.016 | -0.022 | 0.041 | 0.549 | 1.010 | 0.978 | 1.042 | 0.892 |
| SAA2 | CAD | 12 | 0.028 | 0.009 | 0.009 | 0.046 | 0.003 | 1.028 | 1.009 | 1.047 | 0.053 |
| SAA2 | MI | 10 | 0.044 | 0.019 | 0.007 | 0.080 | 0.019 | 1.045 | 1.007 | 1.083 | 0.171 |
| SAA2 | HF | 10 | 0.015 | 0.016 | -0.016 | 0.045 | 0.350 | 1.015 | 0.984 | 1.047 | 0.755 |
| SAA2 | AF | 10 | 0.000 | 0.015 | -0.030 | 0.029 | 0.984 | 1.000 | 0.971 | 1.030 | 1.000 |
| SAA2 | Stroke | 8 | 0.004 | 0.014 | -0.024 | 0.032 | 0.791 | 1.004 | 0.976 | 1.033 | 0.971 |
| SERPINE1 | COPD | 4 | -0.010 | 0.015 | -0.040 | 0.019 | 0.495 | 0.990 | 0.961 | 1.019 | 0.860 |
| SERPINE1 | Asthma | 4 | -0.061 | 0.043 | -0.146 | 0.024 | 0.158 | 0.941 | 0.864 | 1.024 | 0.562 |
| SERPINE1 | CAD | 4 | 0.038 | 0.037 | -0.035 | 0.110 | 0.305 | 1.039 | 0.966 | 1.117 | 0.727 |
| SERPINE1 | MI | 4 | 0.017 | 0.056 | -0.093 | 0.126 | 0.768 | 1.017 | 0.911 | 1.135 | 0.965 |
| SERPINE1 | HF | 4 | -0.067 | 0.063 | -0.191 | 0.058 | 0.293 | 0.936 | 0.826 | 1.059 | 0.718 |
| SERPINE1 | AF | 4 | 0.132 | 0.059 | 0.016 | 0.248 | 0.026 | 1.141 | 1.016 | 1.281 | 0.208 |
| SERPINE1 | Stroke | 4 | 0.140 | 0.057 | 0.029 | 0.251 | 0.013 | 1.151 | 1.030 | 1.286 | 0.136 |
| TNFSF10 | COPD | 4 | 0.010 | 0.017 | -0.023 | 0.043 | 0.562 | 1.010 | 0.977 | 1.044 | 0.896 |
| TNFSF10 | Asthma | 4 | 0.055 | 0.034 | -0.011 | 0.121 | 0.100 | 1.057 | 0.990 | 1.129 | 0.451 |
| TNFSF10 | CAD | 4 | 0.037 | 0.027 | -0.015 | 0.090 | 0.165 | 1.038 | 0.985 | 1.095 | 0.576 |
| TNFSF10 | MI | 4 | 0.060 | 0.042 | -0.022 | 0.143 | 0.152 | 1.062 | 0.978 | 1.154 | 0.550 |
| TNFSF10 | HF | 4 | -0.015 | 0.047 | -0.108 | 0.078 | 0.749 | 0.985 | 0.897 | 1.081 | 0.956 |
| TNFSF10 | AF | 4 | 0.009 | 0.058 | -0.104 | 0.122 | 0.874 | 1.009 | 0.901 | 1.130 | 0.971 |
| TNFSF10 | Stroke | 4 | 0.010 | 0.055 | -0.098 | 0.118 | 0.851 | 1.010 | 0.907 | 1.126 | 0.971 |
| VEGFA | COPD | 18 | 0.011 | 0.005 | 0.001 | 0.021 | 0.024 | 1.011 | 1.001 | 1.021 | 0.198 |
| VEGFA | Asthma | 18 | -0.009 | 0.013 | -0.034 | 0.017 | 0.504 | 0.991 | 0.967 | 1.017 | 0.869 |
| VEGFA | CAD | 18 | -0.003 | 0.011 | -0.025 | 0.020 | 0.821 | 0.997 | 0.975 | 1.020 | 0.971 |
| VEGFA | MI | 17 | -0.011 | 0.015 | -0.041 | 0.018 | 0.444 | 0.989 | 0.960 | 1.018 | 0.824 |
| VEGFA | HF | 18 | 0.029 | 0.017 | -0.004 | 0.063 | 0.087 | 1.030 | 0.996 | 1.065 | 0.418 |
| VEGFA | AF | 18 | 0.009 | 0.015 | -0.021 | 0.039 | 0.556 | 1.009 | 0.979 | 1.040 | 0.896 |
| VEGFA | Stroke | 18 | 0.016 | 0.016 | -0.015 | 0.048 | 0.315 | 1.016 | 0.985 | 1.049 | 0.731 |
| VEGFC | COPD | 1 | -0.025 | 0.016 | -0.055 | 0.006 | 0.110 | 0.975 | 0.946 | 1.006 | 0.475 |
| VEGFC | Asthma | 1 | 0.007 | 0.044 | -0.080 | 0.094 | 0.878 | 1.007 | 0.923 | 1.099 | 0.971 |
| VEGFC | CAD | 1 | 0.011 | 0.039 | -0.065 | 0.087 | 0.773 | 1.011 | 0.937 | 1.091 | 0.965 |
| VEGFC | MI | 1 | -0.016 | 0.061 | -0.135 | 0.103 | 0.794 | 0.984 | 0.874 | 1.109 | 0.971 |
| VEGFC | HF | 1 | -0.042 | 0.063 | -0.166 | 0.082 | 0.509 | 0.959 | 0.847 | 1.086 | 0.870 |
| VEGFC | AF | 1 | 0.010 | 0.053 | -0.093 | 0.114 | 0.845 | 1.010 | 0.911 | 1.121 | 0.971 |
| VEGFC | Stroke | 1 | 0.081 | 0.058 | -0.034 | 0.195 | 0.167 | 1.084 | 0.967 | 1.215 | 0.576 |

Blue highlight denotes suggestive evidence based on FDR-corrected p-value (q-value).

Orange highlight denotes strong evidence based on FDR-corrected p-value (q-value).

CI95Lower: 95% lower CI for the estimate; Ci95Upper: 95% upper CI for the estimate.

ORlow: 95% lower CI for the OR; ORhi: 95% upper CI for the OR.

AF: atrial fibrillation, CAD: coronary artery disease, COPD: chronic obstructive pulmonary diseases, HF: heart failure, IS: ischemia stroke, MI: myocardial infarction,

n: number of SNPs used as instrument variables in each method, SNPs single nucleotide polymorphism, CI confidence intervals, OR odds ratio.

**Supplement Table 8. Cis-MR IVW and sensitivity analyses results for inflammatory markers that associated with both chronic respiratory diseases and cardiovascular diseases.**

| **Exposure** | **Outcome** | **n** | **Method** | **Estimate** | **StdError** | **CI95Lower** | **CI95Upper** | **p-value** | **OR** | **ORlow** | **ORhi** | **q-value** |
| --- | --- | --- | --- | --- | --- | --- | --- | --- | --- | --- | --- | --- |
| IL6R | COPD | 25 | IVW | 0.000 | 0.003 | -0.006 | 0.006 | 0.990 | 1.000 | 0.994 | 1.006 | 1.000 |
|  |  | 25 | Weighted median | 0.003 | 0.003 | -0.002 | 0.008 | 0.265 | 1.003 | 0.998 | 1.008 | 0.685 |
|  |  | 25 | MR-Egger | -0.001 | 0.006 | -0.013 | 0.011 | 0.866 | 0.999 | 0.987 | 1.011 | 0.971 |
|  |  | 22 | MR-PRESSO | 0.003 | 0.002 | -0.001 | 0.008 | 0.174 | 1.003 | 0.999 | 1.008 | 0.593 |
|  |  | 25 | Contamination mixture | 0.003 | 0.000 | 0.003 | 0.003 | 0.085 | 1.003 | 1.003 | 1.003 | 0.410 |
|  | Asthma | 25 | IVW | 0.029 | 0.010 | 0.010 | 0.048 | 0.002 | 1.029 | 1.010 | 1.049 | 0.040 |
|  |  | 25 | Weighted median | 0.039 | 0.009 | 0.023 | 0.056 | 0.000 | 1.040 | 1.023 | 1.058 | 0.000 |
|  |  | 25 | MR-Egger | 0.017 | 0.019 | -0.020 | 0.055 | 0.364 | 1.017 | 0.980 | 1.056 | 0.767 |
|  |  | 23 | MR-PRESSO | 0.032 | 0.008 | 0.016 | 0.049 | 0.001 | 1.033 | 1.016 | 1.050 | 0.020 |
|  |  | 25 | Contamination mixture | 0.054 | 0.003 | -0.006 | 0.004 | 0.098 | 1.055 | 0.994 | 1.004 | 0.450 |
|  |  | 25 | Contamination mixture | 0.054 | 0.010 | 0.024 | 0.064 | 0.098 | 1.055 | 1.024 | 1.066 | 0.450 |
|  | CAD | 25 | IVW | -0.043 | 0.006 | -0.054 | -0.032 | 0.000 | 0.958 | 0.947 | 0.968 | 0.000 |
|  |  | 25 | Weighted median | -0.045 | 0.006 | -0.057 | -0.034 | 0.000 | 0.956 | 0.945 | 0.967 | 0.000 |
|  |  | 25 | MR-Egger | -0.052 | 0.011 | -0.074 | -0.030 | 0.000 | 0.950 | 0.929 | 0.971 | 0.000 |
|  |  | 25 | MR-PRESSO | -0.043 | 0.006 | -0.054 | -0.032 | 0.000 | 0.958 | 0.947 | 0.968 | 0.000 |
|  |  | 25 | Contamination mixture | -0.050 | 0.005 | -0.060 | -0.040 | 0.000 | 0.951 | 0.942 | 0.961 | 0.000 |
|  | MI | 24 | IVW | -0.038 | 0.007 | -0.052 | -0.024 | 0.000 | 0.963 | 0.949 | 0.976 | 0.000 |
|  |  | 24 | Weighted median | -0.046 | 0.009 | -0.063 | -0.030 | 0.000 | 0.955 | 0.939 | 0.971 | 0.000 |
|  |  | 24 | MR-Egger | -0.047 | 0.014 | -0.076 | -0.019 | 0.001 | 0.954 | 0.927 | 0.981 | 0.022 |
|  |  | 24 | MR-PRESSO | -0.038 | 0.007 | -0.052 | -0.024 | 0.000 | 0.963 | 0.949 | 0.976 | 0.001 |
|  |  | 24 | Contamination mixture | -0.046 | 0.005 | -0.056 | -0.036 | 0.000 | 0.955 | 0.946 | 0.965 | 0.000 |
|  | HF | 24 | IVW | -0.011 | 0.010 | -0.030 | 0.007 | 0.235 | 0.989 | 0.971 | 1.007 | 0.649 |
|  |  | 24 | Weighted median | -0.014 | 0.009 | -0.032 | 0.005 | 0.149 | 0.987 | 0.969 | 1.005 | 0.548 |
|  |  | 24 | MR-Egger | -0.029 | 0.019 | -0.067 | 0.009 | 0.136 | 0.971 | 0.935 | 1.009 | 0.527 |
|  |  | 23 | MR-PRESSO | -0.010 | 0.009 | -0.027 | 0.007 | 0.245 | 0.990 | 0.973 | 1.007 | 0.658 |
|  |  | 24 | Contamination mixture | -0.016 | 0.005 | -0.026 | -0.006 | 0.025 | 0.984 | 0.974 | 0.994 | 0.203 |
|  | AF | 25 | IVW | -0.044 | 0.008 | -0.059 | -0.030 | 0.000 | 0.957 | 0.943 | 0.971 | 0.000 |
|  |  | 25 | Weighted median | -0.047 | 0.008 | -0.062 | -0.032 | 0.000 | 0.954 | 0.940 | 0.969 | 0.000 |
|  |  | 25 | MR-Egger | -0.068 | 0.015 | -0.097 | -0.039 | 0.000 | 0.934 | 0.908 | 0.962 | 0.000 |
|  |  | 24 | MR-PRESSO | -0.041 | 0.007 | -0.055 | -0.028 | 0.000 | 0.960 | 0.947 | 0.972 | 0.000 |
|  |  | 25 | Contamination mixture | -0.052 | 0.005 | -0.062 | -0.042 | 0.000 | 0.949 | 0.940 | 0.959 | 0.010 |
|  | Stroke | 23 | IVW | -0.023 | 0.006 | -0.035 | -0.011 | 0.000 | 0.977 | 0.965 | 0.989 | 0.006 |
|  |  | 23 | Weighted median | -0.021 | 0.009 | -0.038 | -0.003 | 0.020 | 0.980 | 0.963 | 0.997 | 0.175 |
|  |  | 23 | MR-Egger | -0.016 | 0.013 | -0.043 | 0.010 | 0.225 | 0.984 | 0.958 | 1.010 | 0.638 |
|  |  | 23 | MR-PRESSO | -0.023 | 0.006 | -0.035 | -0.011 | 0.001 | 0.977 | 0.965 | 0.989 | 0.023 |
|  |  | 23 | Contamination mixture | -0.017 | 0.005 | -0.027 | -0.007 | 0.045 | 0.983 | 0.973 | 0.993 | 0.287 |
| IL1RN | COPD | 13 | IVW | -0.028 | 0.009 | -0.046 | -0.011 | 0.001 | 0.972 | 0.955 | 0.989 | 0.026 |
|  |  | 13 | Weighted median | -0.032 | 0.009 | -0.050 | -0.015 | 0.000 | 0.968 | 0.951 | 0.985 | 0.009 |
|  |  | 13 | MR-Egger | -0.053 | 0.019 | -0.090 | -0.016 | 0.005 | 0.948 | 0.914 | 0.984 | 0.072 |
|  |  | 13 | MR-PRESSO | -0.028 | 0.009 | -0.046 | -0.011 | 0.008 | 0.972 | 0.955 | 0.989 | 0.090 |
|  |  | 13 | Contamination mixture | -0.028 | 0.008 | -0.048 | -0.018 | 0.027 | 0.972 | 0.953 | 0.982 | 0.210 |
|  | Asthma | 14 | IVW | -0.055 | 0.023 | -0.100 | -0.009 | 0.019 | 0.947 | 0.905 | 0.991 | 0.171 |
|  |  | 14 | Weighted median | -0.061 | 0.025 | -0.111 | -0.011 | 0.017 | 0.941 | 0.895 | 0.989 | 0.163 |
|  |  | 14 | MR-Egger | -0.044 | 0.055 | -0.152 | 0.064 | 0.421 | 0.957 | 0.859 | 1.066 | 0.812 |
|  |  | 14 | MR-PRESSO | -0.055 | 0.023 | -0.100 | -0.009 | 0.035 | 0.947 | 0.905 | 0.991 | 0.247 |
|  |  | 14 | Contamination mixture | -0.097 | 0.038 | -0.177 | -0.027 | 0.009 | 0.907 | 0.838 | 0.973 | 0.105 |
|  | CAD | 14 | IVW | 0.083 | 0.019 | 0.045 | 0.121 | 0.000 | 1.087 | 1.046 | 1.129 | 0.001 |
|  |  | 14 | Weighted median | 0.082 | 0.023 | 0.037 | 0.127 | 0.000 | 1.085 | 1.038 | 1.135 | 0.010 |
|  |  | 14 | MR-Egger | 0.074 | 0.045 | -0.014 | 0.162 | 0.100 | 1.077 | 0.986 | 1.176 | 0.451 |
|  |  | 14 | MR-PRESSO | 0.083 | 0.019 | 0.045 | 0.121 | 0.001 | 1.087 | 1.046 | 1.129 | 0.020 |
|  |  | 14 | Contamination mixture | 0.111 | 0.036 | 0.061 | 0.201 | 0.001 | 1.117 | 1.063 | 1.222 | 0.016 |
|  | MI | 14 | IVW | 0.194 | 0.031 | 0.133 | 0.255 | 0.000 | 1.214 | 1.142 | 1.291 | 0.000 |
|  |  | 14 | Weighted median | 0.190 | 0.033 | 0.125 | 0.255 | 0.000 | 1.209 | 1.133 | 1.291 | 0.000 |
|  |  | 14 | MR-Egger | 0.177 | 0.073 | 0.034 | 0.320 | 0.015 | 1.194 | 1.035 | 1.377 | 0.152 |
|  |  | 14 | MR-PRESSO | 0.194 | 0.031 | 0.133 | 0.255 | 0.000 | 1.214 | 1.142 | 1.291 | 0.001 |
|  |  | 14 | Contamination mixture | 0.188 | 0.033 | 0.148 | 0.278 | 0.000 | 1.207 | 1.160 | 1.321 | 0.001 |
|  | HF | 14 | IVW | -0.002 | 0.031 | -0.063 | 0.059 | 0.956 | 0.998 | 0.939 | 1.061 | 0.999 |
|  |  | 14 | Weighted median | 0.043 | 0.043 | -0.041 | 0.127 | 0.310 | 1.044 | 0.960 | 1.136 | 0.727 |
|  |  | 14 | MR-Egger | 0.062 | 0.067 | -0.070 | 0.194 | 0.356 | 1.064 | 0.933 | 1.214 | 0.765 |
|  |  | 14 | MR-PRESSO | -0.002 | 0.031 | -0.063 | 0.059 | 0.957 | 0.998 | 0.939 | 1.061 | 0.999 |
|  |  | 14 | Contamination mixture | 0.060 | 0.041 | -0.040 | 0.120 | 0.360 | 1.062 | 0.961 | 1.127 | 0.767 |
|  | AF | 14 | IVW | 0.013 | 0.025 | -0.036 | 0.061 | 0.616 | 1.013 | 0.964 | 1.063 | 0.912 |
|  |  | 14 | Weighted median | -0.005 | 0.033 | -0.069 | 0.060 | 0.881 | 0.995 | 0.933 | 1.061 | 0.973 |
|  |  | 14 | MR-Egger | -0.007 | 0.054 | -0.113 | 0.099 | 0.893 | 0.993 | 0.893 | 1.104 | 0.976 |
|  |  | 14 | MR-PRESSO | 0.013 | 0.020 | -0.027 | 0.052 | 0.547 | 1.013 | 0.973 | 1.054 | 0.891 |
|  |  | 14 | Contamination mixture | 0.015 | 0.036 | -0.045 | 0.095 | 0.591 | 1.015 | 0.956 | 1.099 | 0.899 |
|  | Stroke | 10 | IVW | -0.023 | 0.053 | -0.126 | 0.080 | 0.662 | 0.977 | 0.881 | 1.084 | 0.928 |
|  |  | 10 | Weighted median | -0.029 | 0.058 | -0.142 | 0.084 | 0.619 | 0.972 | 0.868 | 1.088 | 0.912 |
|  |  | 10 | MR-Egger | 0.032 | 0.125 | -0.213 | 0.278 | 0.797 | 1.033 | 0.808 | 1.321 | 0.971 |
|  |  | 10 | MR-PRESSO | -0.023 | 0.053 | -0.126 | 0.080 | 0.673 | 0.977 | 0.881 | 1.084 | 0.932 |
|  |  | 10 | Contamination mixture | 0.045 | 0.153 | -0.355 | 0.245 | 0.348 | 1.046 | 0.701 | 1.277 | 0.755 |
| CXCL6 | COPD | 12 | IVW | -0.014 | 0.004 | -0.021 | -0.007 | 0.000 | 0.986 | 0.979 | 0.994 | 0.006 |
|  |  | 12 | Weighted median | -0.010 | 0.004 | -0.018 | -0.001 | 0.033 | 0.990 | 0.982 | 0.999 | 0.233 |
|  |  | 12 | MR-Egger | -0.002 | 0.009 | -0.020 | 0.016 | 0.841 | 0.998 | 0.980 | 1.016 | 0.971 |
|  |  | 12 | MR-PRESSO | -0.014 | 0.004 | -0.021 | -0.007 | 0.003 | 0.986 | 0.979 | 0.994 | 0.051 |
|  |  | 12 | Contamination mixture | -0.025 | 0.008 | -0.035 | -0.005 | 0.019 | 0.975 | 0.965 | 0.995 | 0.171 |
|  | Asthma | 12 | IVW | -0.003 | 0.014 | -0.030 | 0.025 | 0.845 | 0.997 | 0.970 | 1.025 | 0.971 |
|  |  | 12 | Weighted median | 0.011 | 0.013 | -0.015 | 0.036 | 0.415 | 1.011 | 0.985 | 1.037 | 0.808 |
|  |  | 12 | MR-Egger | 0.055 | 0.033 | -0.011 | 0.120 | 0.101 | 1.056 | 0.989 | 1.128 | 0.455 |
|  |  | 12 | MR-PRESSO | -0.003 | 0.014 | -0.030 | 0.025 | 0.849 | 0.997 | 0.970 | 1.025 | 0.971 |
|  |  | 12 | Contamination mixture | -0.073 | 0.020 | -0.113 | -0.033 | 0.056 | 0.930 | 0.893 | 0.968 | 0.321 |
|  |  | 12 | Contamination mixture | -0.073 | 0.013 | 0.007 | 0.057 | 0.056 | 0.930 | 1.007 | 1.059 | 0.321 |
|  | CAD | 12 | IVW | 0.012 | 0.010 | -0.008 | 0.032 | 0.238 | 1.012 | 0.992 | 1.032 | 0.653 |
|  |  | 12 | Weighted median | 0.019 | 0.011 | -0.003 | 0.041 | 0.095 | 1.019 | 0.997 | 1.041 | 0.445 |
|  |  | 12 | MR-Egger | 0.004 | 0.027 | -0.050 | 0.057 | 0.895 | 1.004 | 0.951 | 1.059 | 0.978 |
|  |  | 12 | MR-PRESSO | 0.012 | 0.010 | -0.008 | 0.032 | 0.263 | 1.012 | 0.992 | 1.032 | 0.684 |
|  |  | 12 | Contamination mixture | 0.028 | 0.015 | 0.008 | 0.068 | 0.020 | 1.028 | 1.008 | 1.070 | 0.173 |
|  | MI | 12 | IVW | 0.031 | 0.013 | 0.006 | 0.055 | 0.014 | 1.031 | 1.006 | 1.057 | 0.140 |
|  |  | 12 | Weighted median | 0.041 | 0.017 | 0.009 | 0.074 | 0.013 | 1.042 | 1.009 | 1.077 | 0.134 |
|  |  | 12 | MR-Egger | 0.036 | 0.033 | -0.028 | 0.099 | 0.273 | 1.036 | 0.972 | 1.105 | 0.695 |
|  |  | 12 | MR-PRESSO | 0.031 | 0.011 | 0.008 | 0.053 | 0.021 | 1.031 | 1.008 | 1.055 | 0.179 |
|  |  | 12 | Contamination mixture | 0.046 | 0.013 | 0.026 | 0.076 | 0.017 | 1.047 | 1.026 | 1.079 | 0.163 |
|  | HF | 12 | IVW | 0.034 | 0.013 | 0.008 | 0.059 | 0.010 | 1.034 | 1.008 | 1.061 | 0.110 |
|  |  | 12 | Weighted median | 0.032 | 0.016 | -0.001 | 0.064 | 0.055 | 1.032 | 0.999 | 1.066 | 0.321 |
|  |  | 12 | MR-Egger | 0.021 | 0.034 | -0.046 | 0.087 | 0.544 | 1.021 | 0.955 | 1.091 | 0.891 |
|  |  | 12 | MR-PRESSO | 0.034 | 0.009 | 0.017 | 0.051 | 0.003 | 1.034 | 1.017 | 1.052 | 0.041 |
|  |  | 12 | Contamination mixture | 0.040 | 0.015 | 0.010 | 0.070 | 0.010 | 1.041 | 1.010 | 1.073 | 0.110 |
|  | AF | 12 | IVW | 0.041 | 0.013 | 0.016 | 0.065 | 0.001 | 1.042 | 1.016 | 1.068 | 0.022 |
|  |  | 12 | Weighted median | 0.039 | 0.014 | 0.011 | 0.067 | 0.006 | 1.040 | 1.011 | 1.069 | 0.077 |
|  |  | 12 | MR-Egger | 0.022 | 0.033 | -0.044 | 0.088 | 0.510 | 1.022 | 0.957 | 1.091 | 0.870 |
|  |  | 12 | MR-PRESSO | 0.041 | 0.013 | 0.016 | 0.065 | 0.008 | 1.042 | 1.016 | 1.068 | 0.090 |
|  |  | 12 | Contamination mixture | 0.044 | 0.010 | 0.024 | 0.064 | 0.005 | 1.045 | 1.024 | 1.066 | 0.068 |
|  | Stroke | 12 | IVW | 0.005 | 0.012 | -0.018 | 0.028 | 0.681 | 1.005 | 0.982 | 1.029 | 0.932 |
|  |  | 12 | Weighted median | 0.010 | 0.015 | -0.019 | 0.040 | 0.482 | 1.011 | 0.981 | 1.040 | 0.850 |
|  |  | 12 | MR-Egger | 0.022 | 0.031 | -0.038 | 0.083 | 0.469 | 1.023 | 0.962 | 1.087 | 0.838 |
|  |  | 12 | MR-PRESSO | 0.005 | 0.008 | -0.012 | 0.022 | 0.576 | 1.005 | 0.988 | 1.022 | 0.896 |
|  |  | 12 | Contamination mixture | 0.013 | 0.020 | -0.047 | 0.033 | 0.361 | 1.013 | 0.954 | 1.033 | 0.767 |
| IL1RL2 | COPD | 15 | IVW | -0.026 | 0.009 | -0.043 | -0.008 | 0.004 | 0.975 | 0.957 | 0.992 | 0.062 |
|  |  | 15 | Weighted median | -0.033 | 0.008 | -0.049 | -0.018 | 0.000 | 0.967 | 0.953 | 0.982 | 0.001 |
|  |  | 15 | MR-Egger | -0.003 | 0.019 | -0.041 | 0.035 | 0.876 | 0.997 | 0.960 | 1.035 | 0.971 |
|  |  | 12 | MR-PRESSO | -0.036 | 0.006 | -0.049 | -0.024 | 0.000 | 0.964 | 0.953 | 0.976 | 0.004 |
|  |  | 15 | Contamination mixture | -0.044 | 0.005 | -0.054 | -0.034 | 0.003 | 0.957 | 0.948 | 0.967 | 0.041 |
|  | Asthma | 15 | IVW | -0.070 | 0.043 | -0.154 | 0.014 | 0.104 | 0.932 | 0.857 | 1.014 | 0.462 |
|  |  | 15 | Weighted median | -0.063 | 0.031 | -0.123 | -0.002 | 0.043 | 0.939 | 0.884 | 0.998 | 0.284 |
|  |  | 15 | MR-Egger | -0.113 | 0.098 | -0.305 | 0.079 | 0.248 | 0.893 | 0.737 | 1.082 | 0.662 |
|  |  | 9 | MR-PRESSO | -0.072 | 0.035 | -0.140 | -0.003 | 0.076 | 0.931 | 0.869 | 0.997 | 0.386 |
|  |  | 15 | Contamination mixture | -0.021 | 0.015 | -0.241 | -0.181 | 0.295 | 0.979 | 0.786 | 0.835 | 0.718 |
|  |  | 15 | Contamination mixture | -0.021 | 0.071 | -0.141 | 0.139 | 0.295 | 0.979 | 0.869 | 1.149 | 0.718 |
|  | CAD | 15 | IVW | -0.048 | 0.014 | -0.076 | -0.020 | 0.001 | 0.953 | 0.927 | 0.980 | 0.017 |
|  |  | 15 | Weighted median | -0.053 | 0.015 | -0.083 | -0.023 | 0.000 | 0.948 | 0.920 | 0.977 | 0.012 |
|  |  | 15 | MR-Egger | -0.069 | 0.032 | -0.133 | -0.006 | 0.032 | 0.933 | 0.876 | 0.994 | 0.233 |
|  |  | 15 | MR-PRESSO | -0.048 | 0.014 | -0.076 | -0.020 | 0.005 | 0.953 | 0.927 | 0.980 | 0.066 |
|  |  | 15 | Contamination mixture | -0.046 | 0.010 | -0.066 | -0.026 | 0.005 | 0.955 | 0.936 | 0.974 | 0.068 |
|  | MI | 15 | IVW | -0.040 | 0.020 | -0.079 | -0.002 | 0.041 | 0.961 | 0.924 | 0.998 | 0.276 |
|  |  | 15 | Weighted median | -0.046 | 0.023 | -0.091 | -0.001 | 0.047 | 0.955 | 0.913 | 0.999 | 0.298 |
|  |  | 15 | MR-Egger | -0.066 | 0.045 | -0.155 | 0.022 | 0.142 | 0.936 | 0.856 | 1.022 | 0.535 |
|  |  | 15 | MR-PRESSO | -0.040 | 0.020 | -0.079 | -0.002 | 0.060 | 0.961 | 0.924 | 0.998 | 0.337 |
|  |  | 15 | Contamination mixture | -0.062 | 0.015 | -0.092 | -0.032 | 0.005 | 0.940 | 0.912 | 0.968 | 0.068 |
|  | HF | 15 | IVW | 0.005 | 0.017 | -0.030 | 0.039 | 0.794 | 1.005 | 0.971 | 1.039 | 0.971 |
|  |  | 15 | Weighted median | 0.006 | 0.023 | -0.040 | 0.052 | 0.802 | 1.006 | 0.961 | 1.053 | 0.971 |
|  |  | 15 | MR-Egger | -0.041 | 0.038 | -0.116 | 0.033 | 0.280 | 0.960 | 0.891 | 1.034 | 0.703 |
|  |  | 15 | MR-PRESSO | 0.005 | 0.015 | -0.024 | 0.033 | 0.762 | 1.005 | 0.976 | 1.034 | 0.965 |
|  |  | 15 | Contamination mixture | 0.001 | 0.026 | -0.049 | 0.051 | 0.860 | 1.001 | 0.953 | 1.053 | 0.971 |
|  | AF | 16 | IVW | -0.004 | 0.015 | -0.033 | 0.026 | 0.800 | 0.996 | 0.967 | 1.026 | 0.971 |
|  |  | 16 | Weighted median | -0.011 | 0.022 | -0.054 | 0.033 | 0.625 | 0.989 | 0.947 | 1.033 | 0.912 |
|  |  | 16 | MR-Egger | -0.007 | 0.034 | -0.073 | 0.059 | 0.832 | 0.993 | 0.929 | 1.061 | 0.971 |
|  |  | 16 | MR-PRESSO | -0.004 | 0.014 | -0.032 | 0.024 | 0.796 | 0.996 | 0.968 | 1.025 | 0.971 |
|  |  | 16 | Contamination mixture | -0.038 | 0.031 | -0.098 | 0.022 | 0.212 | 0.963 | 0.907 | 1.022 | 0.623 |
|  | Stroke | 15 | IVW | -0.001 | 0.015 | -0.032 | 0.029 | 0.933 | 0.999 | 0.969 | 1.029 | 0.993 |
|  |  | 15 | Weighted median | -0.002 | 0.021 | -0.042 | 0.039 | 0.941 | 0.998 | 0.959 | 1.040 | 0.995 |
|  |  | 15 | MR-Egger | -0.020 | 0.035 | -0.087 | 0.048 | 0.567 | 0.980 | 0.916 | 1.049 | 0.896 |
|  |  | 15 | MR-PRESSO | -0.001 | 0.015 | -0.031 | 0.029 | 0.934 | 0.999 | 0.969 | 1.029 | 0.993 |
|  |  | 15 | Contamination mixture | 0.005 | 0.018 | -0.035 | 0.035 | 1.000 | 1.005 | 0.966 | 1.036 | 1.000 |
| HP | COPD | 13 | IVW | -0.006 | 0.002 | -0.010 | -0.002 | 0.003 | 0.994 | 0.990 | 0.998 | 0.048 |
|  |  | 13 | Weighted median | -0.004 | 0.003 | -0.009 | 0.001 | 0.112 | 0.996 | 0.991 | 1.001 | 0.482 |
|  |  | 13 | MR-Egger | -0.004 | 0.006 | -0.016 | 0.008 | 0.522 | 0.996 | 0.985 | 1.008 | 0.877 |
|  |  | 13 | MR-PRESSO | -0.006 | 0.002 | -0.010 | -0.002 | 0.012 | 0.994 | 0.990 | 0.998 | 0.126 |
|  |  | 13 | Contamination mixture | -0.010 | 0.003 | -0.010 | 0.000 | 0.572 | 0.990 | 0.990 | 1.000 | 0.896 |
|  | Asthma | 13 | IVW | -0.017 | 0.006 | -0.028 | -0.005 | 0.005 | 0.984 | 0.972 | 0.995 | 0.068 |
|  |  | 13 | Weighted median | -0.021 | 0.007 | -0.034 | -0.008 | 0.001 | 0.979 | 0.966 | 0.992 | 0.023 |
|  |  | 13 | MR-Egger | -0.043 | 0.014 | -0.071 | -0.014 | 0.003 | 0.958 | 0.931 | 0.986 | 0.049 |
|  |  | 13 | MR-PRESSO | -0.017 | 0.006 | -0.028 | -0.005 | 0.016 | 0.984 | 0.972 | 0.995 | 0.155 |
|  |  | 13 | Contamination mixture | -0.025 | 0.003 | -0.025 | -0.015 | 0.006 | 0.976 | 0.976 | 0.986 | 0.080 |
|  | CAD | 12 | IVW | -0.020 | 0.008 | -0.035 | -0.004 | 0.016 | 0.981 | 0.965 | 0.996 | 0.159 |
|  |  | 12 | Weighted median | -0.022 | 0.006 | -0.034 | -0.009 | 0.001 | 0.979 | 0.966 | 0.991 | 0.016 |
|  |  | 12 | MR-Egger | -0.037 | 0.023 | -0.082 | 0.008 | 0.109 | 0.964 | 0.922 | 1.008 | 0.475 |
|  |  | 9 | MR-PRESSO | -0.020 | 0.007 | -0.033 | -0.006 | 0.021 | 0.980 | 0.967 | 0.994 | 0.176 |
|  |  | 12 | Contamination mixture | -0.019 | 0.008 | -0.029 | 0.001 | 0.069 | 0.982 | 0.972 | 1.001 | 0.365 |
|  | MI | 12 | IVW | -0.019 | 0.013 | -0.043 | 0.006 | 0.134 | 0.981 | 0.957 | 1.006 | 0.521 |
|  |  | 12 | Weighted median | -0.013 | 0.009 | -0.031 | 0.004 | 0.140 | 0.987 | 0.970 | 1.004 | 0.532 |
|  |  | 12 | MR-Egger | -0.045 | 0.035 | -0.115 | 0.024 | 0.201 | 0.956 | 0.892 | 1.024 | 0.615 |
|  |  | 8 | MR-PRESSO | -0.012 | 0.006 | -0.024 | 0.000 | 0.088 | 0.988 | 0.977 | 1.000 | 0.422 |
|  |  | 12 | Contamination mixture | -0.007 | 0.005 | -0.017 | 0.003 | 0.192 | 0.993 | 0.983 | 1.003 | 0.605 |
|  | HF | 12 | IVW | 0.004 | 0.007 | -0.009 | 0.017 | 0.517 | 1.004 | 0.991 | 1.017 | 0.872 |
|  |  | 12 | Weighted median | -0.003 | 0.009 | -0.020 | 0.014 | 0.701 | 0.997 | 0.980 | 1.014 | 0.942 |
|  |  | 12 | MR-Egger | -0.005 | 0.019 | -0.042 | 0.032 | 0.794 | 0.995 | 0.959 | 1.032 | 0.971 |
|  |  | 12 | MR-PRESSO | 0.004 | 0.007 | -0.009 | 0.017 | 0.531 | 1.004 | 0.991 | 1.017 | 0.883 |
|  |  | 12 | Contamination mixture | 0.001 | 0.008 | -0.019 | 0.011 | 0.828 | 1.001 | 0.981 | 1.011 | 0.971 |
|  | AF | 12 | IVW | 0.002 | 0.006 | -0.009 | 0.013 | 0.713 | 1.002 | 0.991 | 1.013 | 0.943 |
|  |  | 12 | Weighted median | 0.005 | 0.007 | -0.009 | 0.020 | 0.456 | 1.005 | 0.991 | 1.020 | 0.835 |
|  |  | 12 | MR-Egger | 0.006 | 0.017 | -0.027 | 0.038 | 0.734 | 1.006 | 0.973 | 1.039 | 0.951 |
|  |  | 12 | MR-PRESSO | 0.002 | 0.006 | -0.009 | 0.013 | 0.720 | 1.002 | 0.991 | 1.013 | 0.945 |
|  |  | 12 | Contamination mixture | 0.002 | 0.005 | -0.008 | 0.012 | 0.581 | 1.002 | 0.992 | 1.012 | 0.896 |
|  | Stroke | 12 | IVW | -0.013 | 0.006 | -0.025 | -0.001 | 0.031 | 0.987 | 0.975 | 0.999 | 0.231 |
|  |  | 12 | Weighted median | -0.017 | 0.008 | -0.033 | -0.001 | 0.033 | 0.983 | 0.967 | 0.999 | 0.234 |
|  |  | 12 | MR-Egger | -0.021 | 0.017 | -0.055 | 0.013 | 0.219 | 0.979 | 0.946 | 1.013 | 0.628 |
|  |  | 12 | MR-PRESSO | -0.013 | 0.006 | -0.025 | -0.001 | 0.054 | 0.987 | 0.975 | 0.999 | 0.321 |
|  |  | 12 | Contamination mixture | -0.030 | 0.013 | -0.040 | 0.010 | 0.205 | 0.971 | 0.961 | 1.010 | 0.615 |
| IL1R2 | COPD | 14 | IVW | -0.018 | 0.009 | -0.035 | -0.002 | 0.032 | 0.982 | 0.965 | 0.998 | 0.233 |
|  |  | 14 | Weighted median | -0.014 | 0.005 | -0.023 | -0.005 | 0.002 | 0.986 | 0.977 | 0.995 | 0.040 |
|  |  | 14 | MR-Egger | 0.003 | 0.017 | -0.031 | 0.036 | 0.871 | 1.003 | 0.970 | 1.037 | 0.971 |
|  |  | 11 | MR-PRESSO | -0.020 | 0.006 | -0.031 | -0.009 | 0.005 | 0.980 | 0.970 | 0.991 | 0.067 |
|  |  | 14 | Contamination mixture | -0.020 | 0.010 | -0.050 | -0.010 | 0.000 | 0.980 | 0.951 | 0.990 | 0.011 |
|  | Asthma | 14 | IVW | -0.114 | 0.044 | -0.201 | -0.027 | 0.010 | 0.892 | 0.818 | 0.973 | 0.110 |
|  |  | 14 | Weighted median | -0.061 | 0.014 | -0.089 | -0.034 | 0.000 | 0.941 | 0.915 | 0.966 | 0.001 |
|  |  | 14 | MR-Egger | -0.017 | 0.089 | -0.190 | 0.157 | 0.850 | 0.983 | 0.827 | 1.170 | 0.971 |
|  |  | 7 | MR-PRESSO | -0.055 | 0.025 | -0.104 | -0.006 | 0.070 | 0.946 | 0.901 | 0.994 | 0.370 |
|  |  | 14 | Contamination mixture | -0.047 | 0.010 | -0.077 | -0.037 | 0.004 | 0.954 | 0.926 | 0.964 | 0.056 |
|  | CAD | 12 | IVW | -0.020 | 0.010 | -0.039 | -0.001 | 0.043 | 0.980 | 0.962 | 0.999 | 0.284 |
|  |  | 12 | Weighted median | -0.018 | 0.011 | -0.039 | 0.004 | 0.104 | 0.982 | 0.962 | 1.004 | 0.462 |
|  |  | 12 | MR-Egger | -0.020 | 0.021 | -0.060 | 0.021 | 0.343 | 0.980 | 0.941 | 1.021 | 0.752 |
|  |  | 12 | MR-PRESSO | -0.020 | 0.010 | -0.039 | -0.001 | 0.067 | 0.980 | 0.962 | 0.999 | 0.359 |
|  |  | 12 | Contamination mixture | -0.029 | 0.008 | -0.039 | -0.009 | 0.024 | 0.971 | 0.962 | 0.991 | 0.196 |
|  | MI | 11 | IVW | -0.023 | 0.018 | -0.058 | 0.011 | 0.184 | 0.977 | 0.944 | 1.011 | 0.603 |
|  |  | 11 | Weighted median | -0.017 | 0.017 | -0.051 | 0.016 | 0.315 | 0.983 | 0.951 | 1.016 | 0.731 |
|  |  | 11 | MR-Egger | 0.040 | 0.030 | -0.020 | 0.099 | 0.190 | 1.041 | 0.980 | 1.104 | 0.605 |
|  |  | 11 | MR-PRESSO | -0.023 | 0.018 | -0.058 | 0.011 | 0.213 | 0.977 | 0.944 | 1.011 | 0.623 |
|  |  | 11 | Contamination mixture | -0.085 | 0.020 | -0.125 | -0.045 | 0.024 | 0.918 | 0.882 | 0.956 | 0.198 |
|  | HF | 11 | IVW | 0.044 | 0.014 | 0.017 | 0.071 | 0.002 | 1.045 | 1.017 | 1.074 | 0.029 |
|  |  | 11 | Weighted median | 0.034 | 0.017 | 0.001 | 0.067 | 0.046 | 1.034 | 1.001 | 1.069 | 0.293 |
|  |  | 11 | MR-Egger | 0.037 | 0.029 | -0.020 | 0.094 | 0.202 | 1.038 | 0.980 | 1.099 | 0.615 |
|  |  | 11 | MR-PRESSO | 0.044 | 0.011 | 0.023 | 0.065 | 0.002 | 1.045 | 1.023 | 1.067 | 0.040 |
|  |  | 11 | Contamination mixture | 0.047 | 0.028 | 0.017 | 0.127 | 0.011 | 1.049 | 1.018 | 1.136 | 0.118 |
|  | AF | 12 | IVW | -0.010 | 0.012 | -0.033 | 0.013 | 0.401 | 0.990 | 0.967 | 1.013 | 0.800 |
|  |  | 12 | Weighted median | -0.007 | 0.014 | -0.036 | 0.021 | 0.619 | 0.993 | 0.965 | 1.021 | 0.912 |
|  |  | 12 | MR-Egger | 0.010 | 0.025 | -0.039 | 0.059 | 0.686 | 1.010 | 0.962 | 1.061 | 0.933 |
|  |  | 12 | MR-PRESSO | -0.010 | 0.010 | -0.030 | 0.010 | 0.341 | 0.990 | 0.971 | 1.010 | 0.752 |
|  |  | 12 | Contamination mixture | -0.004 | 0.010 | -0.024 | 0.016 | 0.695 | 0.996 | 0.977 | 1.017 | 0.938 |
|  | Stroke | 11 | IVW | 0.016 | 0.014 | -0.011 | 0.043 | 0.246 | 1.016 | 0.989 | 1.044 | 0.659 |
|  |  | 11 | Weighted median | 0.014 | 0.015 | -0.015 | 0.043 | 0.351 | 1.014 | 0.985 | 1.044 | 0.756 |
|  |  | 11 | MR-Egger | -0.010 | 0.029 | -0.067 | 0.047 | 0.734 | 0.990 | 0.935 | 1.048 | 0.951 |
|  |  | 11 | MR-PRESSO | 0.016 | 0.014 | -0.011 | 0.043 | 0.273 | 1.016 | 0.989 | 1.044 | 0.695 |
|  |  | 11 | Contamination mixture | 0.009 | 0.013 | -0.021 | 0.029 | 0.568 | 1.010 | 0.980 | 1.030 | 0.896 |
| IL6ST | COPD | 13 | IVW | 0.014 | 0.006 | 0.003 | 0.025 | 0.013 | 1.014 | 1.003 | 1.025 | 0.135 |
|  |  | 13 | Weighted median | 0.013 | 0.007 | 0.000 | 0.026 | 0.059 | 1.013 | 1.000 | 1.026 | 0.334 |
|  |  | 13 | MR-Egger | 0.015 | 0.011 | -0.007 | 0.037 | 0.181 | 1.015 | 0.993 | 1.038 | 0.599 |
|  |  | 13 | MR-PRESSO | 0.014 | 0.006 | 0.003 | 0.025 | 0.029 | 1.014 | 1.003 | 1.025 | 0.221 |
|  |  | 13 | Contamination mixture | 0.010 | 0.003 | 0.010 | 0.020 | 0.044 | 1.010 | 1.010 | 1.020 | 0.286 |
|  | Asthma | 13 | IVW | 0.054 | 0.015 | 0.025 | 0.082 | 0.000 | 1.055 | 1.025 | 1.086 | 0.007 |
|  |  | 13 | Weighted median | 0.046 | 0.019 | 0.008 | 0.084 | 0.019 | 1.047 | 1.008 | 1.087 | 0.171 |
|  |  | 13 | MR-Egger | 0.033 | 0.028 | -0.021 | 0.088 | 0.232 | 1.034 | 0.979 | 1.092 | 0.645 |
|  |  | 13 | MR-PRESSO | 0.054 | 0.014 | 0.026 | 0.081 | 0.002 | 1.055 | 1.027 | 1.085 | 0.040 |
|  |  | 13 | Contamination mixture | 0.060 | 0.026 | 0.030 | 0.130 | 0.010 | 1.062 | 1.030 | 1.139 | 0.112 |
|  | CAD | 13 | IVW | 0.023 | 0.017 | -0.010 | 0.056 | 0.165 | 1.023 | 0.990 | 1.057 | 0.576 |
|  |  | 13 | Weighted median | -0.008 | 0.017 | -0.041 | 0.025 | 0.652 | 0.992 | 0.960 | 1.026 | 0.926 |
|  |  | 13 | MR-Egger | -0.027 | 0.027 | -0.081 | 0.026 | 0.316 | 0.973 | 0.923 | 1.026 | 0.731 |
|  |  | 13 | MR-PRESSO | 0.023 | 0.017 | -0.010 | 0.056 | 0.191 | 1.023 | 0.990 | 1.057 | 0.605 |
|  |  | 13 | Contamination mixture | 0.078 | 0.043 | -0.012 | 0.158 | 0.130 | 1.081 | 0.988 | 1.171 | 0.518 |
|  | MI | 12 | IVW | 0.049 | 0.019 | 0.011 | 0.087 | 0.011 | 1.050 | 1.011 | 1.091 | 0.121 |
|  |  | 12 | Weighted median | 0.019 | 0.025 | -0.030 | 0.068 | 0.451 | 1.019 | 0.970 | 1.071 | 0.832 |
|  |  | 12 | MR-Egger | -0.018 | 0.036 | -0.088 | 0.053 | 0.622 | 0.982 | 0.916 | 1.054 | 0.912 |
|  |  | 12 | MR-PRESSO | 0.049 | 0.019 | 0.012 | 0.086 | 0.026 | 1.050 | 1.012 | 1.090 | 0.207 |
|  |  | 12 | Contamination mixture | 0.104 | 0.048 | 0.014 | 0.204 | 0.031 | 1.110 | 1.014 | 1.227 | 0.231 |
|  | HF | 12 | IVW | 0.015 | 0.020 | -0.024 | 0.054 | 0.458 | 1.015 | 0.976 | 1.055 | 0.836 |
|  |  | 12 | Weighted median | 0.011 | 0.025 | -0.038 | 0.061 | 0.656 | 1.011 | 0.962 | 1.063 | 0.927 |
|  |  | 12 | MR-Egger | -0.006 | 0.037 | -0.078 | 0.066 | 0.871 | 0.994 | 0.925 | 1.069 | 0.971 |
|  |  | 12 | MR-PRESSO | 0.015 | 0.019 | -0.022 | 0.051 | 0.448 | 1.015 | 0.978 | 1.053 | 0.828 |
|  |  | 12 | Contamination mixture | 0.031 | 0.033 | -0.029 | 0.101 | 0.301 | 1.031 | 0.971 | 1.106 | 0.724 |
|  | AF | 13 | IVW | 0.052 | 0.017 | 0.019 | 0.085 | 0.002 | 1.053 | 1.019 | 1.089 | 0.035 |
|  |  | 13 | Weighted median | 0.046 | 0.021 | 0.004 | 0.088 | 0.032 | 1.047 | 1.004 | 1.092 | 0.233 |
|  |  | 13 | MR-Egger | 0.035 | 0.031 | -0.026 | 0.096 | 0.265 | 1.035 | 0.974 | 1.100 | 0.685 |
|  |  | 13 | MR-PRESSO | 0.052 | 0.013 | 0.027 | 0.077 | 0.001 | 1.053 | 1.028 | 1.080 | 0.026 |
|  |  | 13 | Contamination mixture | 0.044 | 0.018 | 0.014 | 0.084 | 0.009 | 1.045 | 1.014 | 1.087 | 0.101 |
|  | Stroke | 11 | IVW | -0.010 | 0.018 | -0.045 | 0.025 | 0.574 | 0.990 | 0.956 | 1.025 | 0.896 |
|  |  | 11 | Weighted median | -0.005 | 0.022 | -0.047 | 0.038 | 0.835 | 0.996 | 0.954 | 1.039 | 0.971 |
|  |  | 11 | MR-Egger | -0.006 | 0.033 | -0.070 | 0.059 | 0.858 | 0.994 | 0.932 | 1.060 | 0.971 |
|  |  | 11 | MR-PRESSO | -0.010 | 0.010 | -0.030 | 0.010 | 0.343 | 0.990 | 0.971 | 1.010 | 0.752 |
|  |  | 11 | Contamination mixture | -0.003 | 0.020 | -0.053 | 0.027 | 0.769 | 0.997 | 0.948 | 1.027 | 0.965 |
| IL18 | COPD | 6 | IVW | 0.034 | 0.008 | 0.019 | 0.049 | 0.000 | 1.035 | 1.019 | 1.051 | 0.001 |
|  |  | 6 | Weighted median | 0.031 | 0.009 | 0.013 | 0.049 | 0.001 | 1.031 | 1.013 | 1.050 | 0.019 |
|  |  | 6 | MR-Egger | 0.018 | 0.021 | -0.022 | 0.059 | 0.371 | 1.019 | 0.978 | 1.061 | 0.772 |
|  |  | 6 | MR-PRESSO | 0.034 | 0.008 | 0.019 | 0.049 | 0.007 | 1.035 | 1.019 | 1.051 | 0.084 |
|  | Asthma | 6 | IVW | 0.010 | 0.024 | -0.036 | 0.057 | 0.660 | 1.010 | 0.965 | 1.058 | 0.928 |
|  |  | 6 | Weighted median | 0.027 | 0.026 | -0.025 | 0.078 | 0.306 | 1.027 | 0.976 | 1.081 | 0.727 |
|  |  | 6 | MR-Egger | 0.078 | 0.057 | -0.034 | 0.191 | 0.174 | 1.081 | 0.966 | 1.210 | 0.592 |
|  |  | 6 | MR-PRESSO | 0.010 | 0.024 | -0.036 | 0.057 | 0.678 | 1.010 | 0.965 | 1.058 | 0.932 |
|  | CAD | 6 | IVW | -0.004 | 0.019 | -0.041 | 0.033 | 0.833 | 0.996 | 0.960 | 1.033 | 0.971 |
|  |  | 6 | Weighted median | 0.004 | 0.022 | -0.039 | 0.048 | 0.849 | 1.004 | 0.961 | 1.049 | 0.971 |
|  |  | 6 | MR-Egger | 0.018 | 0.049 | -0.079 | 0.115 | 0.720 | 1.018 | 0.924 | 1.121 | 0.945 |
|  |  | 6 | MR-PRESSO | -0.004 | 0.015 | -0.032 | 0.025 | 0.797 | 0.996 | 0.968 | 1.025 | 0.971 |
|  | MI | 6 | IVW | 0.011 | 0.029 | -0.046 | 0.067 | 0.711 | 1.011 | 0.955 | 1.069 | 0.943 |
|  |  | 6 | Weighted median | 0.015 | 0.034 | -0.052 | 0.081 | 0.661 | 1.015 | 0.950 | 1.085 | 0.928 |
|  |  | 6 | MR-Egger | 0.000 | 0.075 | -0.147 | 0.146 | 0.998 | 1.000 | 0.864 | 1.158 | 1.000 |
|  |  | 6 | MR-PRESSO | 0.011 | 0.014 | -0.017 | 0.038 | 0.477 | 1.011 | 0.984 | 1.039 | 0.847 |
|  | HF | 6 | IVW | 0.039 | 0.029 | -0.019 | 0.097 | 0.183 | 1.040 | 0.982 | 1.102 | 0.603 |
|  |  | 6 | Weighted median | 0.047 | 0.034 | -0.019 | 0.114 | 0.165 | 1.049 | 0.981 | 1.121 | 0.576 |
|  |  | 6 | MR-Egger | 0.075 | 0.080 | -0.081 | 0.231 | 0.344 | 1.078 | 0.923 | 1.260 | 0.752 |
|  |  | 6 | MR-PRESSO | 0.039 | 0.018 | 0.003 | 0.075 | 0.084 | 1.040 | 1.003 | 1.078 | 0.409 |
|  | AF | 6 | IVW | 0.075 | 0.028 | 0.021 | 0.129 | 0.007 | 1.078 | 1.021 | 1.137 | 0.083 |
|  |  | 6 | Weighted median | 0.072 | 0.030 | 0.013 | 0.131 | 0.017 | 1.075 | 1.013 | 1.140 | 0.163 |
|  |  | 6 | MR-Egger | 0.064 | 0.083 | -0.099 | 0.227 | 0.442 | 1.066 | 0.906 | 1.255 | 0.824 |
|  |  | 6 | MR-PRESSO | 0.075 | 0.028 | 0.021 | 0.129 | 0.043 | 1.078 | 1.021 | 1.137 | 0.284 |
|  | Stroke | 5 | IVW | -0.017 | 0.029 | -0.074 | 0.039 | 0.552 | 0.983 | 0.929 | 1.040 | 0.893 |
|  |  | 5 | Weighted median | -0.017 | 0.033 | -0.081 | 0.047 | 0.599 | 0.983 | 0.922 | 1.048 | 0.902 |
|  |  | 5 | MR-Egger | -0.059 | 0.091 | -0.237 | 0.119 | 0.516 | 0.943 | 0.789 | 1.126 | 0.872 |
|  |  | 5 | MR-PRESSO | -0.017 | 0.029 | -0.074 | 0.039 | 0.584 | 0.983 | 0.929 | 1.040 | 0.899 |
| CX3CL1 | COPD | 8 | IVW | -0.012 | 0.006 | -0.025 | 0.000 | 0.058 | 0.988 | 0.976 | 1.000 | 0.333 |
|  |  | 8 | Weighted median | -0.008 | 0.008 | -0.024 | 0.009 | 0.364 | 0.992 | 0.976 | 1.009 | 0.767 |
|  |  | 8 | MR-Egger | -0.005 | 0.022 | -0.047 | 0.038 | 0.834 | 0.995 | 0.954 | 1.039 | 0.971 |
|  |  | 8 | MR-PRESSO | -0.012 | 0.006 | -0.025 | 0.000 | 0.098 | 0.988 | 0.976 | 1.000 | 0.450 |
|  |  | 8 | Contamination mixture | -0.034 | 0.015 | -0.054 | 0.006 | 0.199 | 0.966 | 0.947 | 1.006 | 0.614 |
|  | Asthma | 8 | IVW | -0.059 | 0.023 | -0.104 | -0.014 | 0.011 | 0.943 | 0.901 | 0.987 | 0.118 |
|  |  | 8 | Weighted median | -0.050 | 0.025 | -0.098 | -0.002 | 0.043 | 0.951 | 0.907 | 0.998 | 0.284 |
|  |  | 8 | MR-Egger | -0.001 | 0.075 | -0.149 | 0.147 | 0.988 | 0.999 | 0.862 | 1.158 | 1.000 |
|  |  | 8 | MR-PRESSO | -0.059 | 0.023 | -0.104 | -0.014 | 0.038 | 0.943 | 0.901 | 0.987 | 0.264 |
|  |  | 8 | Contamination mixture | -0.120 | 0.054 | -0.210 | 0.000 | 0.054 | 0.887 | 0.810 | 1.000 | 0.321 |
|  | CAD | 8 | IVW | 0.078 | 0.015 | 0.048 | 0.107 | 0.000 | 1.081 | 1.049 | 1.113 | 0.000 |
|  |  | 8 | Weighted median | 0.062 | 0.019 | 0.024 | 0.099 | 0.001 | 1.064 | 1.024 | 1.104 | 0.025 |
|  |  | 8 | MR-Egger | 0.084 | 0.048 | -0.011 | 0.179 | 0.084 | 1.087 | 0.989 | 1.195 | 0.409 |
|  |  | 8 | MR-PRESSO | 0.078 | 0.012 | 0.055 | 0.100 | 0.000 | 1.081 | 1.057 | 1.105 | 0.008 |
|  |  | 8 | Contamination mixture | 0.080 | 0.015 | 0.050 | 0.110 | 0.000 | 1.083 | 1.051 | 1.116 | 0.005 |
|  | MI | 8 | IVW | 0.113 | 0.023 | 0.068 | 0.158 | 0.000 | 1.120 | 1.071 | 1.172 | 0.000 |
|  |  | 8 | Weighted median | 0.100 | 0.029 | 0.043 | 0.157 | 0.001 | 1.105 | 1.044 | 1.170 | 0.014 |
|  |  | 8 | MR-Egger | 0.086 | 0.074 | -0.059 | 0.230 | 0.244 | 1.090 | 0.943 | 1.259 | 0.658 |
|  |  | 8 | MR-PRESSO | 0.113 | 0.014 | 0.086 | 0.141 | 0.000 | 1.120 | 1.090 | 1.151 | 0.003 |
|  |  | 8 | Contamination mixture | 0.119 | 0.026 | 0.069 | 0.169 | 0.000 | 1.127 | 1.072 | 1.184 | 0.004 |
|  | HF | 8 | IVW | 0.032 | 0.027 | -0.021 | 0.085 | 0.242 | 1.032 | 0.979 | 1.088 | 0.657 |
|  |  | 8 | Weighted median | 0.015 | 0.033 | -0.049 | 0.080 | 0.646 | 1.015 | 0.952 | 1.083 | 0.924 |
|  |  | 8 | MR-Egger | 0.112 | 0.087 | -0.057 | 0.282 | 0.195 | 1.119 | 0.944 | 1.326 | 0.611 |
|  |  | 8 | MR-PRESSO | 0.032 | 0.027 | -0.021 | 0.085 | 0.280 | 1.032 | 0.979 | 1.088 | 0.704 |
|  |  | 8 | Contamination mixture | 0.001 | 0.071 | -0.079 | 0.201 | 0.931 | 1.001 | 0.924 | 1.223 | 0.993 |
|  | AF | 8 | IVW | 0.020 | 0.023 | -0.025 | 0.065 | 0.386 | 1.020 | 0.975 | 1.067 | 0.786 |
|  |  | 8 | Weighted median | 0.021 | 0.027 | -0.033 | 0.075 | 0.442 | 1.021 | 0.968 | 1.078 | 0.824 |
|  |  | 8 | MR-Egger | 0.105 | 0.071 | -0.033 | 0.243 | 0.137 | 1.111 | 0.967 | 1.276 | 0.528 |
|  |  | 8 | MR-PRESSO | 0.020 | 0.023 | -0.025 | 0.065 | 0.415 | 1.020 | 0.975 | 1.067 | 0.808 |
|  |  | 8 | Contamination mixture | 0.010 | 0.069 | -0.130 | 0.140 | 0.767 | 1.010 | 0.878 | 1.150 | 0.965 |
|  | Stroke | 8 | IVW | 0.054 | 0.022 | 0.010 | 0.097 | 0.015 | 1.055 | 1.010 | 1.102 | 0.152 |
|  |  | 8 | Weighted median | 0.043 | 0.028 | -0.012 | 0.098 | 0.125 | 1.044 | 0.988 | 1.103 | 0.512 |
|  |  | 8 | MR-Egger | -0.001 | 0.071 | -0.140 | 0.138 | 0.989 | 0.999 | 0.870 | 1.148 | 1.000 |
|  |  | 8 | MR-PRESSO | 0.054 | 0.014 | 0.027 | 0.081 | 0.006 | 1.055 | 1.027 | 1.084 | 0.077 |
|  |  | 8 | Contamination mixture | 0.057 | 0.038 | 0.017 | 0.167 | 0.029 | 1.058 | 1.017 | 1.181 | 0.221 |
| VEGFA | COPD | 18 | IVW | 0.011 | 0.005 | 0.001 | 0.021 | 0.024 | 1.011 | 1.001 | 1.021 | 0.198 |
|  |  | 18 | Weighted median | 0.008 | 0.005 | -0.002 | 0.018 | 0.115 | 1.008 | 0.998 | 1.019 | 0.485 |
|  |  | 18 | MR-Egger | 0.005 | 0.009 | -0.011 | 0.022 | 0.527 | 1.005 | 0.989 | 1.022 | 0.880 |
|  |  | 18 | MR-PRESSO | 0.011 | 0.005 | 0.001 | 0.021 | 0.038 | 1.011 | 1.001 | 1.021 | 0.261 |
|  |  | 18 | Contamination mixture | 0.014 | 0.003 | 0.004 | 0.014 | 0.043 | 1.014 | 1.004 | 1.014 | 0.284 |
|  | Asthma | 18 | IVW | -0.009 | 0.013 | -0.034 | 0.017 | 0.504 | 0.991 | 0.967 | 1.017 | 0.869 |
|  |  | 18 | Weighted median | 0.004 | 0.015 | -0.026 | 0.033 | 0.812 | 1.004 | 0.974 | 1.034 | 0.971 |
|  |  | 18 | MR-Egger | 0.008 | 0.023 | -0.036 | 0.052 | 0.719 | 1.008 | 0.964 | 1.054 | 0.945 |
|  |  | 18 | MR-PRESSO | -0.009 | 0.013 | -0.034 | 0.017 | 0.513 | 0.991 | 0.967 | 1.017 | 0.871 |
|  |  | 18 | Contamination mixture | -0.005 | 0.023 | -0.075 | 0.015 | 1.000 | 0.995 | 0.928 | 1.015 | 1.000 |
|  | CAD | 18 | IVW | -0.003 | 0.011 | -0.025 | 0.020 | 0.821 | 0.997 | 0.975 | 1.020 | 0.971 |
|  |  | 18 | Weighted median | -0.007 | 0.012 | -0.031 | 0.018 | 0.594 | 0.993 | 0.969 | 1.018 | 0.900 |
|  |  | 18 | MR-Egger | 0.004 | 0.020 | -0.035 | 0.044 | 0.827 | 1.004 | 0.966 | 1.045 | 0.971 |
|  |  | 18 | MR-PRESSO | -0.003 | 0.011 | -0.025 | 0.020 | 0.824 | 0.997 | 0.975 | 1.020 | 0.971 |
|  |  | 18 | Contamination mixture | -0.012 | 0.008 | -0.022 | 0.008 | 0.495 | 0.988 | 0.978 | 1.008 | 0.860 |
|  | MI | 17 | IVW | -0.011 | 0.015 | -0.041 | 0.018 | 0.444 | 0.989 | 0.960 | 1.018 | 0.824 |
|  |  | 17 | Weighted median | -0.022 | 0.019 | -0.059 | 0.015 | 0.236 | 0.978 | 0.943 | 1.015 | 0.650 |
|  |  | 17 | MR-Egger | -0.031 | 0.025 | -0.080 | 0.019 | 0.227 | 0.970 | 0.923 | 1.019 | 0.638 |
|  |  | 17 | MR-PRESSO | -0.011 | 0.013 | -0.037 | 0.014 | 0.394 | 0.989 | 0.964 | 1.014 | 0.796 |
|  |  | 17 | Contamination mixture | -0.030 | 0.015 | -0.060 | 0.000 | 0.152 | 0.971 | 0.942 | 1.000 | 0.550 |
|  | HF | 18 | IVW | 0.029 | 0.017 | -0.004 | 0.063 | 0.087 | 1.030 | 0.996 | 1.065 | 0.418 |
|  |  | 18 | Weighted median | 0.009 | 0.020 | -0.030 | 0.048 | 0.652 | 1.009 | 0.970 | 1.050 | 0.926 |
|  |  | 18 | MR-Egger | -0.029 | 0.026 | -0.080 | 0.023 | 0.275 | 0.972 | 0.923 | 1.023 | 0.697 |
|  |  | 18 | MR-PRESSO | 0.029 | 0.017 | -0.004 | 0.063 | 0.105 | 1.030 | 0.996 | 1.065 | 0.467 |
|  |  | 18 | Contamination mixture | 0.138 | 0.056 | -0.012 | 0.208 | 0.108 | 1.148 | 0.988 | 1.231 | 0.475 |
|  | AF | 18 | IVW | 0.009 | 0.015 | -0.021 | 0.039 | 0.556 | 1.009 | 0.979 | 1.040 | 0.896 |
|  |  | 18 | Weighted median | 0.004 | 0.017 | -0.029 | 0.038 | 0.794 | 1.004 | 0.971 | 1.039 | 0.971 |
|  |  | 18 | MR-Egger | 0.021 | 0.027 | -0.033 | 0.074 | 0.451 | 1.021 | 0.968 | 1.077 | 0.832 |
|  |  | 18 | MR-PRESSO | 0.009 | 0.015 | -0.021 | 0.039 | 0.564 | 1.009 | 0.979 | 1.040 | 0.896 |
|  |  | 18 | Contamination mixture | 0.010 | 0.023 | -0.020 | 0.070 | 0.584 | 1.010 | 0.980 | 1.072 | 0.899 |
|  | Stroke | 18 | IVW | 0.016 | 0.016 | -0.015 | 0.048 | 0.315 | 1.016 | 0.985 | 1.049 | 0.731 |
|  |  | 18 | Weighted median | -0.014 | 0.022 | -0.058 | 0.030 | 0.544 | 0.986 | 0.944 | 1.031 | 0.891 |
|  |  | 18 | MR-Egger | 0.002 | 0.029 | -0.055 | 0.060 | 0.937 | 1.002 | 0.946 | 1.061 | 0.995 |
|  |  | 18 | MR-PRESSO | 0.016 | 0.016 | -0.015 | 0.048 | 0.329 | 1.016 | 0.985 | 1.049 | 0.737 |
|  |  | 18 | Contamination mixture | 0.001 | 0.046 | -0.049 | 0.131 | 0.922 | 1.001 | 0.952 | 1.140 | 0.988 |
| CRP | COPD | 5 | IVW | 0.024 | 0.008 | 0.008 | 0.041 | 0.004 | 1.025 | 1.008 | 1.042 | 0.055 |
|  |  | 5 | Weighted median | 0.022 | 0.010 | 0.003 | 0.042 | 0.026 | 1.022 | 1.003 | 1.043 | 0.206 |
|  |  | 5 | MR-Egger | -0.007 | 0.026 | -0.058 | 0.044 | 0.782 | 0.993 | 0.944 | 1.044 | 0.971 |
|  |  | 5 | MR-PRESSO | 0.024 | 0.007 | 0.011 | 0.037 | 0.021 | 1.025 | 1.011 | 1.038 | 0.179 |
|  | Asthma | 5 | IVW | 0.001 | 0.024 | -0.046 | 0.048 | 0.974 | 1.001 | 0.955 | 1.049 | 1.000 |
|  |  | 5 | Weighted median | -0.011 | 0.028 | -0.066 | 0.044 | 0.690 | 0.989 | 0.936 | 1.045 | 0.935 |
|  |  | 5 | MR-Egger | -0.118 | 0.074 | -0.263 | 0.026 | 0.109 | 0.888 | 0.769 | 1.027 | 0.475 |
|  |  | 5 | MR-PRESSO | 0.001 | 0.022 | -0.043 | 0.044 | 0.973 | 1.001 | 0.958 | 1.045 | 1.000 |
|  | CAD | 5 | IVW | -0.012 | 0.023 | -0.058 | 0.033 | 0.597 | 0.988 | 0.944 | 1.034 | 0.901 |
|  |  | 5 | Weighted median | -0.019 | 0.024 | -0.066 | 0.029 | 0.440 | 0.981 | 0.936 | 1.029 | 0.824 |
|  |  | 5 | MR-Egger | -0.148 | 0.064 | -0.274 | -0.023 | 0.020 | 0.862 | 0.760 | 0.977 | 0.175 |
|  |  | 5 | MR-PRESSO | -0.012 | 0.023 | -0.058 | 0.033 | 0.625 | 0.988 | 0.944 | 1.034 | 0.912 |
|  | MI | 5 | IVW | 0.009 | 0.031 | -0.051 | 0.069 | 0.769 | 1.009 | 0.950 | 1.071 | 0.965 |
|  |  | 5 | Weighted median | -0.005 | 0.034 | -0.072 | 0.062 | 0.883 | 0.995 | 0.930 | 1.064 | 0.974 |
|  |  | 5 | MR-Egger | -0.129 | 0.095 | -0.315 | 0.058 | 0.177 | 0.879 | 0.730 | 1.060 | 0.594 |
|  |  | 5 | MR-PRESSO | 0.009 | 0.031 | -0.051 | 0.069 | 0.783 | 1.009 | 0.950 | 1.071 | 0.971 |
|  | HF | 5 | IVW | 0.005 | 0.032 | -0.057 | 0.068 | 0.868 | 1.005 | 0.944 | 1.070 | 0.971 |
|  |  | 5 | Weighted median | 0.004 | 0.037 | -0.069 | 0.078 | 0.906 | 1.004 | 0.933 | 1.081 | 0.981 |
|  |  | 5 | MR-Egger | 0.125 | 0.111 | -0.092 | 0.343 | 0.258 | 1.134 | 0.912 | 1.409 | 0.675 |
|  |  | 5 | MR-PRESSO | 0.005 | 0.022 | -0.037 | 0.048 | 0.818 | 1.005 | 0.964 | 1.049 | 0.971 |
|  | AF | 5 | IVW | 0.027 | 0.035 | -0.042 | 0.095 | 0.445 | 1.027 | 0.959 | 1.100 | 0.824 |
|  |  | 5 | Weighted median | 0.049 | 0.031 | -0.012 | 0.111 | 0.114 | 1.051 | 0.988 | 1.117 | 0.485 |
|  |  | 5 | MR-Egger | 0.125 | 0.124 | -0.118 | 0.369 | 0.314 | 1.133 | 0.888 | 1.446 | 0.731 |
|  |  | 5 | MR-PRESSO | 0.027 | 0.035 | -0.042 | 0.095 | 0.487 | 1.027 | 0.959 | 1.100 | 0.855 |
|  | Stroke | 4 | IVW | -0.013 | 0.037 | -0.086 | 0.060 | 0.727 | 0.987 | 0.918 | 1.062 | 0.948 |
|  |  | 4 | Weighted median | 0.012 | 0.036 | -0.058 | 0.082 | 0.738 | 1.012 | 0.943 | 1.086 | 0.953 |
|  |  | 4 | MR-Egger | -0.127 | 0.148 | -0.417 | 0.164 | 0.393 | 0.881 | 0.659 | 1.178 | 0.794 |
| CCL17 | COPD | 3 | IVW | -0.008 | 0.007 | -0.021 | 0.005 | 0.217 | 0.992 | 0.979 | 1.005 | 0.627 |
|  |  | 3 | Weighted median | -0.008 | 0.007 | -0.022 | 0.006 | 0.268 | 0.992 | 0.978 | 1.006 | 0.689 |
|  |  | 3 | MR-Egger | -0.017 | 0.023 | -0.063 | 0.028 | 0.455 | 0.983 | 0.939 | 1.029 | 0.835 |
|  | Asthma | 3 | IVW | -0.054 | 0.019 | -0.091 | -0.017 | 0.004 | 0.947 | 0.913 | 0.983 | 0.062 |
|  |  | 3 | Weighted median | -0.050 | 0.021 | -0.091 | -0.008 | 0.020 | 0.951 | 0.913 | 0.992 | 0.173 |
|  |  | 3 | MR-Egger | -0.003 | 0.065 | -0.131 | 0.125 | 0.958 | 0.997 | 0.877 | 1.133 | 0.999 |
|  | CAD | 3 | IVW | 0.042 | 0.015 | 0.012 | 0.072 | 0.007 | 1.043 | 1.012 | 1.075 | 0.081 |
|  |  | 3 | Weighted median | 0.043 | 0.018 | 0.008 | 0.078 | 0.016 | 1.044 | 1.008 | 1.081 | 0.155 |
|  |  | 3 | MR-Egger | 0.034 | 0.064 | -0.091 | 0.159 | 0.598 | 1.034 | 0.913 | 1.172 | 0.902 |
|  | MI | 3 | IVW | 0.056 | 0.023 | 0.010 | 0.102 | 0.017 | 1.057 | 1.010 | 1.107 | 0.163 |
|  |  | 3 | Weighted median | 0.057 | 0.028 | 0.002 | 0.112 | 0.042 | 1.059 | 1.002 | 1.119 | 0.279 |
|  |  | 3 | MR-Egger | 0.028 | 0.141 | -0.248 | 0.304 | 0.843 | 1.028 | 0.781 | 1.355 | 0.971 |
|  | HF | 3 | IVW | 0.016 | 0.025 | -0.033 | 0.065 | 0.524 | 1.016 | 0.967 | 1.068 | 0.879 |
|  |  | 3 | Weighted median | 0.012 | 0.028 | -0.042 | 0.066 | 0.661 | 1.012 | 0.959 | 1.068 | 0.928 |
|  |  | 3 | MR-Egger | -0.022 | 0.087 | -0.193 | 0.150 | 0.805 | 0.979 | 0.825 | 1.162 | 0.971 |
|  | AF | 3 | IVW | 0.023 | 0.021 | -0.018 | 0.064 | 0.281 | 1.023 | 0.982 | 1.066 | 0.704 |
|  |  | 3 | Weighted median | 0.022 | 0.023 | -0.024 | 0.068 | 0.344 | 1.022 | 0.977 | 1.070 | 0.753 |
|  |  | 3 | MR-Egger | -0.036 | 0.072 | -0.178 | 0.106 | 0.623 | 0.965 | 0.837 | 1.112 | 0.912 |
|  | Stroke | 3 | IVW | 0.030 | 0.023 | -0.015 | 0.076 | 0.191 | 1.031 | 0.985 | 1.079 | 0.605 |
|  |  | 3 | Weighted median | 0.045 | 0.026 | -0.006 | 0.097 | 0.083 | 1.047 | 0.994 | 1.102 | 0.409 |
|  |  | 3 | MR-Egger | 0.096 | 0.095 | -0.090 | 0.282 | 0.310 | 1.101 | 0.914 | 1.326 | 0.727 |
| CCL8 | COPD | 5 | IVW | -0.005 | 0.002 | -0.009 | -0.001 | 0.016 | 0.995 | 0.991 | 0.999 | 0.157 |
|  |  | 5 | Weighted median | -0.003 | 0.002 | -0.007 | 0.001 | 0.140 | 0.997 | 0.993 | 1.001 | 0.531 |
|  |  | 5 | MR-Egger | -0.003 | 0.004 | -0.010 | 0.005 | 0.536 | 0.997 | 0.990 | 1.005 | 0.888 |
|  |  | 5 | MR-PRESSO | -0.005 | 0.002 | -0.009 | -0.001 | 0.074 | 0.995 | 0.991 | 0.999 | 0.380 |
|  | Asthma | 5 | IVW | 0.001 | 0.005 | -0.009 | 0.012 | 0.826 | 1.001 | 0.991 | 1.012 | 0.971 |
|  |  | 5 | Weighted median | 0.002 | 0.006 | -0.010 | 0.014 | 0.723 | 1.002 | 0.990 | 1.014 | 0.946 |
|  |  | 5 | MR-Egger | -0.001 | 0.010 | -0.021 | 0.019 | 0.921 | 0.999 | 0.979 | 1.019 | 0.988 |
|  |  | 5 | MR-PRESSO | 0.001 | 0.002 | -0.004 | 0.006 | 0.649 | 1.001 | 0.996 | 1.006 | 0.924 |
|  | CAD | 5 | IVW | 0.002 | 0.008 | -0.014 | 0.019 | 0.782 | 1.002 | 0.986 | 1.019 | 0.971 |
|  |  | 5 | Weighted median | 0.003 | 0.005 | -0.008 | 0.013 | 0.619 | 1.003 | 0.992 | 1.013 | 0.912 |
|  |  | 5 | MR-Egger | 0.016 | 0.016 | -0.015 | 0.048 | 0.309 | 1.016 | 0.985 | 1.049 | 0.727 |
|  |  | 5 | MR-PRESSO | 0.002 | 0.008 | -0.014 | 0.019 | 0.796 | 1.002 | 0.986 | 1.019 | 0.971 |
|  | MI | 5 | IVW | -0.002 | 0.006 | -0.014 | 0.011 | 0.802 | 0.998 | 0.986 | 1.011 | 0.971 |
|  | MI | 5 | Weighted median | -0.001 | 0.007 | -0.015 | 0.013 | 0.905 | 0.999 | 0.985 | 1.013 | 0.981 |
|  | MI | 5 | MR-Egger | 0.012 | 0.013 | -0.012 | 0.037 | 0.327 | 1.012 | 0.988 | 1.037 | 0.734 |
|  | MI | 5 | MR-PRESSO | -0.002 | 0.006 | -0.013 | 0.009 | 0.787 | 0.998 | 0.988 | 1.009 | 0.971 |
|  | HF | 5 | IVW | 0.011 | 0.007 | -0.003 | 0.025 | 0.122 | 1.011 | 0.997 | 1.025 | 0.504 |
|  | HF | 5 | Weighted median | 0.013 | 0.008 | -0.003 | 0.028 | 0.108 | 1.013 | 0.997 | 1.029 | 0.475 |
|  | HF | 5 | MR-Egger | 0.026 | 0.014 | 0.000 | 0.053 | 0.054 | 1.026 | 1.000 | 1.054 | 0.321 |
|  | HF | 5 | MR-PRESSO | 0.011 | 0.007 | -0.003 | 0.024 | 0.188 | 1.011 | 0.997 | 1.025 | 0.605 |
|  | AF | 5 | IVW | 0.009 | 0.006 | -0.003 | 0.021 | 0.139 | 1.009 | 0.997 | 1.021 | 0.529 |
|  | AF | 5 | Weighted median | 0.010 | 0.007 | -0.003 | 0.023 | 0.138 | 1.010 | 0.997 | 1.023 | 0.528 |
|  | AF | 5 | MR-Egger | 0.013 | 0.011 | -0.009 | 0.036 | 0.249 | 1.013 | 0.991 | 1.036 | 0.664 |
|  | AF | 5 | MR-PRESSO | 0.009 | 0.002 | 0.004 | 0.013 | 0.020 | 1.009 | 1.004 | 1.014 | 0.174 |
|  |  | 5 | IVW | 0.009 | 0.007 | -0.004 | 0.022 | 0.190 | 1.009 | 0.996 | 1.022 | 0.605 |
|  |  | 5 | Weighted median | 0.005 | 0.007 | -0.009 | 0.019 | 0.464 | 1.005 | 0.991 | 1.019 | 0.838 |
|  |  | 5 | MR-Egger | 0.004 | 0.015 | -0.024 | 0.033 | 0.776 | 1.004 | 0.976 | 1.033 | 0.965 |
|  |  | 5 | MR-PRESSO | 0.009 | 0.007 | -0.004 | 0.022 | 0.260 | 1.009 | 0.996 | 1.022 | 0.677 |
| CCL22 | COPD | 3 | IVW | 0.017 | 0.008 | 0.001 | 0.032 | 0.032 | 1.017 | 1.001 | 1.033 | 0.233 |
|  |  | 3 | Weighted median | 0.004 | 0.011 | -0.017 | 0.025 | 0.683 | 1.004 | 0.984 | 1.026 | 0.932 |
|  |  | 3 | MR-Egger | 0.040 | 0.076 | -0.110 | 0.190 | 0.598 | 1.041 | 0.896 | 1.210 | 0.902 |
|  | Asthma | 3 | IVW | 0.052 | 0.022 | 0.009 | 0.095 | 0.018 | 1.053 | 1.009 | 1.100 | 0.167 |
|  |  | 3 | Weighted median | 0.043 | 0.034 | -0.023 | 0.109 | 0.198 | 1.044 | 0.978 | 1.116 | 0.614 |
|  |  | 3 | MR-Egger | 0.036 | 0.247 | -0.448 | 0.519 | 0.885 | 1.036 | 0.639 | 1.681 | 0.974 |
|  | CAD | 3 | IVW | -0.031 | 0.018 | -0.067 | 0.004 | 0.080 | 0.969 | 0.936 | 1.004 | 0.400 |
|  |  | 3 | Weighted median | -0.062 | 0.025 | -0.111 | -0.013 | 0.012 | 0.940 | 0.895 | 0.987 | 0.133 |
|  |  | 3 | MR-Egger | 0.072 | 0.158 | -0.237 | 0.381 | 0.647 | 1.075 | 0.789 | 1.464 | 0.924 |
|  | MI | 3 | IVW | -0.040 | 0.026 | -0.092 | 0.012 | 0.128 | 0.961 | 0.912 | 1.012 | 0.516 |
|  |  | 3 | Weighted median | -0.093 | 0.037 | -0.167 | -0.020 | 0.012 | 0.911 | 0.847 | 0.980 | 0.132 |
|  |  | 3 | MR-Egger | 0.154 | 0.277 | -0.389 | 0.696 | 0.579 | 1.166 | 0.678 | 2.005 | 0.896 |
|  | HF | 3 | IVW | -0.012 | 0.029 | -0.069 | 0.046 | 0.689 | 0.988 | 0.933 | 1.047 | 0.935 |
|  |  | 3 | Weighted median | -0.002 | 0.041 | -0.082 | 0.079 | 0.969 | 0.998 | 0.922 | 1.082 | 1.000 |
|  |  | 3 | MR-Egger | -0.001 | 0.244 | -0.480 | 0.478 | 0.997 | 0.999 | 0.619 | 1.613 | 1.000 |
|  | AF | 3 | IVW | -0.057 | 0.025 | -0.106 | -0.009 | 0.019 | 0.944 | 0.900 | 0.991 | 0.171 |
|  |  | 3 | Weighted median | -0.046 | 0.031 | -0.106 | 0.014 | 0.130 | 0.955 | 0.899 | 1.014 | 0.518 |
|  |  | 3 | MR-Egger | -0.077 | 0.083 | -0.239 | 0.085 | 0.351 | 0.926 | 0.787 | 1.089 | 0.757 |
|  | Stroke | 3 | IVW | -0.005 | 0.027 | -0.059 | 0.048 | 0.851 | 0.995 | 0.943 | 1.050 | 0.971 |
|  |  | 3 | Weighted median | -0.005 | 0.032 | -0.068 | 0.057 | 0.866 | 0.995 | 0.935 | 1.058 | 0.971 |
|  |  | 3 | MR-Egger | 0.004 | 0.092 | -0.176 | 0.185 | 0.965 | 1.004 | 0.838 | 1.203 | 1.000 |
| IL1R1 | COPD | 5 | IVW | 0.006 | 0.024 | -0.040 | 0.053 | 0.786 | 1.006 | 0.961 | 1.054 | 0.971 |
|  |  | 5 | Weighted median | 0.000 | 0.015 | -0.029 | 0.030 | 0.982 | 1.000 | 0.971 | 1.030 | 1.000 |
|  |  | 5 | MR-Egger | -0.028 | 0.131 | -0.283 | 0.228 | 0.833 | 0.973 | 0.753 | 1.256 | 0.971 |
|  |  | 3 | MR-PRESSO | -0.008 | 0.012 | -0.032 | 0.016 | 0.580 | 0.992 | 0.968 | 1.016 | 0.896 |
|  | Asthma | 5 | IVW | 0.186 | 0.158 | -0.123 | 0.496 | 0.238 | 1.205 | 0.884 | 1.642 | 0.653 |
|  |  | 5 | Weighted median | 0.367 | 0.050 | 0.268 | 0.465 | 0.000 | 1.443 | 1.308 | 1.592 | 0.000 |
|  |  | 5 | MR-Egger | 0.867 | 0.779 | -0.660 | 2.393 | 0.266 | 2.379 | 0.517 | 10.948 | 0.686 |
|  |  | 1 | MR-PRESSO | 0.281 | NA | NA | NA | NA | 1.324 | NA | NA | NA |
|  | CAD | 5 | IVW | 0.084 | 0.033 | 0.020 | 0.148 | 0.010 | 1.088 | 1.020 | 1.160 | 0.110 |
|  |  | 5 | Weighted median | 0.058 | 0.034 | -0.008 | 0.124 | 0.084 | 1.060 | 0.992 | 1.132 | 0.409 |
|  |  | 5 | MR-Egger | -0.127 | 0.133 | -0.387 | 0.133 | 0.339 | 0.881 | 0.679 | 1.142 | 0.750 |
|  |  | 5 | MR-PRESSO | 0.084 | 0.033 | 0.020 | 0.148 | 0.061 | 1.088 | 1.020 | 1.160 | 0.342 |
|  | MI | 5 | IVW | 0.123 | 0.057 | 0.012 | 0.234 | 0.029 | 1.131 | 1.012 | 1.264 | 0.224 |
|  |  | 5 | Weighted median | 0.110 | 0.051 | 0.010 | 0.210 | 0.031 | 1.117 | 1.010 | 1.234 | 0.230 |
|  |  | 5 | MR-Egger | 0.015 | 0.310 | -0.592 | 0.622 | 0.962 | 1.015 | 0.553 | 1.862 | 0.999 |
|  |  | 5 | MR-PRESSO | 0.123 | 0.057 | 0.012 | 0.234 | 0.095 | 1.131 | 1.012 | 1.264 | 0.445 |
|  | HF | 5 | IVW | -0.058 | 0.041 | -0.137 | 0.022 | 0.155 | 0.944 | 0.872 | 1.022 | 0.557 |
|  |  | 5 | Weighted median | -0.069 | 0.048 | -0.162 | 0.025 | 0.149 | 0.933 | 0.850 | 1.025 | 0.548 |
|  |  | 5 | MR-Egger | 0.069 | 0.197 | -0.317 | 0.455 | 0.726 | 1.071 | 0.728 | 1.576 | 0.948 |
|  |  | 5 | MR-PRESSO | -0.058 | 0.025 | -0.106 | -0.010 | 0.078 | 0.944 | 0.900 | 0.990 | 0.395 |
|  | AF | 5 | IVW | -0.073 | 0.041 | -0.153 | 0.007 | 0.074 | 0.930 | 0.858 | 1.007 | 0.382 |
|  |  | 5 | Weighted median | -0.068 | 0.045 | -0.156 | 0.020 | 0.127 | 0.934 | 0.855 | 1.020 | 0.516 |
|  |  | 5 | MR-Egger | -0.056 | 0.228 | -0.503 | 0.390 | 0.805 | 0.945 | 0.605 | 1.477 | 0.971 |
|  |  | 5 | MR-PRESSO | -0.073 | 0.041 | -0.153 | 0.007 | 0.149 | 0.930 | 0.858 | 1.007 | 0.548 |
|  | Stroke | 5 | IVW | 0.072 | 0.036 | 0.001 | 0.142 | 0.048 | 1.074 | 1.001 | 1.153 | 0.300 |
|  |  | 5 | Weighted median | 0.079 | 0.045 | -0.009 | 0.168 | 0.080 | 1.082 | 0.991 | 1.183 | 0.399 |
|  |  | 5 | MR-Egger | 0.248 | 0.179 | -0.103 | 0.599 | 0.166 | 1.282 | 0.902 | 1.821 | 0.576 |
|  |  | 5 | MR-PRESSO | 0.072 | 0.031 | 0.010 | 0.133 | 0.086 | 1.074 | 1.010 | 1.143 | 0.413 |

Blue highlight denotes suggestive evidence based on FDR-corrected p-value (q-value). Orange highlight denotes strong evidence based on FDR-corrected p-value (q-value).

CI95Lower: 95% lower CI for the estimate; Ci95Upper: 95% upper CI for the estimate. ORlow: 95% lower CI for the OR; ORhi: 95% upper CI for the OR. AF: atrial fibrillation, CAD: coronary artery disease, COPD: chronic obstructive pulmonary diseases, HF: heart failure, IS: ischemia stroke, MI: myocardial infarction, n: number of SNPs used as instrument variables in each method, SNPs single nucleotide polymorphism, CI confidence intervals, OR odds ratio.
